# The impact of later trading hours for bars and clubs on alcohol-related ambulance call-outs and crimes in Scotland: a controlled interrupted time series study

**DOI:** 10.1101/2025.10.30.25339133

**Authors:** Nurnabi Sheikh, Jim Lewsey, Ines Henriques-Cadby, Colin Angus, Francesco Manca, Houra Haghpanahan, Emma McIntosh, Gemma Mitchell, Megan Cook, Karen Maxwell, Andrea Mohan, Isabelle Uny, Elaina Smith, James Nicholls, Carol Emslie, Rachel O’Donnell, Niamh Fitzgerald

**Affiliations:** Health Economics and Health Technology Assessment (HEHTA), School of Health and Wellbeing, University of Glasgow, United Kingdom; Department of Mathematics, The University of Manchester, United Kingdom; School of Medicine and Population Health, University of Sheffield, United Kingdom; Institute for Social Marketing & Health, University of Stirling, United Kingdom; Centre for Alcohol Policy Research, La Trobe University, Melbourne, Australia; Research Centre for Health, School of Health and Life Sciences, Glasgow Caledonian University, Glasgow, Scotland, United Kingdom; School of Health Sciences, University of Dundee, Dundee, United Kingdom; Health Improvement, NHS Greater Glasgow and Clyde, United Kingdom

**Keywords:** later alcohol trading hours, alcohol-related ambulance call-outs, crimes, controlled interrupted time series, Scotland

## Abstract

**Background:** Alcohol-related harms are prevalent late at night, especially on weekends, when high levels of intoxication contribute to increased rates of injury and violence. Reducing or increasing alcohol trading hours late at night in bars and clubs is generally associated with reduced and increased harms, respectively. This study evaluates the impact of later alcohol trading hours in the Scottish cities of Aberdeen and Glasgow on alcohol-related ambulance call-outs and crimes. Under local policy changes, 38 bars in Aberdeen had trading hours extended between 1 to 3 hours up to 3am, and 10 nightclubs in Glasgow had a 1-hour extension to 4am.

**Methods:** Following a natural experiment evaluation framework, we used a controlled interrupted time series design to compare outcomes before and after policy changes, from 2015 to 2021. The primary outcome was a count of total weekend night-time alcohol-related ambulance call-outs. Secondary outcomes included weekend night-time crimes.

**Results:** In Aberdeen, the policy led to a significant relative increase of 17·4% (effect size=7·061; 95% CI=3·902 to 10·220; p<0·001) in alcohol-related ambulance call-outs, and 7·9% (effect size=3·187; 95% CI=0·564 to 5·810; p=0·017) in reported crimes (not robust across sensitivity analyses), at weekend night-times compared to Edinburgh (control). Findings were not significant and robust across analyses for Glasgow.

**Conclusion:** Later alcohol trading hours had a significant negative impact on alcohol-related ambulance call-outs in Aberdeen (where more premises had longer extensions) but not in Glasgow, suggesting the number, capacity and type of premises moderated outcomes. This is important for the design of future national and local licensing policies and regulations.

**KEY MESSAGES:** *What is already known on this topic?:* Prior research has shown a strong association between alcohol availability, including late-night trading hours, and alcohol-related harms. Systematic reviews report that reducing hours of alcohol sales is linked to decreased alcohol-related ambulance-callouts, assaults, and road accidents, whereas later trading hours tend to intensify these alcohol-related harms. International studies, such as those conducted in Netherlands, Norway, and Australia, have provided empirical evidence that even modest reductions or extensions in trading hours can lead to significant decreases or increases in alcohol-related ambulance call-outs, crimes, and emergency department visits.

*What this study adds?:* This study provides novel and robust evidence on the impact of later trading hours for bars and clubs on alcohol-related ambulance call-outs and reported crimes in Scotland, using a controlled interrupted time series (CITS) design. By focusing on local policy changes in both Aberdeen and Glasgow, where certain venues of different types were permitted to extend their alcohol trading hours, this research leverages a natural experiment to provide novel insights. Our evaluation provides new evidence on the importance of local contexts on the magnitude and timing of change in trading hours for informing future policymaking decisions and research.

*How this study might affect research, practice or policy?:* Our findings raise important questions for policymakers seeking to balance economic and community interests in late-night entertainment with the negative impacts of such premises on services including: what is a proportionate policy given our evidence? Is it legally possible for current licensing systems to contain later trading hours to a subset of premises? And, if so, how would those premises be selected?

## INTRODUCTION

Alcohol consumption led to approximately 1·8 million deaths globally in 2021 and is a risk factor for over 200 diseases [1,2]. Alcohol’s harms extend beyond those consuming alcohol, including in contributing to increased crime, violence, road accidents, and various forms of injury [3,4]. In Scotland, 1,277 deaths from causes wholly attributable to alcohol were recorded in 2023, averaging 24 deaths weekly [5]. Alcohol also poses a significant burden for the healthcare system in Scotland: in 2022/23, there were 31,206 alcohol-specific hospital admissions [6] and in 2019 it was estimated that 16% of ambulance call-outs were alcohol-related [7]. Generally, alcohol harms are associated with greater density of premises licensed to sell alcohol, as well as increased days and hours during which alcohol can legally be sold [8]. Acute alcohol harms are particularly prevalent late at night (i.e., 12am to 5am), especially on weekends, when higher levels of intoxication, can contribute to increased rates of injuries and violence [9]. In 2019, the highest concentration of alcohol-related ambulance call-outs in Scotland occurred between 9pm and 1am on weekends, whereas call-outs unrelated to alcohol were more frequent between 10am and 9pm [7].

Reducing late-night trading hours for alcohol on-premises outlets is associated with reductions in alcohol harms [10]. Considering extensions to permitted trading hours for alcohol in on-premises outlets, separate systematic reviews concluded that extensions generally increased the likelihood of assaults, injuries, and drink-driving incidents [11,12]. A 2009 review found evidence that extensions were associated with increased harm, while reductions was associated with a decrease [13]. The review emphasised the need for well-controlled research to further verify these associations. Another review based on a small number of studies concluded that extensions of two or more hours per day significantly increased alcohol harms, although no strong association was found for shorter spanning extensions due to limited evidence [14].

In the UK, three separate systems (one in England and Wales, one in Scotland and one in Northern Ireland) control the granting of licences required to legally sell alcohol. Both the systems in England/Wales and in Scotland were substantially reformed in the mid-2000s. The Licensing Act (Scotland) 2005 introduced a wholly reformed system empowering local government licensing boards to set local policy and make decisions on premises licensing, guided by five overarching objectives. These boards set local on-trade alcohol trading hours policy, but the legislation includes a presumption against 24-hour licensing. Under this system, late night alcohol trading hours have expanded in recent years [15], but their impact has never been evaluated in Scotland. The Licensing Act (2003) also established a completely new local government-led system to manage licensing in England and Wales that created a general presumption in favour of granting premises licences, with no presumption against 24-hour alcohol sales. The 2003 Act was underpinned by the view that the fixed closing times that predated the Act (most pubs closing at 11pm) contributed to overcrowding, violence, and disorder and that staggered dispersal may reduce these issues [16]. Studies on the impact of extended licensing in England and Wales have not found consistent evidence of their impact on violence [15], they used weaker before-and-after designs without considering a control series compared to those in other international studies [13,16]. Using a more robust interrupted time series (ITS) design, Humphreys and colleagues found no evidence of an overall increase in violent crime in the city of Manchester following the Act but a significant 36% increase between 3am and 6am [17]. Although more robust than earlier studies, wider health outcomes were not considered, further evidencing the need for additional research [17]. Another UK-based study reported small changes in the timing of self-reported drinking occasions due to the implementation of the Act but no evidence of changes in finish time variation, post-loading behaviour, late-night drinking, or overall alcohol consumption [18]. To our knowledge, no study in the UK has considered the impact of later trading hours on alcohol-related ambulance call-outs.

Under the system established by the 2005 Act, local Licensing Boards (made up of elected members of local government) in Aberdeen and Glasgow have in recent years changed their local policies to allow premises in certain licence categories to apply for, and be granted, permission to sell alcohol later at night. This study aims to evaluate the impact of these later trading hours for bars and clubs on alcohol-related ambulance call-outs and reported crimes in both cities.

## METHODS

### The policy changes: later trading hours for bars and clubs in Glasgow and Aberdeen

In Glasgow, from 12^th^ April 2019, 10 nightclubs, which met certain quality and operating standards, were granted permission by the Glasgow City Licensing Board (GLB) to remain open and sell alcohol for an additional hour to 4am (instead of 3am). This initial 12-month pilot became standard policy in 2023. In Aberdeen, a policy was introduced in January 2017 that allowed a wider range of alcohol premises to apply for later trading hours, which had previously only been available to nightclubs. From March 2017 to October 2020, 38 bars/pubs in Aberdeen were granted permission to sell alcohol between 1 and 2 hours later at night, up to 3am. Nightclubs remained closed throughout the COVID-19 restrictions and were therefore unable to use the additional hour, while bars and pubs were permitted to reopen earlier and to use extended hours during some periods. These recent policy changes in Aberdeen and Glasgow serve as the basis for this natural experiment evaluation.

### Study Design

We used a controlled interrupted time series (CITS) design to evaluate the later closing times, comparing outcomes before and after policy implementation (May 2015-July 2022) in Glasgow and Aberdeen with a control group. The policy change date, 12 April 2019 for Glasgow, and a gradual adoption of extended hours in Aberdeen beginning in early 2017, being the "interruption" in the time series.

### Data Sources

We obtained all ambulance call-outs in Scotland from the Scottish Ambulance Service (SAS) and crime data from Police Scotland. Ambulance data included records such as call-out time, coordinates of location, patient demographics, and a flag indicating whether a call-out was alcohol-related (algorithmically generated from free-text fields in call-out records), as previously developed with SAS [7]. Crime data were gathered from Police Scotland using Scottish Government Justice Directorate (SGJD) crime codes. Two authors (NF & JL) scrutinised and selected crime codes they judged to be more likely to be associated with alcohol, in consultation with Police Scotland. These included assaults, drunkenness, driving under the influence of alcohol, and antisocial behaviour offences (see Table A4 in the appendix 2.1). Variables included time of recording, crime type, local council name, incident date, and coordinates of crime location.

### Outcomes

Primary outcome: total weekend night-time alcohol-related ambulance call-outs (time between Fridays 20:00 to 23:59, Saturdays 00:00 to 05:59 and 20:00 to 23:59, and Sundays 00:00 to 05.59). Secondary outcomes: total weekend night-time ambulance call-outs and reported crimes.

The outcomes were analysed as weekly counts. We also modelled population size-adjusted outcomes to check robustness of our analysis.

### Control Selection

We developed a principled and structured framework for selecting the study control, shown in Figure A1 in the appendix 1 (with steps in the appendix 1.1 for alcohol-related ambulance call-outs and in the appendix 1.2 for reported crimes) and details in the published statistical analysis plan (SAP) [19]. Following this framework, among the potential 30 control candidates (Scottish council areas, apart from Glasgow City and Aberdeen City), the City of Edinburgh council area (Edinburgh) was identified as the most suitable control for Aberdeen and Glasgow.

### Measurement of Policy Exposure Variable

Separate exposure variables were generated for Glasgow and Aberdeen. The dates on which premises were permitted to trade later were staggered over time in Aberdeen; in contrast, in Glasgow, the later trading hours came into effect from the same date for all premises. In Glasgow, a binary exposure variable taking the value “0”, for time periods before the implementation of the new trading hours (i.e., 12th April 2019), and “1” for time periods thereafter, was used. In Aberdeen, a numerical measure of exposure was developed based on the additional person-hours available weekly across all premises. This measure reflects the staggered granting of later licensed hours, taking values between 0 (no extended hours) and 1 (all additional person-hours had been adopted across participating premisses). The potential additional person-hours for a given venue in a week was calculated by using the formula [(additional hours) x (the number of nights additional hours are permitted to be used in a week) x (capacity of the venue)]. Full details can be found in the SAP [19].

### Confounding Variables

We included data on per capita gross disposable household income, weather conditions (mean temperature, rainfall), and total number of on-premises alcohol outlets to account for potential time-varying confounding effects. Additional adjusting for COVID-19 lockdown (imposing closure or restricted hours for alcohol-premises), was done through dummy variables. We included dummy variables to account for public holidays, outliers and COVID-19 restrictions which meant that the premises with later hours were not open at certain times (see appendix 2.2 for details). As noted earlier, the value of the COVID-19 dummy variable over time reflected that nightclubs were fully closed during COVID-19 restrictions, while bars and pubs remained open for some periods. Data sources for the confounding variables are provided in appendix 2.2.1.

### Statistical Analysis

Data were analysed using autoregressive integrated moving average (ARIMA) models for both primary and secondary outcomes following the CITS design. We modelled the outcome differences between the interventions and control over time, for example, differences between alcohol-related ambulance call-outs in Aberdeen and Edinburgh. We followed Box-Jenkins model selection steps (identification, estimation, and diagnostic checking) to obtain the best ARIMA model that adequately captures the underlying patterns in the data and provides accurate estimation. First, we used an Augmented Dickey-Fuller test to establish that the stationarity (i.e., constant mean and variance over time) assumption was met. Then, we used autocorrelation and partial autocorrelation functions (ACF and PACF) to identify the terms of the autoregressive (AR) and moving average (MA) models. After that, from the candidate models, the best fit was chosen according to Bayesian (BIC) and Akaike (AIC) Information Criterions. We next tested suitability of the fitted model by performing diagnostic checks to ensure the residuals presented white noise characteristics. Policy exposure variables were added to the model as covariates to estimate intervention effect sizes.

To check the robustness of our findings, we examined the effect size across ARIMA models adjusting for all covariates, and for statistically significant covariates only.

Subgroup analyses to examine differential impacts by age, and sex, were conducted for the primary outcome only. Analyses were performed in Stata/MP 17.0 and RStudio (version 4.2.0) using the *Synth* package [20].

### Sensitivity analysis

We conducted sensitivity analyses by restricting the time series to pre-COVID-19 restriction periods in Scotland to assess the robustness of our estimates. Additionally, we modelled population size-adjusted outcome rates. To check robustness of our estimation, we additionally created two binary exposure variables specific to Aberdeen: one similar to Glasgow on the introduction of the new policy, and a second to indicate when the policy reached at least half its “strength”, taking a value of "0" when Aberdeen’s policy exposure variable is less than 0·5, and "1" otherwise.

We also conducted Synthetic Control (SC) modelling. A separate SC for Aberdeen and Glasgow was created for the population-adjusted rates of alcohol-related ambulance call-outs and reported crimes, applying various specifications of pre-intervention outcomes [21]. Covariates such as per-capita gross disposable household income and population-adjusted number of on-premises alcohol outlets were incorporated in the construction of the SC. To assess the validity of our SC models, we split the pre-intervention period into training and validation phases and compared MSPEs (across validation periods) for the different model specifications. Appendices 3.1.1 and 3.2.1 present the model specifications and validity tests for alcohol-related ambulance call-outs in Aberdeen and Glasgow, respectively. Appendices 3.3.1 and 3.4.1 contain the model specifications and validity tests for reported crimes in Aberdeen and Glasgow, respectively. We performed in-space placebo tests and calculated frequentist (frequency-based) p-values for the effect sizes generated through SC. Details of synthetic control analyses and sensitivity tests are provided in appendices 3.1.2 (Aberdeen) and 3.2.2 (Glasgow) for alcohol-related ambulance call-outs, and in appendices 3.3.2 (Aberdeen) and 3.4.2 (Glasgow) for reported crimes. Additionally, we performed time-series models (ARIMA) to the synthetic control series to check the robustness of the estimated effect size (see Tables 17 and 18 in the appendix 4). Finally, a falsification test was conducted, by generating a false policy exposure variable signalling exposure one year before the true intervention date.

## RESULTS

Between 1 May 2015 and 31 July 2022, a total of 6,461 alcohol-related ambulance call-outs occurred in Aberdeen, 19,956 in Glasgow, and 13,781 in Edinburgh during weekend night-times (Table A1 in the appendix 2.1). A total of 5,927 crimes were reported in Aberdeen, 20,038 in Glasgow, and 12,268 in Edinburgh during weekend night-times, more than 80% of which in each city were ‘common assaults’ (Table A3 and A4 in the appendix 2.1).

Prior to the policy changes, trends in alcohol-related ambulance call-outs and reported crimes between Aberdeen/Glasgow and Edinburgh were broadly comparable, with seasonal variation (Figure 1).

**Figure 1:**
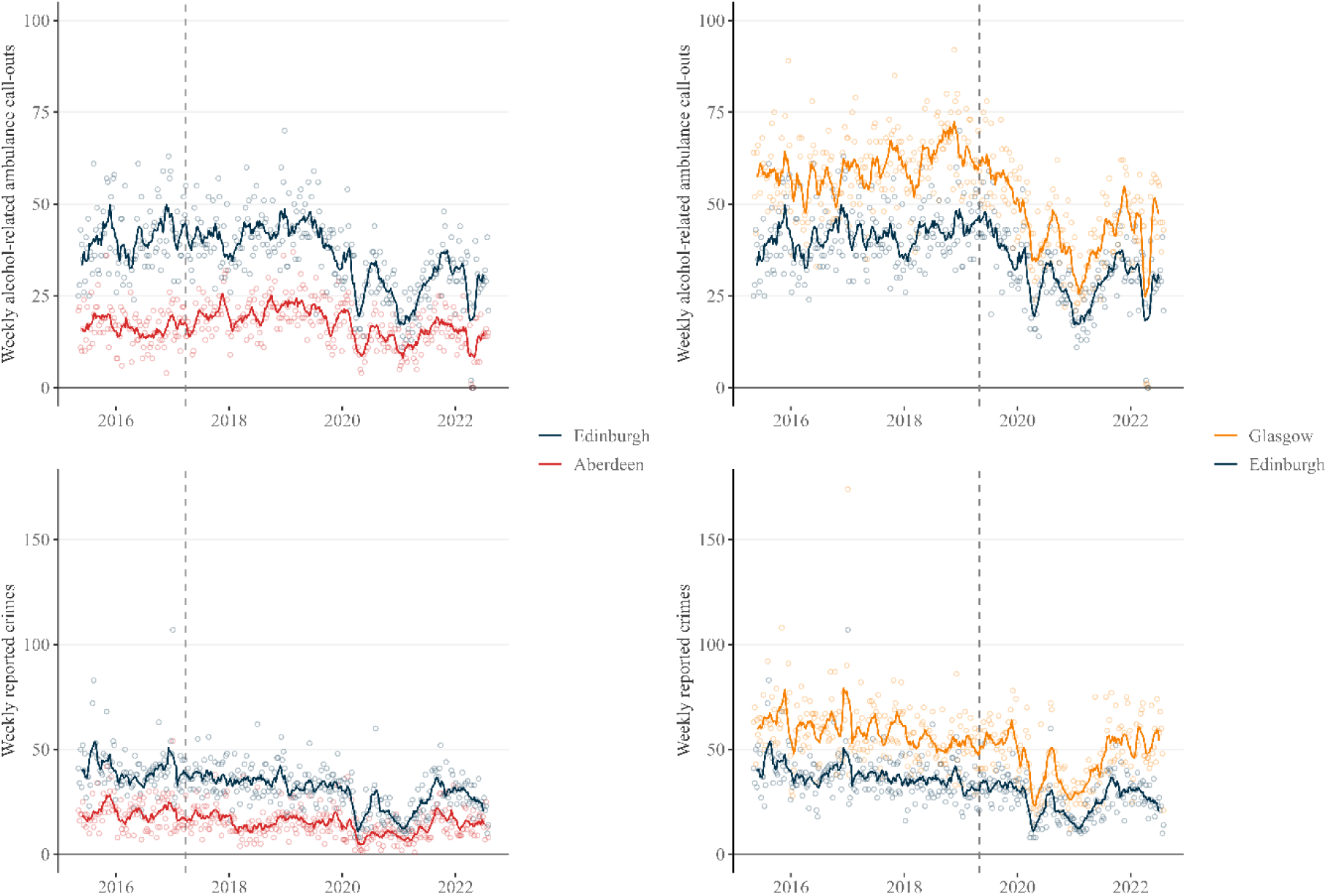
Number of weekend night-time alcohol-related ambulance call-outs and reported crimes over-time in the intervention and control cities.

In Aberdeen, alcohol-related ambulance call-outs peaked between 00:00–00:59 before the new policy but shifted to 01:00–01:59 after the policy change (Figure 2); such a shift was not observed in the control city (Edinburgh) and for reported crimes (Figure A2 in the appendix 2.1).

**Figure 2.**
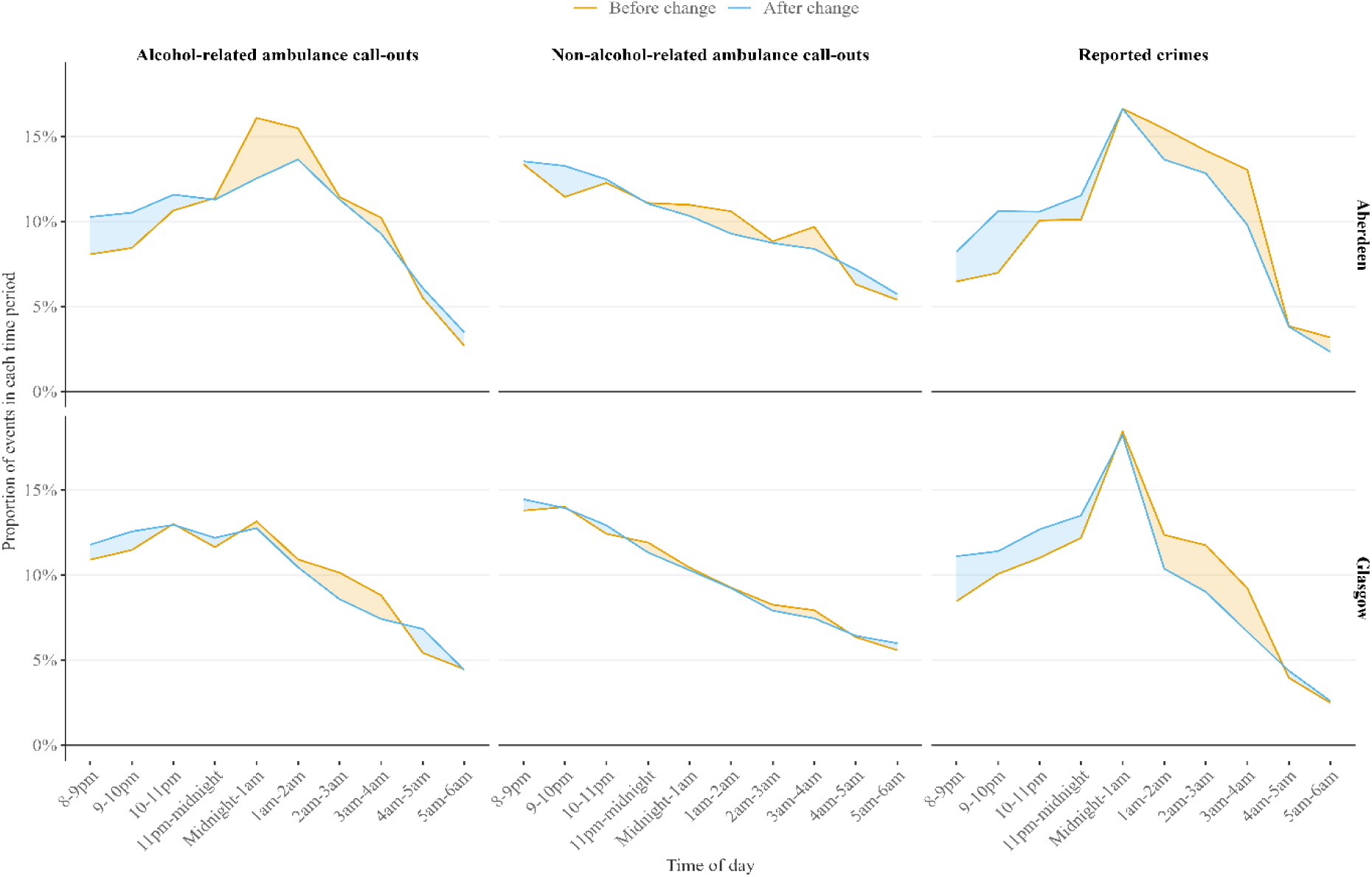
Distribution of weekend night-time alcohol-related ambulance call-out and reported crimes in Aberdeen and Glasgow.

The estimated policy effect for Aberdeen using a staggered policy variable when the model included only statistically significant covariates, was a significant average relative increase of 7·061 extra call-outs per week in Aberdeen following the policy change compared with Edinburgh (corresponding to a 17·4% relative increase in Aberdeen compared to Edinburgh) (Table 1). Increasing policy effects were consistently observed for Aberdeen across various sensitivity analyses. However, the policy effect term was not significant when using the SC method. Subgroup analyses revealed that the policy effects in Aberdeen were comparatively higher for males (coefficient = 3·726, p-value <0·001, 95% CI = 1·946 to 5·507) and individuals under 45 years of age (coefficient = 7·050, p-value<0·001, 95% CI = 4·077 to 10·023) compared to their respective counterparts (Tables A9 to A12 in the appendix 2.3). Similarly, when only statistically significant covariates were included in the model, the effect was 3·187 (p-value = 0·017, 95% CI = 0·564 to 5·810) for reported crimes in Aberdeen (Table 2). This means an estimated average of 3·187 extra reported crimes per week in Aberdeen than Edinburgh, following the policy change (a 7·9% relative increase of reported crimes for Aberdeen compared to Edinburgh). Across our sensitivity analyses, the estimated policy effects show consistent increases after the policy change but are not consistently significant at 5% significance level (Table 2).

**Table 1.** Effect of policy changes on weekend night-time alcohol-related ambulance call-outs in Aberdeen and Glasgow.

| Primary outcome,<br>weekend night-time<br>alcohol-related<br>ambulance callouts | Main analysis |  |  | Sensitivity analysis |  |  |  |  |  | Synthetic control |  |
| --- | --- | --- | --- | --- | --- | --- | --- | --- | --- | --- | --- |
|  | Outcome: number of incidents |  |  | Restricted time series only before<br>COVID-19 restrictions, (outcome:<br>number of incidents) |  |  | Outcome: population adjusted<br>incident rates |  |  | Outcome:<br>population<br>adjusted incident<br>rates |  |
|  | Coefficient | P-value | 95% CI | Coefficient | P-value | 95% CI | Coefficient | P-value | 95% CI | Coefficient | P-value |
| Intervention area: Aberdeen |  |  |  |  |  |  |  |  |  |  |  |
| **Staggered policy**<br>**covariate** |  |  |  |  |  |  |  |  |  |  |  |
| Policy effect (all<br>covariates) | 5·824 | 0·002 | 2·225, 9·423 | 4·641 | 0·037 | 0·279, 9·002 | 0·140 | <0·001 | 0·071, 0·209 | 0·103 | 0·222 |
| Policy effect<br>(significant covariates) | 7·061 | <0·001 | 3·902, 10·220 | 4·643 | 0·036 | 0·292, 8·994 | 0·130 | <0·001 | 0·061, 0·198 |  |  |
| **Dummy policy**<br>**covariate, sensitivity**<br>**analysis** |  |  |  |  |  |  |  |  |  |  |  |
| Policy effect (all<br>covariates) | 2·898 | 0·056 | -0·078, 5·873 | 2·486 | 0·101 | -0·483, 5·455 | 0·119 | <0·001 | 0·052, 0·185 | 0·140 | 0·185 |
| Policy effect<br>(significant covariates) | 2·970 | 0·051 | -0·014, 5·955 | 2·565 | 0·098 | -0·470, 5·599 | 0·126 | <0·001 | 0·072, 0·180 |  |  |
| **Dummy policy**<br>**covariate with policy**<br>**implemented with at**<br>**least half strength,**<br>**sensitivity analysis** |  |  |  |  |  |  |  |  |  |  |  |
| Policy effect (all<br>covariates) | 3·145 | 0·041 | 0·136, 6·154 | 2·018 | 0·243 | -1·369, 5·406 | 0·088 | 0·007 | 0·024, 0·152 | - | - |
| Policy effect<br>(significant covariates) | 4.473 | 0.001 | 1.846, 7.100 | 1.744 | 0.305 | -1.585, 5.072 | 0.080 | 0.011 | 0.018, 0.142 | - | - |
| **Intervention area: Glasgow** |  |  |  |  |  |  |  |  |  |  |  |
| **Dummy policy covariate** |  |  |  |  |  |  |  |  |  |  |  |
| Policy effect (all covariates) | -4.016 | 0.050 | -8.027, -0.005 | -2.307 | 0.329 | -6.942, 2.329 | -0.057 | 0.064 | -0.116, 0.003 | 0.012 | 0.926 |
| Policy effect<br>(significant covariates) | -5.110 | <0.001 | -7.491, -2.730 | -2.951 | 0.096 | -6.423, 0.520 | -0.045 | 0.018 | -0.081, -0.008 |  |  |

*\*Effect size for synthetic control adjusted by the covariates, number of on-premises alcohol outlets (population adjusted), and per capita gross disposable household income*

**Table 2.**
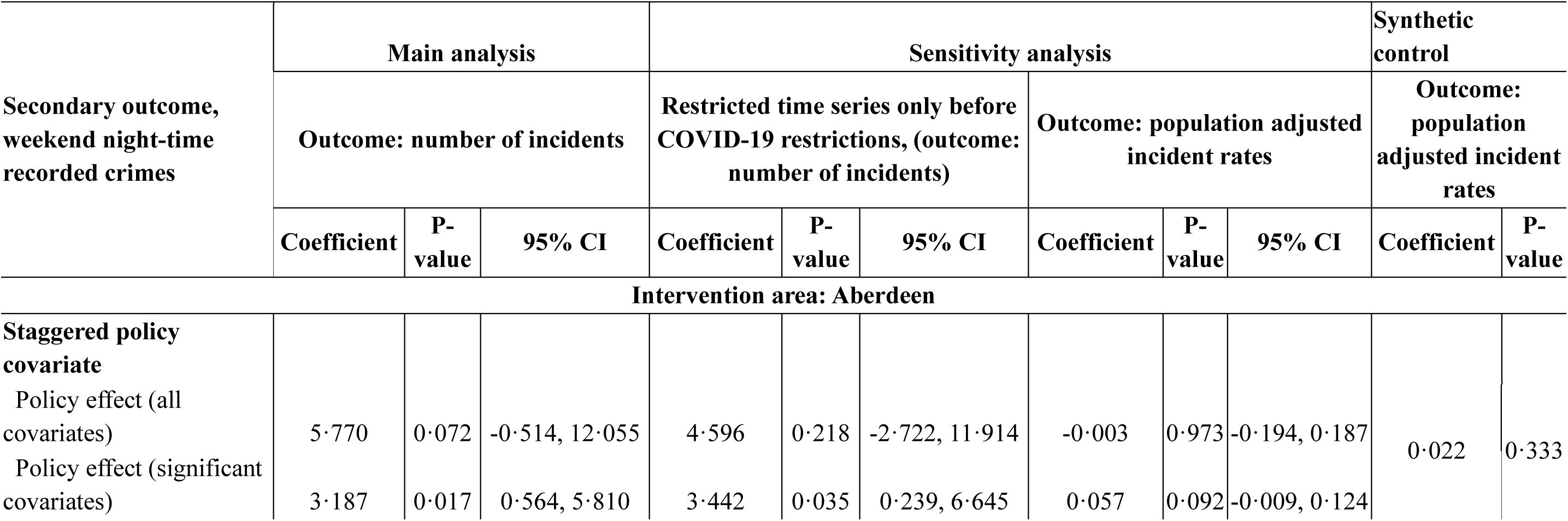
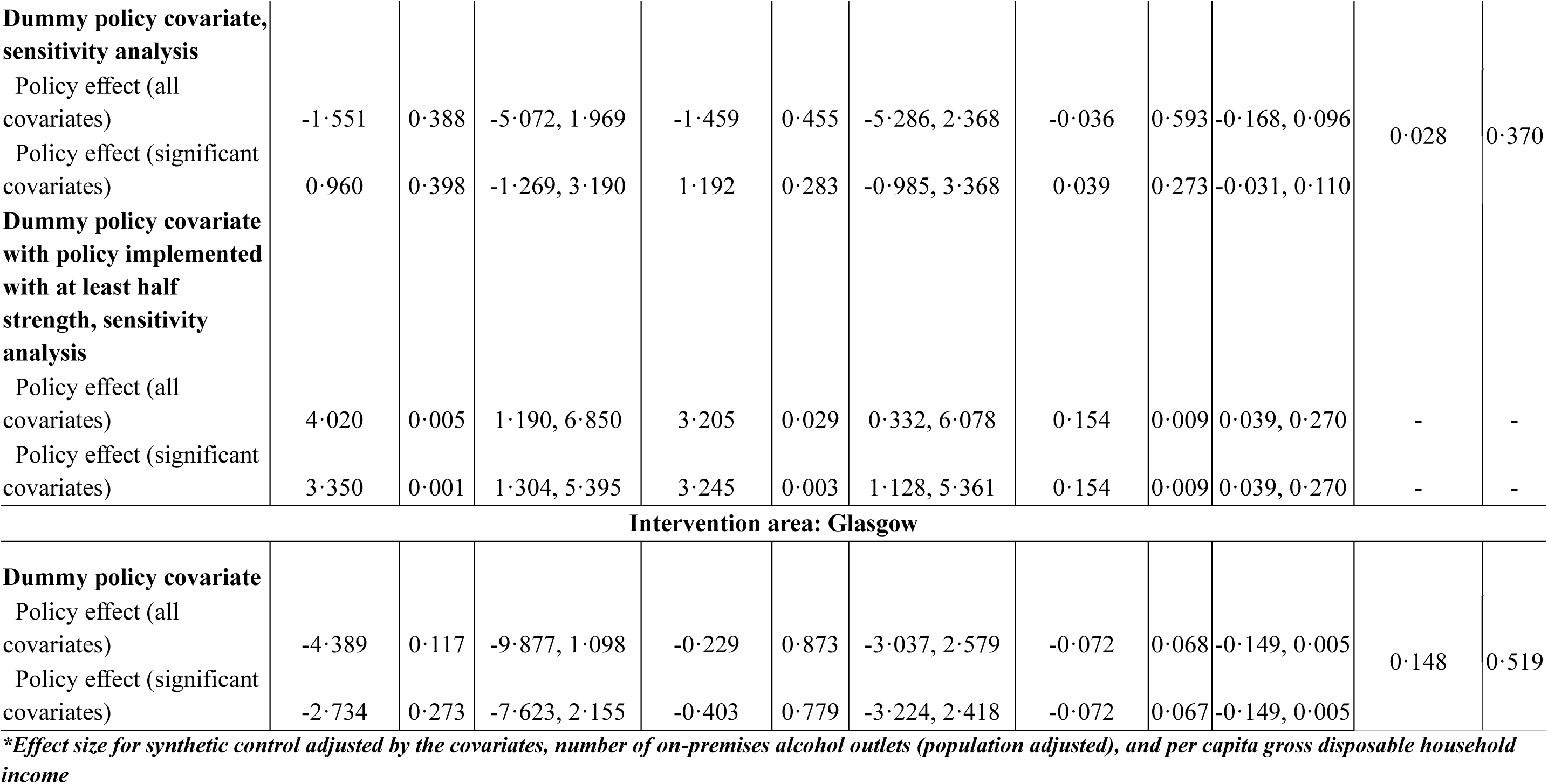
Effect of policy changes on weekend recorded crimes in Aberdeen and Glasgow.

| Secondary outcome,<br>weekend night-time<br>recorded crimes | Main analysis |  |  | Sensitivity analysis |  |  |  |  |  | Synthetic control |  |
| --- | --- | --- | --- | --- | --- | --- | --- | --- | --- | --- | --- |
|  | Outcome: number of incidents |  |  | Restricted time series only before COVID-19 restrictions, (outcome: number of incidents) |  |  | Outcome: population adjusted incident rates |  |  | Outcome: population adjusted incident rates |  |
|  | Coefficient | P-value | 95% CI | Coefficient | P-value | 95% CI | Coefficient | P-value | 95% CI | Coefficient | P-value |
| **Intervention area: Aberdeen** |  |  |  |  |  |  |  |  |  |  |  |
| **Staggered policy covariate** |  |  |  |  |  |  |  |  |  |  |  |
| Policy effect (all covariates) | 5.770 | 0.072 | -0.514, 12.055 | 4.596 | 0.218 | -2.722, 11.914 | -0.003 | 0.973 | -0.194, 0.187 | 0.022 | 0.333 |
| Policy effect (significant covariates) | 3.187 | 0.017 | 0.564, 5.810 | 3.442 | 0.035 | 0.239, 6.645 | 0.057 | 0.092 | -0.009, 0.124 |  |  |
| **Dummy policy covariate, sensitivity analysis** |  |  |  |  |  |  |  |  |  |  |  |
| Policy effect (all covariates) | -1.551 | 0.388 | -5.072, 1.969 | -1.459 | 0.455 | -5.286, 2.368 | -0.036 | 0.593 | -0.168, 0.096 | 0.028 | 0.370 |
| Policy effect (significant covariates) | 0.960 | 0.398 | -1.269, 3.190 | 1.192 | 0.283 | -0.985, 3.368 | 0.039 | 0.273 | -0.031, 0.110 |  |  |
| **Dummy policy covariate with policy implemented with at least half strength, sensitivity analysis** |  |  |  |  |  |  |  |  |  |  |  |
| Policy effect (all covariates) | 4.020 | 0.005 | 1.190, 6.850 | 3.205 | 0.029 | 0.332, 6.078 | 0.154 | 0.009 | 0.039, 0.270 | - | - |
| Policy effect (significant covariates) | 3.350 | 0.001 | 1.304, 5.395 | 3.245 | 0.003 | 1.128, 5.361 | 0.154 | 0.009 | 0.039, 0.270 | - | - |
| **Intervention area: Glasgow** |  |  |  |  |  |  |  |  |  |  |  |
| **Dummy policy covariate** |  |  |  |  |  |  |  |  |  |  |  |
| Policy effect (all covariates) | -4.389 | 0.117 | -9.877, 1.098 | -0.229 | 0.873 | -3.037, 2.579 | -0.072 | 0.068 | -0.149, 0.005 | 0.148 | 0.519 |
| Policy effect (significant covariates) | -2.734 | 0.273 | -7.623, 2.155 | -0.403 | 0.779 | -3.224, 2.418 | -0.072 | 0.067 | -0.149, 0.005 |  |  |

*\*Effect size for synthetic control adjusted by the covariates, number of on-premises alcohol outlets (population adjusted), and per capita gross disposable household income*

In contrast, Glasgow showed a significant decreasing policy effect on alcohol-related ambulance call-outs (i.e., a reduction in 5·110 call-outs per week) based on ARIMA models (Table 1). However, Glasgow results varied substantially across the range of sensitivity analyses. Moreover, for reported crimes in Glasgow, no significant policy effects were observed (Table 2).

We also conducted falsification tests for both primary and secondary outcomes, results are provided in the appendix (Tables A14 to A16 in the appendix 2.4).

In Figure 3, we compared the rates of weekend night-time alcohol-related ambulance call-outs and reported crimes in Aberdeen and Glasgow with their respective synthetic counterparts (a counterfactual scenario that assumes no policy changes occurred). For alcohol-related ambulance call-out rates, the observed and synthetic trends in Aberdeen prior to the intervention show the best fit compared to Glasgow. After the policy change, the observed rates in Aberdeen mostly lie above the synthetic rates, suggesting an increase in call-outs due to the policy. In contrast, Glasgow shows considerable overlap between observed and synthetic rates after the policy change, indicating no clear effect.

**Figure 3:**
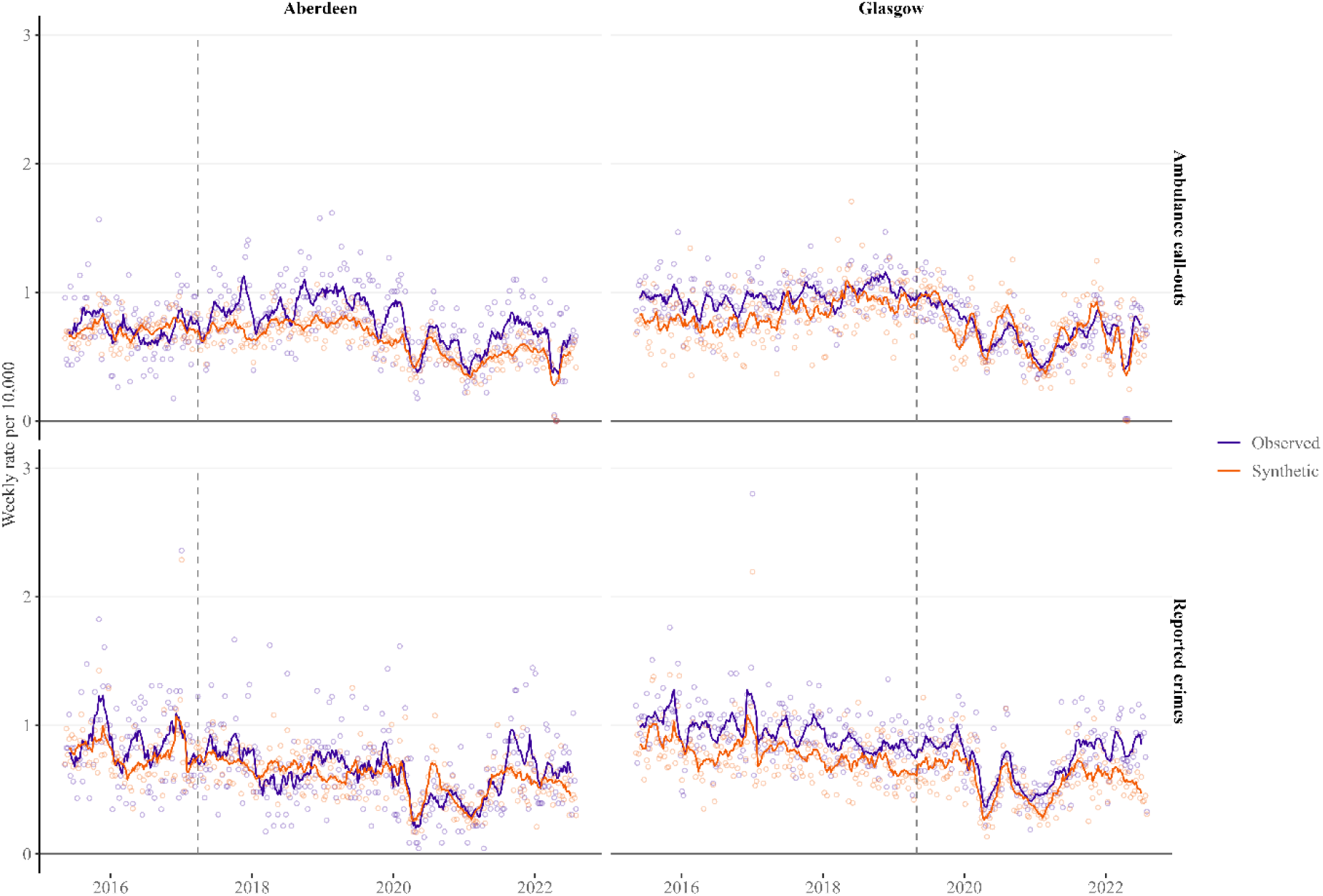
Rate of weekend night-time alcohol-related ambulance call-outs and reported crimes in Aberdeen, Glasgow and synthetic Aberdeen and synthetic Glasgow. * Dots represent weekly data; solid lines represent 8-week moving averages; vertical dashed line represents start of intervention

For reported crime rates, although some overlap exists between the observed and synthetic trends in Aberdeen after the policy change, the observed rates lie above the synthetic rates at most time points, suggesting a potential increase. In Glasgow, the observed crime rates also mostly lie above the synthetic rates following the policy change, implying an increase. However, the pre-intervention fit between observed and synthetic crime rates is weaker in Glasgow than in Aberdeen.

## DISCUSSION

The results revealed clear differences in the effects of the policy changes in the two cities. In Aberdeen, the later trading hours policy applied to 38 bars and pubs led to a statistically significant 17·4% increase in alcohol-related ambulance call-outs and 7·9% increase in reported crimes relative to Edinburgh (control city) at weekend night-times. The inconsistency of the estimated effect size for reported crimes compared to alcohol-related ambulance call-outs in Aberdeen may be due to under reporting of night-time crime incidents compared to the actual occurrences [22]. Furthermore, unlike ambulance call-outs, reported crimes do not have a specific alcohol-related flag, making it more challenging to assess their connection to alcohol intake and policy changes. Our findings in Aberdeen align with previous studies reporting an increase of alcohol harms after the implementation of extended alcohol premises hours policies [11,12]. Our findings also show a temporal shift in peak times for alcohol-related ambulance call-outs in Aberdeen, from 00:00–00:59 to 01:00–01:59, suggesting that the extended premises hours may have altered drinking patterns or increased alcohol availability, shifting the time at which individuals required ambulance services. Additionally, we found the impact of the policy change on alcohol-related ambulance call-outs in Aberdeen was significantly higher among men and individuals under 45 years of age. This is consistent with prior research showing that younger individuals, particularly men, are more likely to engage in risky drinking behaviours and are more at risk of alcohol-related ambulance call-outs [7].

In contrast, no significant association was observed due to the later trading hours policy for the 10 nightclubs in Glasgow in either alcohol-related ambulance call-outs or reported crimes when compared with our control, Edinburgh. Our Glasgow findings, of no consistent significant effect, conflict with one study that found a 1-hour extension to existing late-night licences was associated with a 34% more alcohol-related ambulance call-outs in Amsterdam, Netherlands [23]. Our findings are consistent with conclusions drawn from a systematic review (albeit based on a small number of studies), which suggested that extending alcohol premises hours by two or more hours is significantly associated with increased alcohol-related harms, while mixed results were found for extensions of less than two hours [14]. However, in separate qualitative interviews [24], frontline staff in both Glasgow and Aberdeen reported several harms from the extensions in trading hours including sometimes unmanageable pressures on police and support services due to greater numbers of vulnerable or intoxicated individuals later at night. These incidents do not feature in the crime or ambulance data we analysed in this study.

A key strength of our study is its robust design, which incorporated a structured pre-specified control selection process [19]. We triangulated our findings across modelling approaches to enhance reliability of the results. Unlike previous research that often overlooks staggered policy implementation, venue capacity, and premises type [16], our design incorporated a staggered policy covariate for Aberdeen based on the person-hours, calculated based on the number of permitted premises, their maximum capacities, and extended hours.

While the study employs robust methodological approaches, including CITS and SC methods, limitations such as potential underreporting of crimes, incidents that are not recorded in our datasets (as above), and the observational nature of the study, including potential for unmeasured time-varying confounding factors should be acknowledged. We used the approval dates rather than implementation dates, since it is unclear whether all approved premises actually implemented extended hours; qualitative findings indicate that use of the extended hours was unpredictable across both cities due to mixed demand (paper forthcoming).

For rate calculations, we used cities population as the denominator to represent those at risk, although visitors may also contribute to the night-time activities and related harms. Population data are recorded annually, therefore we used extrapolation technique. Both ARIMA and SC analyses showed similar effect direction in Aberdeen, but SC estimates were not statistically significant. This is maybe due to the frequentist (frequency based) nature of SC p-values, which are limited by number of control units in the donor pool [25]. Moreover, effect sizes are not directly comparable across models’ estimating counts versus rates, since ARIMA estimators are sensitive to scale/unit of analysis [26].

Our findings have important policy implications. This is the first UK study to find that extending trading hours for late-night licensed premises, may cause significant additional pressures on ambulance services. This ought to give pause to policymakers in both Ireland [27] and England and Wales [28,29] where further liberalisation of late-night opening hours has been proposed, especially given existing pressures on emergency services. Secondly, the findings illustrate some of the limitations of UK licensing systems [30]. If the volume and type of premises with later alcohol trading hours is an important factor dictating harms, then policymakers should have powers to control these factors. The current system in Scotland (and in England and Wales) does not distinguish between premises types [31]. Glasgow was only able to establish different standard trading hours for bars and nightclubs by including a robust definition of a nightclub in their licensing policy statement [32]. Further, local authorities cannot regulate the volume of premises eligible for later trading hours because each application has to be considered on its individual merits [33]. Although we studied the impact of the 4am extension in Glasgow’s pilot with 10 nightclubs, our findings suggest that granting the same hours to additional nightclubs would carry a further risk of negative outcomes, yet there is little to prevent multiple premises now being included. To protect public health and safety late at night, licensing systems may need to include powers to create type-specific caps on venue numbers with very late hours, for example per population or locality, similar to practice in several North American jurisdictions.

## CONCLUSION

This study demonstrates the value of robust, multicomponent analysis of outcomes when assessing the impact of changes to licensing hours. While our findings confirm the general principle that widespread extending alcohol trading hours, especially late into the night, are associated with some increased harms they also show that the intensity of these impacts is likely mediated by type and volume of outlets. It is therefore important that licensing systems can account for these factors.

## Data Availability

Time-series data on alcohol-related ambulance call-outs and reported crimes, used in this study can be accessible through formal application to the Scottish Ambulance Service and Police Scotland. Meta data used in this study have already been published online (Per capita gross disposable household income: https://www.webarchive.org.uk/wayback/archive/20180212144444/http://www.gov.scot/Topics/Statistics/Browse/Economy/QNA2017Q3; Temperature and rainfall: https://data.ceda.ac.uk/badc/ukmo-hadobs/data/insitu/MOHC/HadOBS/HadUK-Grid/v1.2.0.ceda/region; On-premises alcohol outlets: https://www.gov.scot/publications/scottish-liquor-licensing-statistics/; COVID-19 lockdown: https://spice-spotlight.scot/2023/05/10/timeline-of-coronavirus-covid-19-in-scotland/; and Public holidays: https://www.mygov.scot/scotland-bank-holidays)

## AUTHOR CONTRIBUTIONS

NF led the overall design of the mixed-methods ELEPHANT (Evaluating Later and Expanded Premises Hours for Alcohol in the Night-time Economy) study. JL led the design of the statistical analyses and supervised NS and HH in conducting the analyses. JL, NF, CA, IHC, EM, HH, and NS contributed to the conceptualisation and methodology. NS was responsible for data curation, formal analysis, drafting the original manuscript. NS, JL, and FM contributed to validation of data curation and analysis. NS and CA were responsible for data visualisation. NF, EM, JN, ES, CE, RO, GM, IHC, MC, KM, AM, and IU contributed to interpretation. NF, JL, EM, CA, CE, ES, and AM are lead (NF) and co-applicants on the NIHR grant that funded ELEPHANT. All authors critically reviewed and edited subsequent versions of the manuscript and approved the final version of the manuscript for publication.

## ACKNOWLEDGEMENTS

We want to thanks to the Scottish Ambulance Service and Police Scotland.

## PATIENT AND PUBLIC INVOLVEMENT

Patients and/or the public were involved in the design, or conduct, or reporting, or dissemination plans of this research.

## Appendix 1: Control selection process and study design

**Figure A1:**
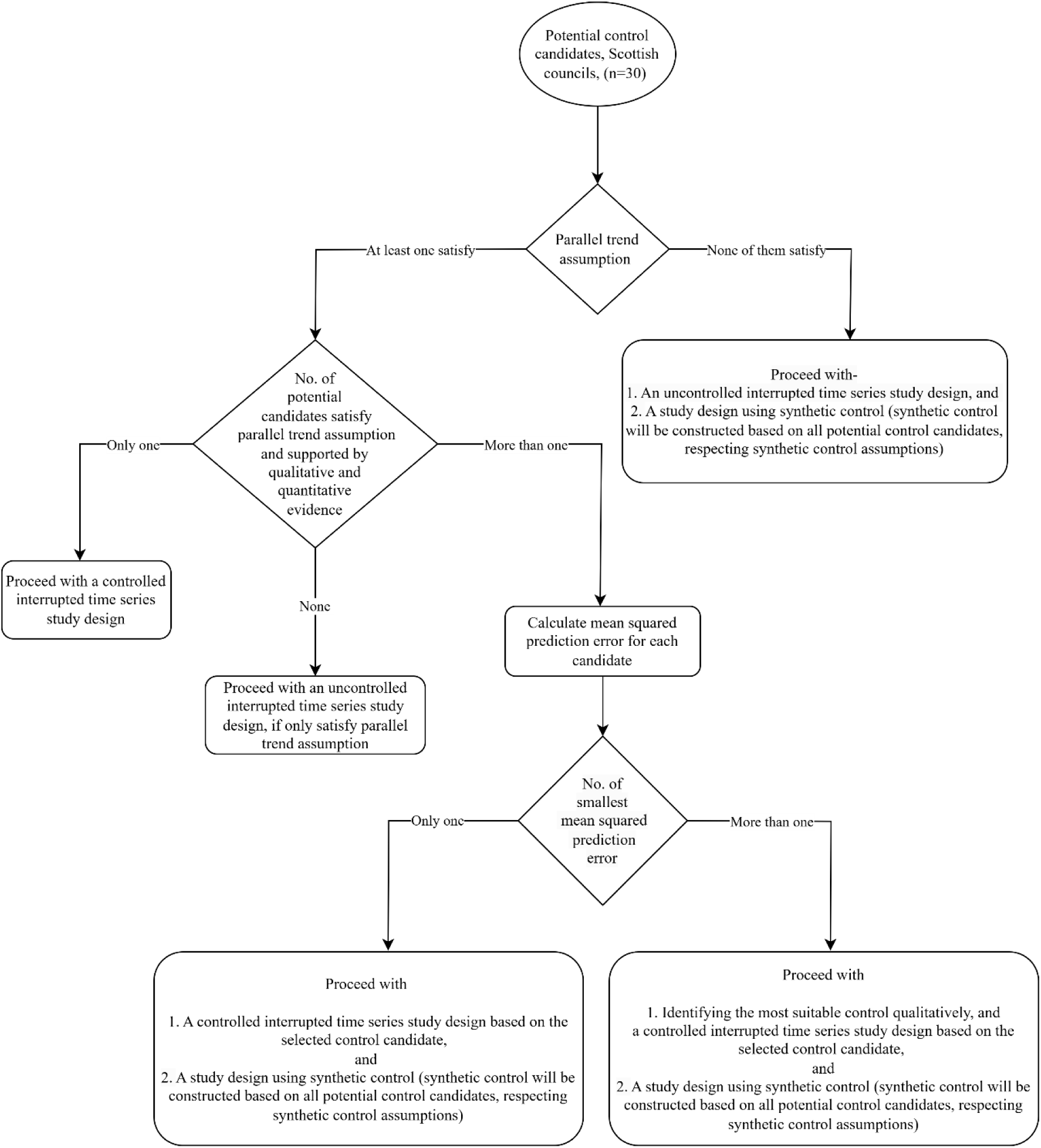
Flow diagram for the control selection procedure.

### Appendix 1.1. Alcohol-related ambulance call-outs

In step 1, we tested the parallel trend assumption both graphically and statistically for 30 potential Scottish cities/council areas, excluding Aberdeen and Glasgow. We estimated the slope coefficients for Aberdeen and Glasgow against each of the 30 potential control candidates, using data from the pre-intervention periods only. Any control candidate with a statistically significant slope coefficient, indicating a statistically significant difference in trends from Aberdeen or Glasgow, was excluded. As a result, Stirling was excluded as a control for Aberdeen, while Angus, Argyll and Bute, Dundee, East Ayrshire, Highland, Perth and Kinross, Scottish Borders, and Stirling were excluded as controls for Glasgow.

In step 2, we reviewed the licensing policies of the remaining potential control cities/councils and excluded those with similar extended premises hours policies to Aberdeen or Glasgow. Additionally, we removed candidates whose population-adjusted density of on-premises alcohol outlets was more than one standard deviation above or below the densities of the intervention areas (Aberdeen and Glasgow).

In step 3, we computed the mean squared prediction errors (MSPEs) for all remaining control candidates. The MSPE represents the mean squared differences between the intervention areas (Aberdeen/Glasgow) and the potential control candidates during the pre-intervention periods. For Aberdeen, the candidates with the lowest MSPEs were Edinburgh, Fife, and North Lanarkshire, while for Glasgow, Edinburgh had the lowest MSPE. Ultimately, Edinburgh was chosen as the control for both Aberdeen and Glasgow. For Aberdeen, this decision was informed by Edinburgh’s significant contribution in synthetic control analysis compared to Fife and North Lanarkshire, alongside our a priori decision to use Edinburgh as a control.

#### Step 1: Testing parallel trend assumptions graphically and statistically

| **Parallel trend assumption test: Ambulance call-outs (based on rate)** |  |  |  |  |
| --- | --- | --- | --- | --- |
| **Council name** | **Aberdeen** |  | **Glasgow** |  |
|  | **Slope** | **P-value** | **Slope** | **P-value** |
| Aberdeenshire | -0.0009 | 0.268 | -0.0001 | 0.763 |
| Angus | -0.0002 | 0.826 | -0.0006 | 0.049 |
| Argyll and Bute | -0.0022 | 0.054 | -0.0016 | <0.001 |
| City of Edinburgh | -0.0017 | 0.061 | 0.0003 | 0.248 |
| Clackmannanshire | -0.0009 | 0.554 | -0.0006 | 0.240 |
| Dumfries and Galloway | -0.0007 | 0.468 | -0.0004 | 0.191 |
| Dundee City | -0.0007 | 0.514 | -0.0009 | 0.010 |
| East Ayrshire | -0.0014 | 0.169 | -0.0009 | 0.010 |
| East Dunbartonshire | -0.0011 | 0.258 | 0.0004 | 0.097 |
| East Lothian | 0.0003 | 0.760 | 0.0000 | 0.913 |
| East Renfrewshire | -0.0019 | 0.050 | -0.0002 | 0.384 |
| Falkirk | 0.0003 | 0.803 | -0.0003 | 0.319 |
| Fife | -0.0007 | 0.389 | -0.0003 | 0.186 |
| Highland | 0.0006 | 0.469 | -0.0006 | 0.007 |
| Inverclyde | 0.0013 | 0.321 | 0.0005 | 0.219 |
| Midlothian | -0.0005 | 0.639 | -0.0001 | 0.847 |
| Moray | -0.0006 | 0.612 | -0.0007 | 0.055 |
| North Ayrshire | 0.0004 | 0.757 | 0.0004 | 0.221 |
| North Lanarkshire | 0.0000 | 0.989 | 0.0003 | 0.286 |
| Orkney Islands | 0.0003 | 0.825 | 0.0005 | 0.307 |
| Perth and Kinross | -0.0014 | 0.169 | 0.0007 | 0.009 |
| Renfrewshire | 0.0001 | 0.925 | 0.0003 | 0.325 |
| Scottish Borders | -0.0018 | 0.095 | 0.0012 | 0.001 |
| Shetland Islands | -0.0008 | 0.587 | -0.0002 | 0.718 |
| South Ayrshire | -0.0005 | 0.688 | -0.0004 | 0.186 |
| South Lanarkshire | -0.0013 | 0.195 | -0.0002 | 0.536 |
| Stirling | -0.0026 | 0.019 | -0.0008 | 0.017 |
| West Dunbartonshire | 0.0014 | 0.246 | -0.0003 | 0.477 |
| West Lothian | 0.0001 | 0.912 | -0.0001 | 0.620 |
| Western Isles - Eilean Siar | -0.0001 | 0.945 | -0.0008 | 0.090 |

#### Step 2: Assess qualitatively and quantitatively the councils/cities that passed parallel test assumption at Step 1

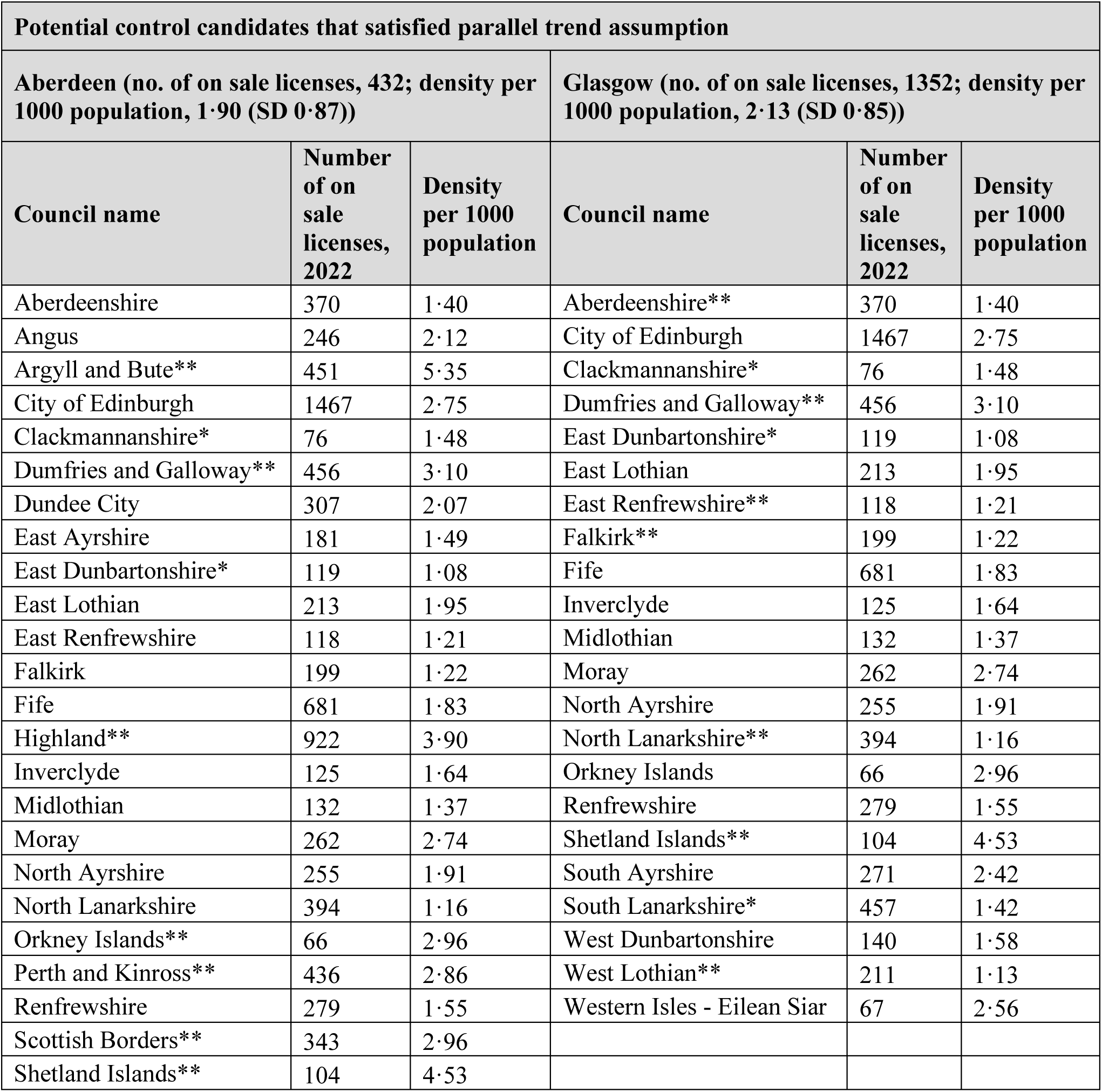
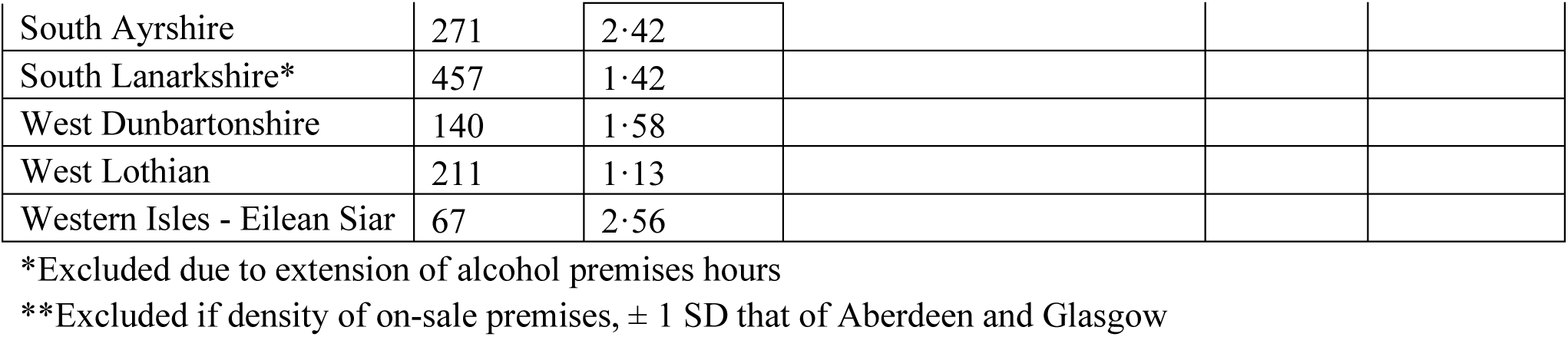

| Potential control candidates that satisfied parallel trend assumption |  |  |  |  |  |
| --- | --- | --- | --- | --- | --- |
| Aberdeen (no. of on sale licenses, 432; density per 1000 population, 1.90 (SD 0.87)) |  |  | Glasgow (no. of on sale licenses, 1352; density per 1000 population, 2.13 (SD 0.85)) |  |  |
| Council name | Number of on sale licenses, 2022 | Density per 1000 population | Council name | Number of on sale licenses, 2022 | Density per 1000 population |
| Aberdeenshire | 370 | 1.40 | Aberdeenshire** | 370 | 1.40 |
| Angus | 246 | 2.12 | City of Edinburgh | 1467 | 2.75 |
| Argyll and Bute** | 451 | 5.35 | Clackmannanshire* | 76 | 1.48 |
| City of Edinburgh | 1467 | 2.75 | Dumfries and Galloway** | 456 | 3.10 |
| Clackmannanshire* | 76 | 1.48 | East Dunbartonshire* | 119 | 1.08 |
| Dumfries and Galloway** | 456 | 3.10 | East Lothian | 213 | 1.95 |
| Dundee City | 307 | 2.07 | East Renfrewshire** | 118 | 1.21 |
| East Ayrshire | 181 | 1.49 | Falkirk** | 199 | 1.22 |
| East Dunbartonshire* | 119 | 1.08 | Fife | 681 | 1.83 |
| East Lothian | 213 | 1.95 | Inverclyde | 125 | 1.64 |
| East Renfrewshire | 118 | 1.21 | Midlothian | 132 | 1.37 |
| Falkirk | 199 | 1.22 | Moray | 262 | 2.74 |
| Fife | 681 | 1.83 | North Ayrshire | 255 | 1.91 |
| Highland** | 922 | 3.90 | North Lanarkshire** | 394 | 1.16 |
| Inverclyde | 125 | 1.64 | Orkney Islands | 66 | 2.96 |
| Midlothian | 132 | 1.37 | Renfrewshire | 279 | 1.55 |
| Moray | 262 | 2.74 | Shetland Islands** | 104 | 4.53 |
| North Ayrshire | 255 | 1.91 | South Ayrshire | 271 | 2.42 |
| North Lanarkshire | 394 | 1.16 | South Lanarkshire* | 457 | 1.42 |
| Orkney Islands** | 66 | 2.96 | West Dunbartonshire | 140 | 1.58 |
| Perth and Kinross** | 436 | 2.86 | West Lothian** | 211 | 1.13 |
| Renfrewshire | 279 | 1.55 | Western Isles - Eilean Siar | 67 | 2.56 |
| Scottish Borders** | 343 | 2.96 |  |  |  |
| Shetland Islands** | 104 | 4.53 |  |  |  |

|  |  |  |
| --- | --- | --- |
| South Ayrshire | 271 | 2.42 |
| South Lanarkshire* | 457 | 1.42 |
| West Dunbartonshire | 140 | 1.58 |
| West Lothian | 211 | 1.13 |
| Western Isles - Eilean Siar | 67 | 2.56 |

\*Excluded due to extension of alcohol premises hours
\*\*Excluded if density of on-sale premises, $\pm 1$ SD that of Aberdeen and Glasgow

#### Step 3: Calculate mean square prediction error based on pre-intervention data to choose control candidate based on lowest value

| Potential control candidates satisfied parallel trend assumption and passed qualitative and quantitative assessment |  |  |  |
| --- | --- | --- | --- |
| Intervention: Aberdeen |  | Intervention: Glasgow |  |
| Council name | Mean squared prediction error (MSPE) | Council name | Mean squared prediction error (MSPE) |
| Aberdeenshire | 0.23 | City of Edinburgh | 0.07 |
| Angus | 0.14 | East Lothian | 0.29 |
| City of Edinburgh | 0.07 | Fife | 0.13 |
| Dundee City | 0.10 | Inverclyde | 0.16 |
| East Ayrshire | 0.11 | Midlothian | 0.20 |
| East Lothian | 0.16 | Moray | 0.24 |
| East Renfrewshire | 0.28 | North Ayrshire | 0.11 |
| Falkirk | 0.11 | Orkney Islands | 0.63 |
| Fife | 0.07 | Renfrewshire | 0.13 |
| Inverclyde | 0.14 | South Ayrshire | 0.16 |
| Midlothian | 0.12 | West Dunbartonshire | 0.16 |
| Moray | 0.16 | Western Isles - Eilean Siar | 0.45 |
| North Ayrshire | 0.12 |  |  |
| North Lanarkshire | 0.07 |  |  |
| Renfrewshire | 0.08 |  |  |
| South Ayrshire | 0.13 |  |  |
| West Dunbartonshire | 0.12 |  |  |
| West Lothian | 0.12 |  |  |
| Western Isles - Eilean Siar | 0.31 |  |  |

### Appendix 1.2. Reported crimes

In step 1, for reported crimes, Shetland Islands was excluded as a control for Aberdeen, while Aberdeenshire, East Dunbartonshire, East Lothian, East Renfrewshire, Highland, North Lanarkshire, South Ayrshire, and South Lanarkshire were excluded as controls for Glasgow.

In step 2, Argyll and Bute, Clackmannanshire, Dumfries and Galloway, East Dunbartonshire, Highland, Orkney Islands, Perth and Kinross, Scottish Borders, South Lanarkshire, and Stirling excluded as controls for Aberdeen. Similarly, for Glasgow, Argyll and Bute, Clackmannanshire, Dumfries and Galloway, Falkirk, Orkney Islands, Perth and Kinross, Scottish Borders, Shetland Islands, and West Lothian were excluded.

In step 3, Edinburgh was chosen as the control for both Aberdeen and Glasgow for reported crimes based on the lowest MSPEs among the surviving control candidates.

#### Step 1: Testing parallel trend assumptions graphically and statistically

| **Parallel trend assumption test: Police call-outs (based on rate)** |  |  |  |  |
| --- | --- | --- | --- | --- |
| **Council name** | **Aberdeen** |  | **Glasgow** |  |
|  | **Slope** | **P-value** | **Slope** | **P-value** |
| Aberdeenshire | 0.0000 | 0.973 | -0.0008 | 0.012 |
| Angus | -0.0015 | 0.289 | -0.0003 | 0.495 |
| Argyll and Bute | -0.0003 | 0.866 | -0.0009 | 0.078 |
| City of Edinburgh | 0.0006 | 0.558 | -0.0002 | 0.538 |
| Clackmannanshire | 0.0007 | 0.665 | -0.0002 | 0.695 |
| Dumfries and Galloway | 0.0004 | 0.757 | -0.0006 | 0.163 |
| Dundee City | -0.0002 | 0.899 | 0.0001 | 0.772 |
| East Ayrshire | -0.0019 | 0.168 | 0.0001 | 0.851 |
| East Dunbartonshire | -0.0008 | 0.514 | -0.0011 | 0.001 |
| East Lothian | -0.0001 | 0.924 | -0.0011 | 0.007 |
| East Renfrewshire | -0.0010 | 0.461 | -0.0007 | 0.034 |
| Falkirk | 0.0012 | 0.414 | -0.0007 | 0.061 |
| Fife | 0.0005 | 0.643 | -0.0002 | 0.505 |
| Highland | 0.0024 | 0.081 | 0.0007 | 0.042 |
| Inverclyde | -0.0002 | 0.911 | -0.0009 | 0.070 |
| Midlothian | 0.0011 | 0.492 | -0.0005 | 0.210 |
| Moray | 0.0003 | 0.837 | -0.0005 | 0.305 |
| North Ayrshire | 0.0008 | 0.512 | 0.0003 | 0.428 |
| North Lanarkshire | -0.0006 | 0.562 | -0.0009 | 0.001 |
| Orkney Islands | 0.0024 | 0.369 | -0.0007 | 0.349 |
| Perth and Kinross | 0.0002 | 0.878 | -0.0005 | 0.118 |
| Renfrewshire | 0.0002 | 0.834 | -0.0007 | 0.079 |
| Scottish Borders | 0.0004 | 0.758 | -0.0006 | 0.075 |
| Shetland Islands | 0.0067 | 0.009 | 0.0012 | 0.101 |
| South Ayrshire | 0.0009 | 0.569 | -0.0008 | 0.035 |
| South Lanarkshire | 0.0000 | 0.971 | -0.0008 | 0.005 |
| Stirling | 0.0012 | 0.433 | -0.0001 | 0.809 |
| West Dunbartonshire | 0.0022 | 0.207 | -0.0002 | 0.687 |
| West Lothian | 0.0002 | 0.868 | -0.0005 | 0.106 |
| Western Isles - Eilean Siar | 0.0036 | 0.063 | -0.0006 | 0.325 |

#### Step 2: Assess qualitatively and quantitatively to the councils/cities passed parallel test assumption

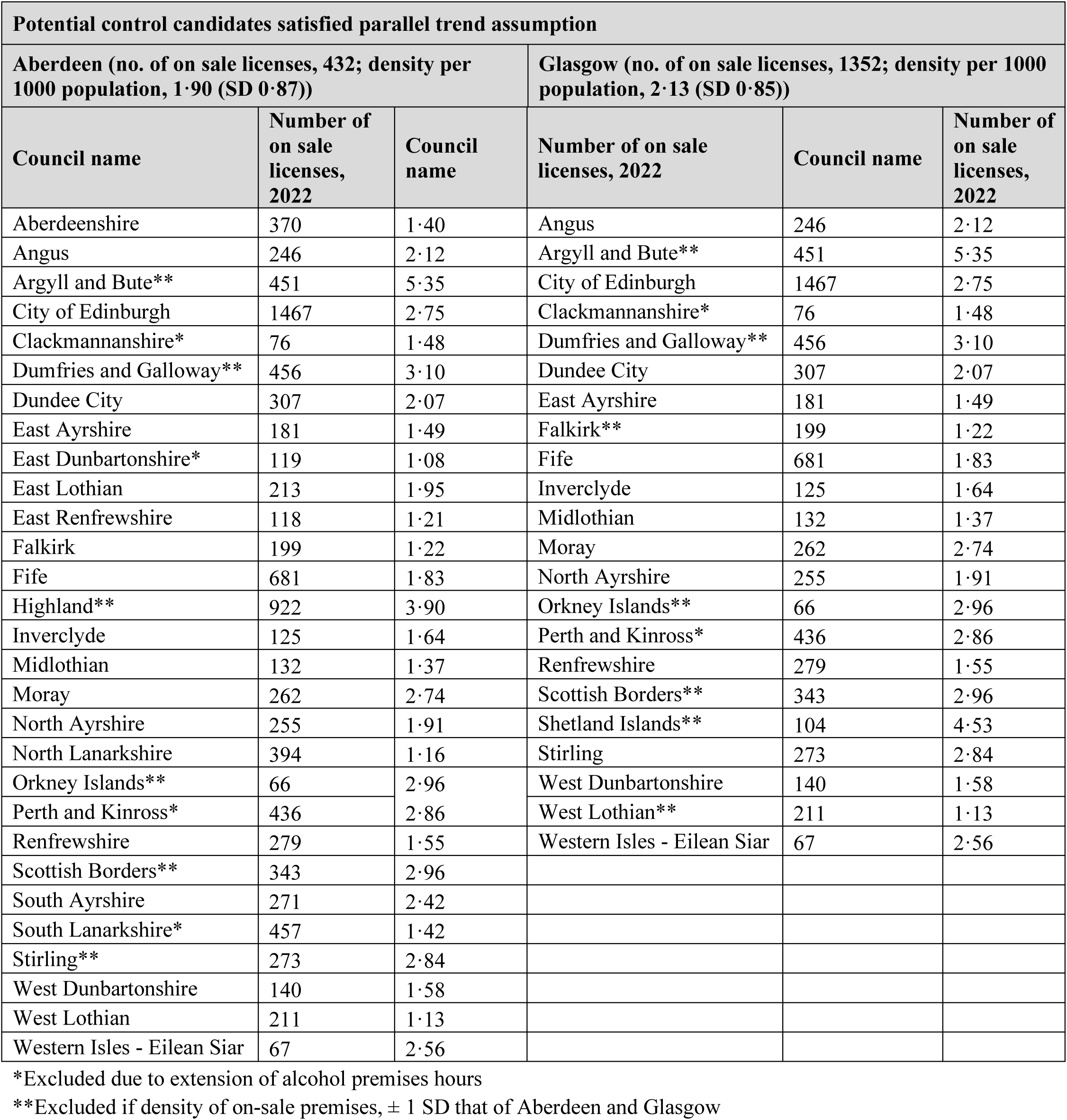

#### Step 3: Calculate mean square prediction error based on pre-intervention data to choose control candidate based on lowest value

**Potential control candidates satisfied parallel trend assumption and passed qualitative and quantitative assessment**
| **Council name** | **Mean squared prediction error (MSPE)** | **Council name** | **Mean squared prediction error (MSPE)** |
| --- | --- | --- | --- |
| Aberdeenshire | 0.26 | Angus | 0.23 |
| Angus | 0.19 | City of Edinburgh | 0.09 |
| City of Edinburgh | 0.09 | Dundee City | 0.12 |
| Dundee City | 0.15 | East Ayrshire | 0.25 |
| East Ayrshire | 0.19 | Fife | 0.16 |
| East Lothian | 0.37 | Inverclyde | 0.34 |
| East Renfrewshire | 0.50 | Midlothian | 0.28 |
| Falkirk | 0.18 | Moray | 0.24 |
| Fife | 0.13 | North Ayrshire | 0.19 |
| Inverclyde | 0.31 | Orkney Islands | 0.63 |
| Midlothian | 0.26 | Renfrewshire | 0.22 |
| Moray | 0.20 | Scottish Borders | 0.36 |
| North Ayrshire | 0.15 | Stirling | 0.32 |
| North Lanarkshire | 0.15 | West Dunbartonshire | 0.23 |
| Renfrewshire | 0.16 | Western Isles - Eilean Siar | 0.67 |
| South Ayrshire | 0.26 |  |  |
| West Dunbartonshire | 0.27 |  |  |
| West Lothian | 0.13 |  |  |
| Western Isles - Eilean Siar | 0.57 |  |  |

## Appendix 2: Analysis

### Appendix 2.1. Descriptive analysis

**Table A1.**
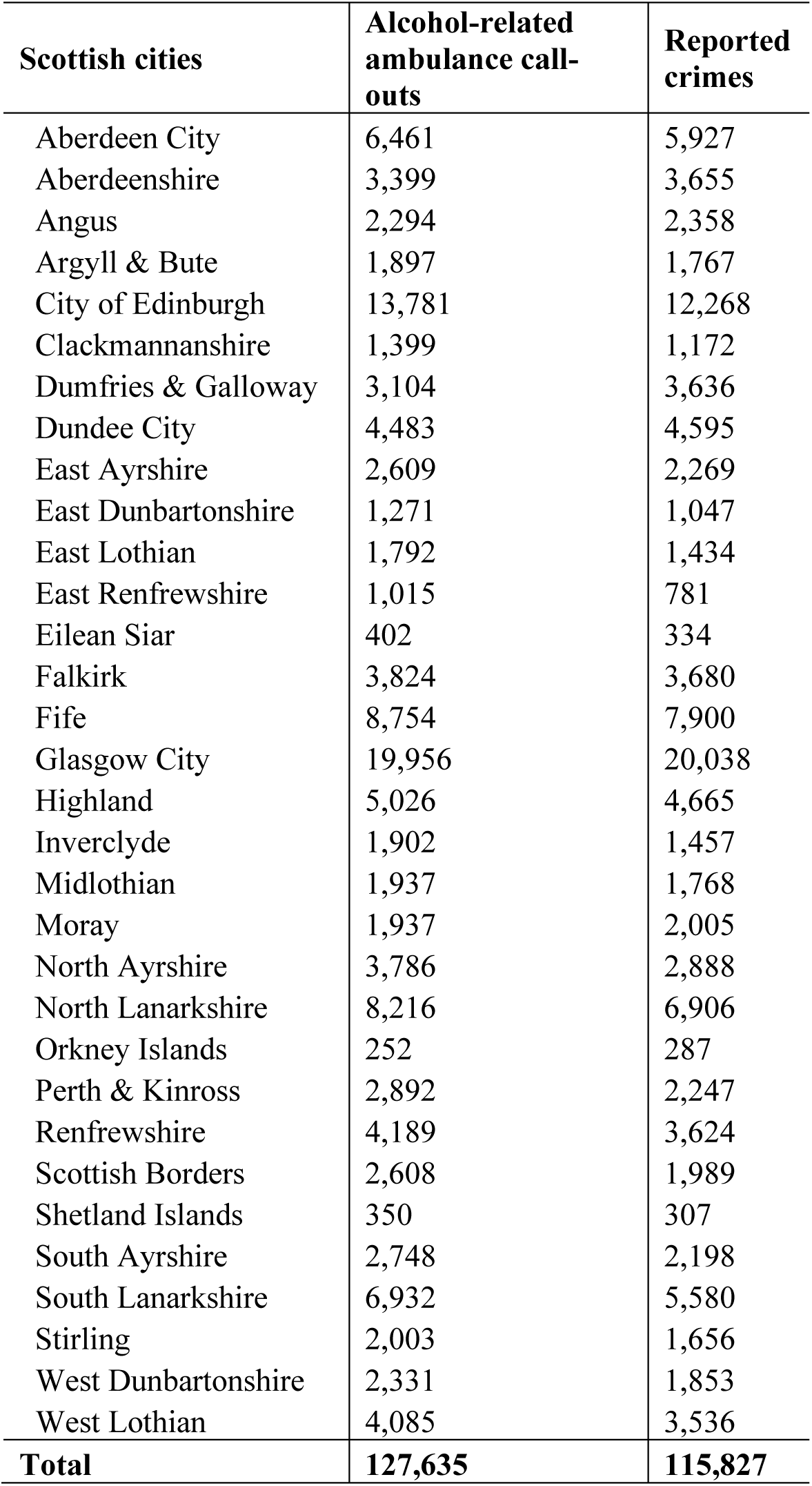
Total number of weekend night-time alcohol-related ambulance call-outs and reported crimes by Scottish cities from 1 May 2015 to 31 July 2022.

| Scottish cities | Alcohol-related ambulance call-outs | Reported crimes |
| --- | --- | --- |
| Aberdeen City | 6,461 | 5,927 |
| Aberdeenshire | 3,399 | 3,655 |
| Angus | 2,294 | 2,358 |
| Argyll & Bute | 1,897 | 1,767 |
| City of Edinburgh | 13,781 | 12,268 |
| Clackmannanshire | 1,399 | 1,172 |
| Dumfries & Galloway | 3,104 | 3,636 |
| Dundee City | 4,483 | 4,595 |
| East Ayrshire | 2,609 | 2,269 |
| East Dunbartonshire | 1,271 | 1,047 |
| East Lothian | 1,792 | 1,434 |
| East Renfrewshire | 1,015 | 781 |
| Eilean Siar | 402 | 334 |
| Falkirk | 3,824 | 3,680 |
| Fife | 8,754 | 7,900 |
| Glasgow City | 19,956 | 20,038 |
| Highland | 5,026 | 4,665 |
| Inverclyde | 1,902 | 1,457 |
| Midlothian | 1,937 | 1,768 |
| Moray | 1,937 | 2,005 |
| North Ayrshire | 3,786 | 2,888 |
| North Lanarkshire | 8,216 | 6,906 |
| Orkney Islands | 252 | 287 |
| Perth & Kinross | 2,892 | 2,247 |
| Renfrewshire | 4,189 | 3,624 |
| Scottish Borders | 2,608 | 1,989 |
| Shetland Islands | 350 | 307 |
| South Ayrshire | 2,748 | 2,198 |
| South Lanarkshire | 6,932 | 5,580 |
| Stirling | 2,003 | 1,656 |
| West Dunbartonshire | 2,331 | 1,853 |
| West Lothian | 4,085 | 3,536 |
| **Total** | **127,635** | **115,827** |

**Table A2.** Age, sex and socio-economic deprivation* distribution of patients in alcohol-related ambulance call-outs at the weekend night-time.

| Patient characteristics | Aberdeen,<br>n(%) | Glasgow,<br>n(%) | Edinburgh,<br>n(%) |
| --- | --- | --- | --- |
| **Age group (years)** |  |  |  |
| < 25 | 1,786 (27·64) | 4,609 (23·1) | 3,954 (28·69) |
| 25-34 | 1,136 (17·58) | 3,531 (17·69) | 2,366 (17·17) |
| 35-44 | 849 (13·14) | 2,827 (14·17) | 1,671 (12·13) |
| 45+ | 2,190 (33·9) | 7,588 (38·02) | 4,828 (35·03) |
| Unknown | 500 (7·74) | 1,401 (7·02) | 962 (6·98) |
| **Gender** |  |  |  |
| Female | 2,649 (41·00) | 7,300 (36·58) | 5,655 (41·03) |
| Male | 3,576 (55·35) | 11,892 (59·59) | 7,663 (55·61) |
| Others | - | 3 (0·02) | 4 (0·03) |
| Unknown | 236 (3·65) | 761 (3·81) | 459 (3·33) |
| **Socio-economic deprivation** |  |  |  |
| 1 (most deprived) | 111 (1·72) | 7,385 (37·01) | 1,189 (8·63) |
| 2 | 580 (8·98) | 2,868 (14·37) | 1,311 (9·51) |
| 3 | 1,012 (15·66) | 1,492 (7·48) | 1,141 (8·28) |
| 4 | 600 (9·29) | 1,278 (6·40) | 1,852 (13·44) |
| 5 | 1,387 (21·47) | 1,342 (6·72) | 1,826 (13·25) |
| 6 | 624 (9·66) | 2,447 (12·26) | 1,467 (10·65) |
| 7 | 627 (9·70) | 601 (3·01) | 914 (6·63) |
| 8 | 431 (6·67) | 1,655 (8·29) | 1,422 (10·32) |
| 9 | 423 (6·55) | 619 (3·1) | 795 (5·77) |
| 10 (less deprived) | 666 (10·31) | 269 (1·35) | 1,864 (13·53) |
| **Total** | **6,461** | **19,956** | **13,781** |

\*Socio-economic deprivation score is based on the postcode of the incident location

**Table A3.**
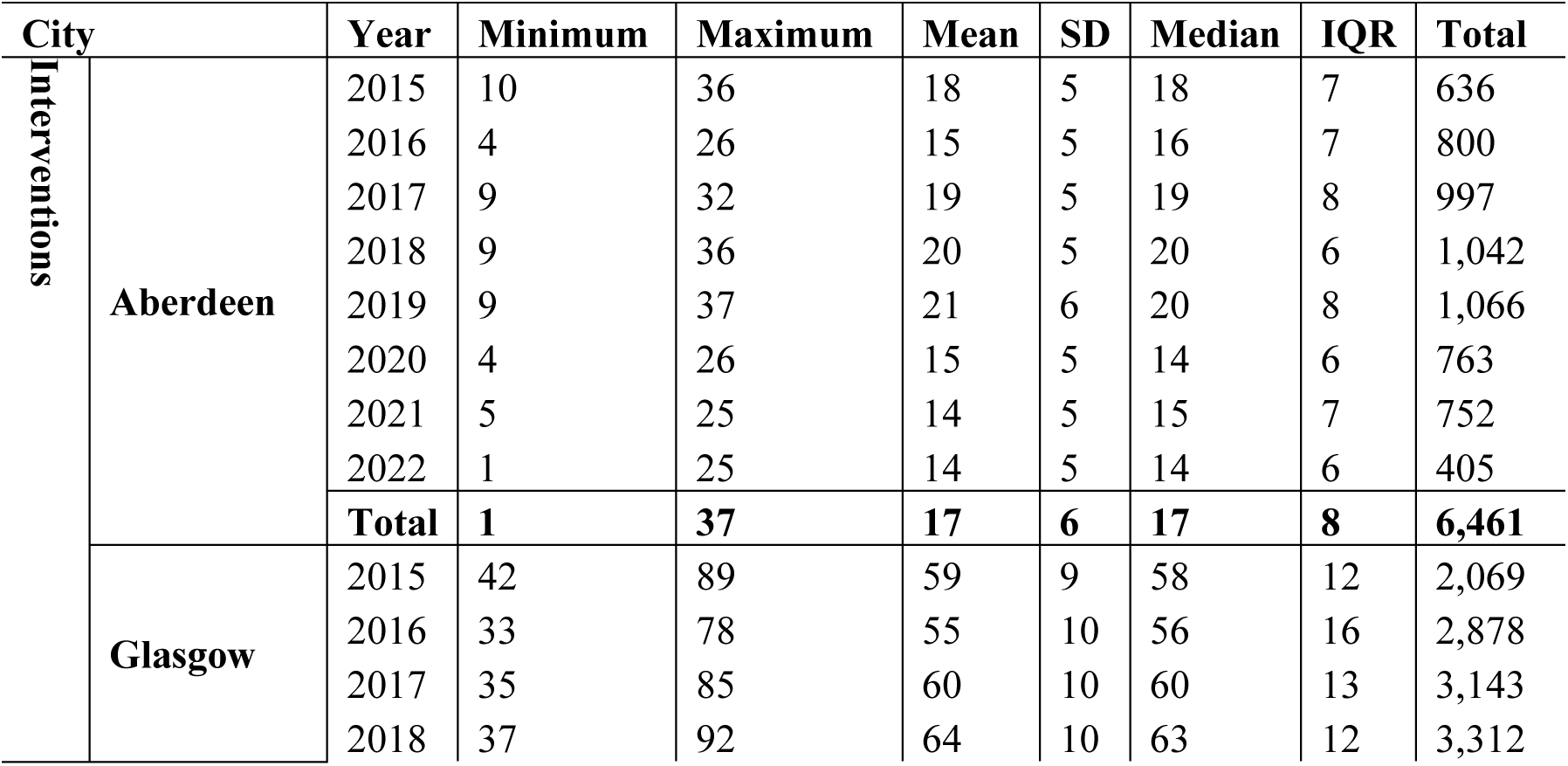
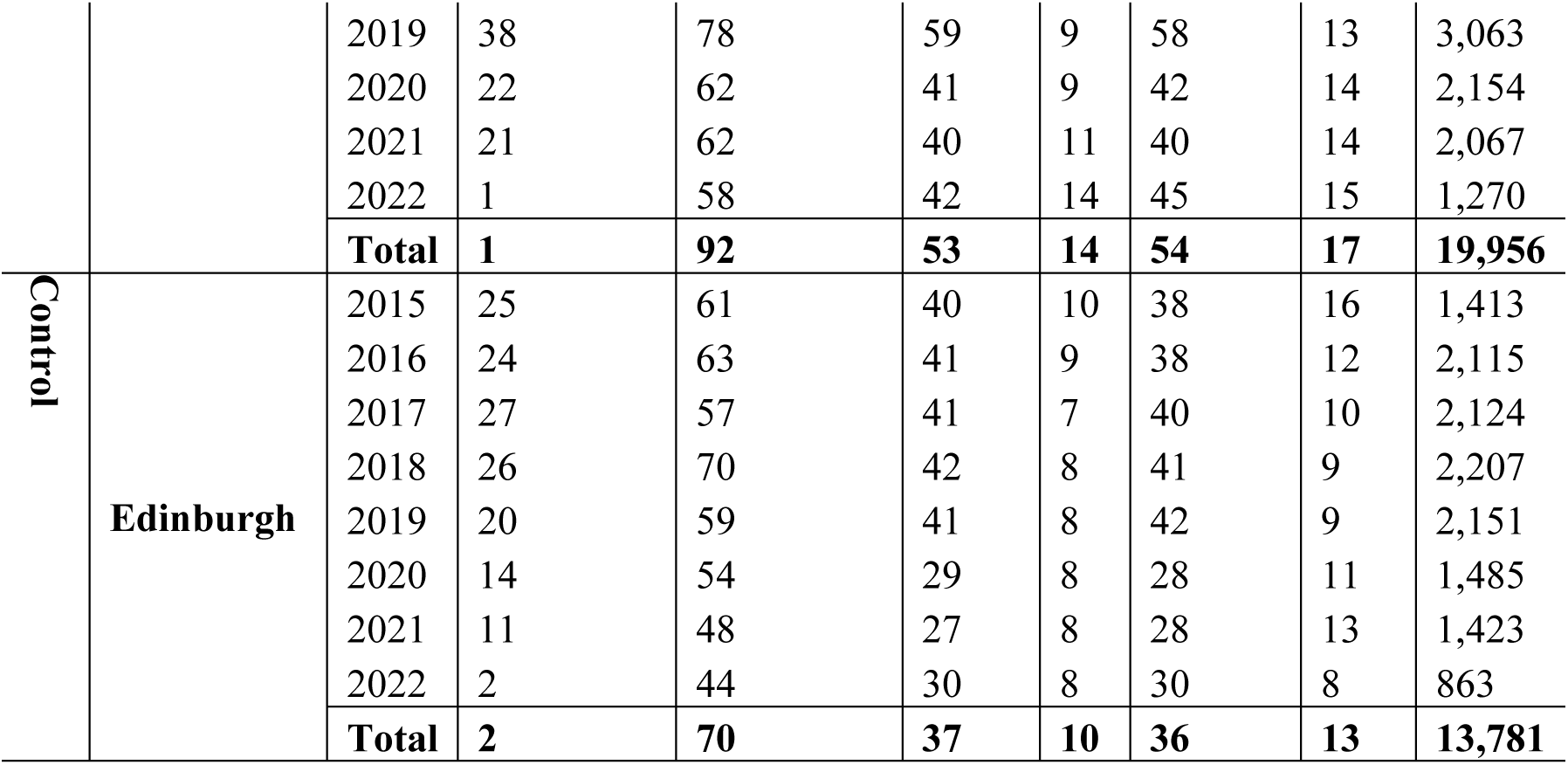
Summary of weekend night-time alcohol-related ambulance call-outs by cities.

**Table A4.** Total number of weekend night-time reported crimes according to crime types from 1 May 2015 to 31 July 2022.

| Type of reported crimes | Aberdeen,<br>n(%) | Glasgow,<br>n(%) | Edinburgh,<br>n(%) |
| --- | --- | --- | --- |
| Common Assault | 4,172 (70·39) | 13,702 (68·38) | 9,493 (77·38) |
| Common assault of an emergency worker | 765 (12·91) | 3,091 (15·43) | 1,135 (9·25) |
| Consuming outwith permitted hours | - | - | 4 (0·03) |
| Culpable and reckless, causing injury | 3 (0·05) | 13 (0·06) | 9 (0·07) |
| Disorderly on licensed premises | 104 (1·75) | 200 (1) | 60 (0·49) |
| Drunk and Incapable | 5 (0·08) | 294 (1·47) | 99 (0·81) |
| Drunk and attempting to enter licensed premises | 22 (0·37) | 24 (0·12) | 5 (0·04) |
| Drunk in charge of a child | 5 (0·08) | 24 (0·12) | 6 (0·05) |
| Drunk in or attempting to enter designated sports ground | 1 (0·02) | 2 (0·01) | - |
| Licensed person, employee or agent drunk in licensed premise | 1 (0·02) | 3 (0·01) | 6 (0·05) |
| Licensed persons, other offences | 2 (0·03) | 9 (0·04) | 5 (0·04) |
| Liquor licensing laws, other offences | - | 8 (0·04) | - |
| Person under 18 buying excisable liquor in bar | - | - | 2 (0·02) |
| Permitting riotous behaviour in licensed premise | - | 7 (0·03) | - |
| Purchasing excise liquor for consumption by person under 18 | 1 (0·02) | - | - |
| Refusing to quit a licensed premise | 207 (3·49) | 335 (1·67) | 205 (1·67) |
| Robbery | 131 (2·21) | 408 (2·04) | 306 (2·49) |
| Sale of drink to person under 18 | - | 7 (0·03) | - |
| Serious Assault | 474 (8) | 1,841 (9·19) | 858 (6·99) |
| Sports grounds offences possessing alcohol | 1 (0·02) | 9 (0·04) | 2 (0·02) |
| Threats and Extortion | 33 (0·56) | 61 (0·3) | 73 (0·6) |
| **Total reported crimes** | **5,927** | **20,038** | **12,268** |

**Table A5.**
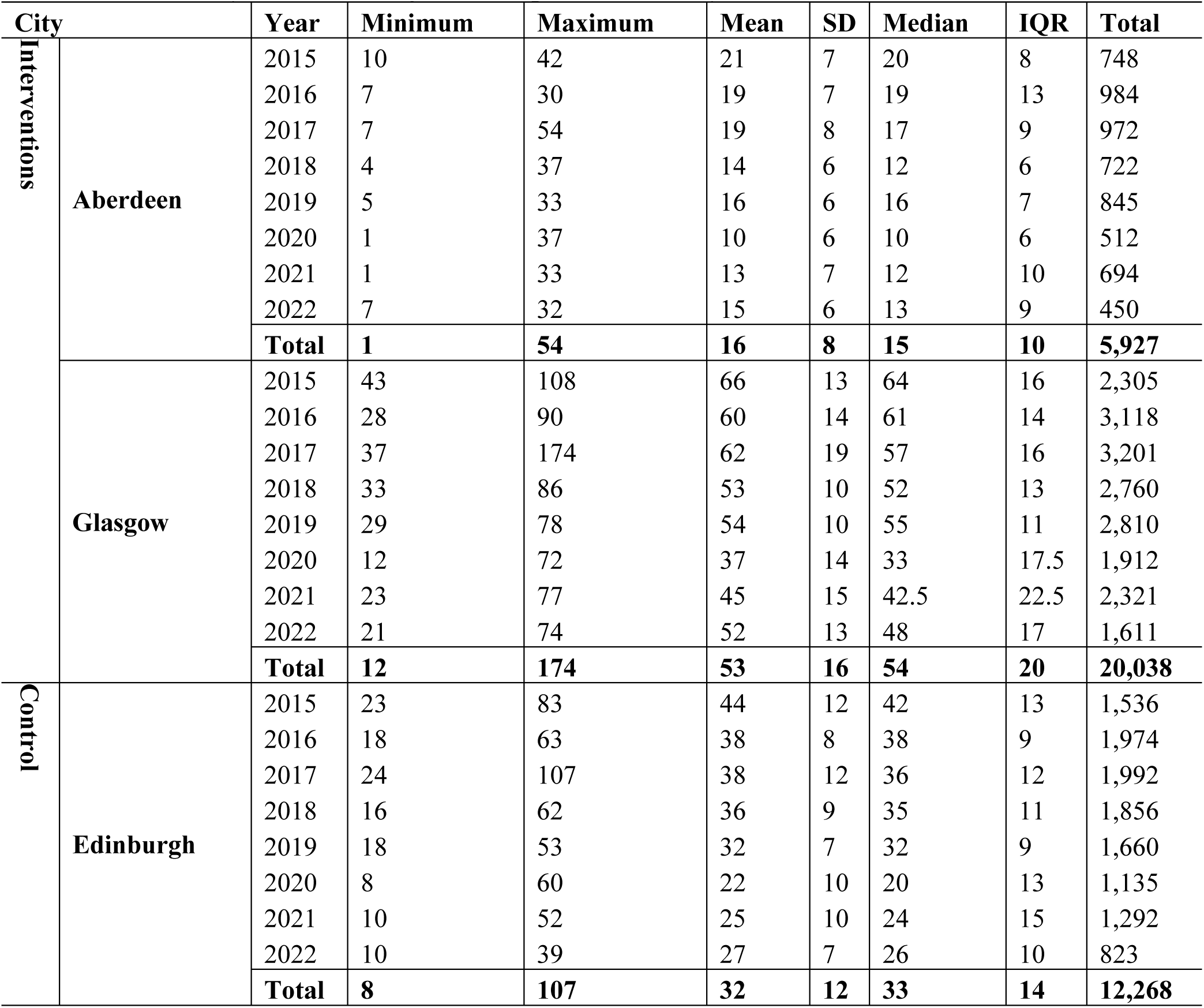
Summary of weekend night-time reported crimes by cities.

**Figure A2.**
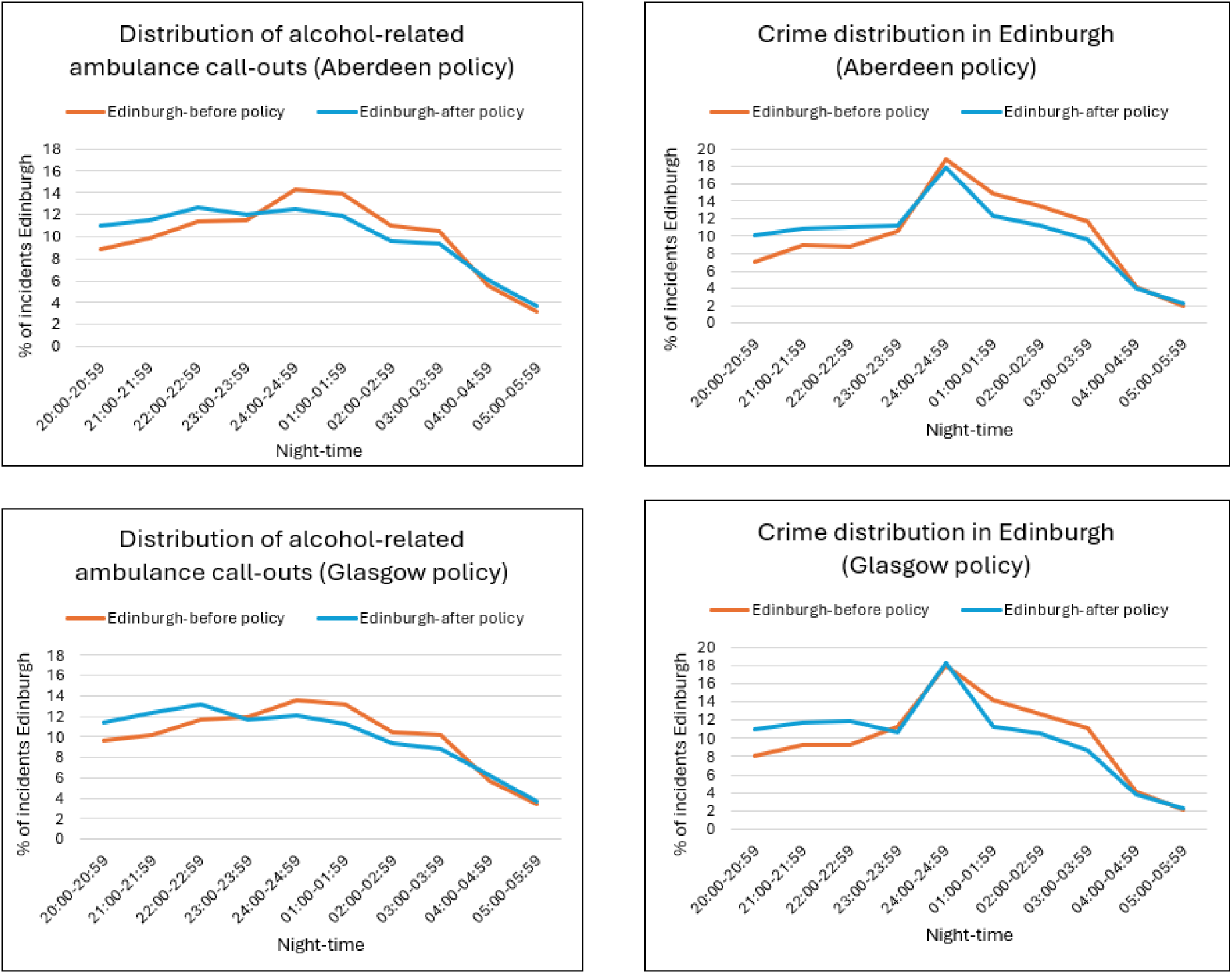
Distribution of weekend night-time alcohol-related ambulance call-out and reported crimes in Edinburgh.

**Figure A3:**
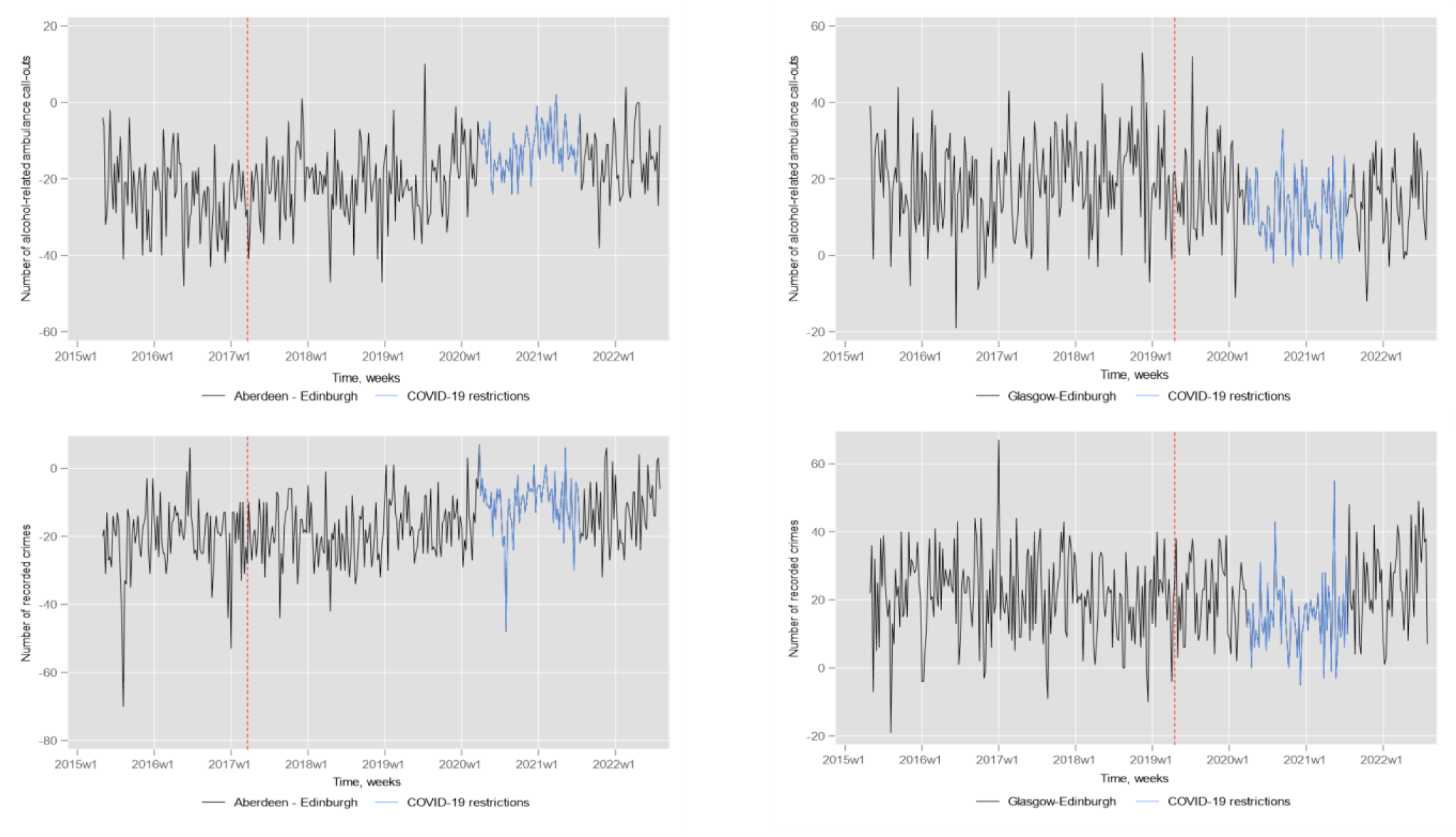
Differences (intervention city minus (-) control city) of number of weekend night-time alcohol-related ambulance call-outs and reported crimes over-time between intervention and control cities.

### Appendix 2.2. Description of confounding variables

We included several confounding variables in our model, such as per capita gross disposable household income, weather conditions (mean temperature and rainfall), and the total number of on-premises alcohol outlets. These variables were incorporated as differences between the intervention and control areas (e.g., number of on-premises outlets in Aberdeen minus the number in Edinburgh). Additionally, we adjusted for dummy variables representing COVID-19 restrictions, public holidays, and outliers. These dummy variables were binary, taking the value “1” to indicate the presence of a restriction, holiday, or outlier, and “0” to indicate their absence. Public holiday data were extracted separately for each Scottish city. Outliers were defined as data points that fell more than 1.5 times the interquartile range above the upper quartile or below the lower quartile. To account for COVID-19 restrictions, we used city-specific lockdown policies based on the type of premises, as issued by the Scottish Government. While restrictions on bars, clubs, and nightclubs were implemented simultaneously across Scotland on 21st March 2020, those affecting nightclubs remained in place for a longer period compared to bars and pubs. We also included a time trend variable in the model to account for temporal effects, if it was statistically significant.

#### Appendix 2.2.1. Data sources for confounding variables

1. Per capita gross disposable household income [34]
2. Weather conditions (mean temperature, rainfall) [35]
3. Number of on-premises alcohol outlets [36]
4. COVID-19 lockdown (imposing closure or restricted hours for alcohol-premises) [37]
5. Public holidays [38]

### Appendix 2.3. Secondary and Sub-group analysis

**Table A6.**
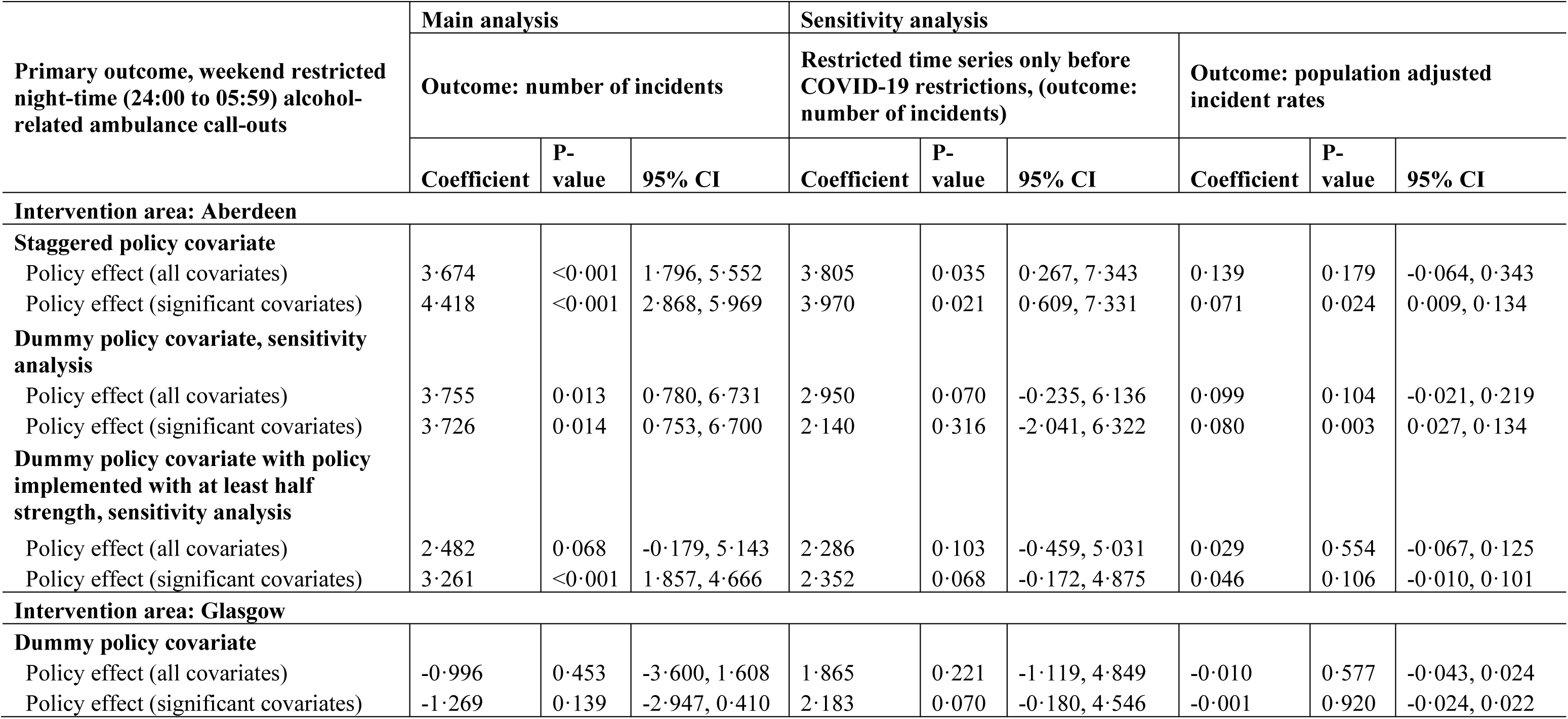
Effect of policy changes on weekend restricted night-time (24:00 to 05:59) alcohol-related ambulance call-outs in Aberdeen and Glasgow.

| Primary outcome, weekend restricted night-time (24:00 to 05:59) alcohol-related ambulance call-outs | Main analysis |  |  | Sensitivity analysis |  |  |  |  |  |
| --- | --- | --- | --- | --- | --- | --- | --- | --- | --- |
|  | Outcome: number of incidents |  |  | Restricted time series only before COVID-19 restrictions, (outcome: number of incidents) |  |  | Outcome: population adjusted incident rates |  |  |
|  | Coefficient | P-value | 95% CI | Coefficient | P-value | 95% CI | Coefficient | P-value | 95% CI |
| **Intervention area: Aberdeen** |  |  |  |  |  |  |  |  |  |
| **Staggered policy covariate** |  |  |  |  |  |  |  |  |  |
| Policy effect (all covariates) | 3.674 | <0.001 | 1.796, 5.552 | 3.805 | 0.035 | 0.267, 7.343 | 0.139 | 0.179 | -0.064, 0.343 |
| Policy effect (significant covariates) | 4.418 | <0.001 | 2.868, 5.969 | 3.970 | 0.021 | 0.609, 7.331 | 0.071 | 0.024 | 0.009, 0.134 |
| **Dummy policy covariate, sensitivity analysis** |  |  |  |  |  |  |  |  |  |
| Policy effect (all covariates) | 3.755 | 0.013 | 0.780, 6.731 | 2.950 | 0.070 | -0.235, 6.136 | 0.099 | 0.104 | -0.021, 0.219 |
| Policy effect (significant covariates) | 3.726 | 0.014 | 0.753, 6.700 | 2.140 | 0.316 | -2.041, 6.322 | 0.080 | 0.003 | 0.027, 0.134 |
| **Dummy policy covariate with policy implemented with at least half strength, sensitivity analysis** |  |  |  |  |  |  |  |  |  |
| Policy effect (all covariates) | 2.482 | 0.068 | -0.179, 5.143 | 2.286 | 0.103 | -0.459, 5.031 | 0.029 | 0.554 | -0.067, 0.125 |
| Policy effect (significant covariates) | 3.261 | <0.001 | 1.857, 4.666 | 2.352 | 0.068 | -0.172, 4.875 | 0.046 | 0.106 | -0.010, 0.101 |
| **Intervention area: Glasgow** |  |  |  |  |  |  |  |  |  |
| **Dummy policy covariate** |  |  |  |  |  |  |  |  |  |
| Policy effect (all covariates) | -0.996 | 0.453 | -3.600, 1.608 | 1.865 | 0.221 | -1.119, 4.849 | -0.010 | 0.577 | -0.043, 0.024 |
| Policy effect (significant covariates) | -1.269 | 0.139 | -2.947, 0.410 | 2.183 | 0.070 | -0.180, 4.546 | -0.001 | 0.920 | -0.024, 0.022 |

**Table A7.** Effect of policy changes on weekend restricted night-time (20:00 to 23:59) alcohol-related ambulance call-outs in Aberdeen and Glasgow.

| Primary outcome, weekend restricted night-time (20:00 to 23:59) alcohol-related ambulance call-outs | Main analysis |  |  | Sensitivity analysis |  |  |  |  |  |
| --- | --- | --- | --- | --- | --- | --- | --- | --- | --- |
|  | Outcome: number of incidents |  |  | Restricted time series only before COVID-19 restrictions, (outcome: number of incidents) |  |  | Outcome: population adjusted incident rates |  |  |
|  | Coefficient | P-value | 95% CI | Coefficient | P-value | 95% CI | Coefficient | P-value | 95% CI |
| **Intervention area: Aberdeen** |  |  |  |  |  |  |  |  |  |
| **Staggered policy covariate** |  |  |  |  |  |  |  |  |  |
| Policy effect (all covariates) | 1.716 | 0.041 | 0.072, 3.360 | 0.497 | 0.631 | -1.528, 2.521 | 0.117 | 0.044 | 0.003, 0.231 |
| Policy effect (significant covariates) | 2.408 | 0.001 | 1.000, 3.816 | 0.440 | 0.669 | -1.573, 2.453 | 0.083 | <0.001 | 0.050, 0.117 |
| **Dummy policy covariate, sensitivity analysis** |  |  |  |  |  |  |  |  |  |
| Policy effect (all covariates) | -0.196 | 0.873 | -2.606, 2.214 | 0.304 | 0.823 | -2.365, 2.973 | 0.007 | 0.842 | -0.065, 0.080 |
| Policy effect (significant covariates) | -0.334 | 0.751 | -2.396, 1.728 | -0.014 | 0.983 | -1.365, 1.337 | 0.007 | 0.859 | -0.066, 0.079 |
| **Dummy policy covariate with policy implemented with at least half strength, sensitivity analysis** |  |  |  |  |  |  |  |  |  |
| Policy effect (all covariates) | -2.118 | 0.038 | -4.115, -0.120 | 0.093 | 0.935 | -2.135, 2.322 | -0.044 | 0.180 | -0.108, 0.020 |
| Policy effect (significant covariates) | -2.040 | 0.113 | -4.564, 0.484 | -0.163 | 0.845 | -1.792, 1.466 | -0.046 | 0.156 | -0.110, 0.018 |
| **Intervention area: Glasgow** |  |  |  |  |  |  |  |  |  |
| **Dummy policy covariate** |  |  |  |  |  |  |  |  |  |
| Policy effect (all covariates) | -2.642 | 0.067 | -5.468, 0.183 | -2.281 | 0.182 | -5.630, 1.068 | -0.030 | 0.144 | -0.069, 0.010 |
| Policy effect (significant covariates) | -3.218 | <0.001 | -4.701, -1.734 | -2.183 | 0.066 | -4.513, 0.147 | -0.040 | 0.002 | -0.065, -0.014 |

**Table A8.** Effect of policy changes on weekend night-time all ambulance call-outs in Aberdeen and Glasgow.

| Secondary outcome, weekend night-time all ambulance call-outs | Main analysis |  |  | Sensitivity analysis |  |  |  |  |  |
| --- | --- | --- | --- | --- | --- | --- | --- | --- | --- |
|  | Outcome: number of incidents |  |  | Restricted time series only before COVID-19 restrictions, (outcome: number of incidents) |  |  | Outcome: population adjusted incident rates |  |  |
|  | Coefficient | P-value | 95% CI | Coefficient | P-value | 95% CI | Coefficient | P-value | 95% CI |
| **Intervention area: Aberdeen** |  |  |  |  |  |  |  |  |  |
| **Staggered policy covariate** |  |  |  |  |  |  |  |  |  |
| Policy effect (all covariates) | 11.761 | 0.005 | 3.550, 19.972 | 6.373 | 0.139 | -2.070, 14.815 | 0.328 | <0.001 | 0.189, 0.467 |
| Policy effect (significant covariates) | 12.895 | <0.001 | 6.003, 19.787 | 6.055 | 0.161 | -2.406, 14.517 | 0.284 | <0.001 | 0.145, 0.424 |
| **Dummy policy covariate, sensitivity analysis** |  |  |  |  |  |  |  |  |  |
| Policy effect (all covariates) | 5.788 | 0.197 | -3.011, 14.587 | 4.204 | 0.134 | -1.297, 9.704 | 0.240 | 0.006 | 0.068, 0.411 |
| Policy effect (significant covariates) | 7.337 | 0.102 | -1.469, 16.143 | 3.997 | 0.157 | -1.543, 9.537 | 0.202 | 0.010 | 0.048, 0.357 |
| **Dummy policy covariate with policy implemented with at least half strength, sensitivity analysis** |  |  |  |  |  |  |  |  |  |
| Policy effect (all covariates) | 8.816 | 0.010 | 2.119, 15.512 | 5.294 | 0.083 | -0.683, 11.272 | 0.227 | 0.001 | 0.094, 0.360 |
| Policy effect (significant covariates) | 9.704 | 0.001 | 4.050, 15.358 | 4.593 | 0.122 | -1.234, 10.419 | 0.208 | 0.001 | 0.084, 0.331 |
| **Intervention area: Glasgow** |  |  |  |  |  |  |  |  |  |
| **Dummy policy covariate** |  |  |  |  |  |  |  |  |  |
| Policy effect (all covariates) | -5.843 | 0.200 | -14.787, 3.102 | -3.437 | 0.468 | -12.711, 5.836 | -0.035 | 0.619 | -0.173, 0.103 |
| Policy effect (significant covariates) | -5.990 | 0.084 | -12.779, 0.798 | -1.266 | 0.763 | -9.509, 6.976 | -0.021 | 0.674 | -0.117, 0.075 |

**Table A9.** Effect of policy changes on weekend night-time alcohol-related ambulance call-outs in Aberdeen and Glasgow among female sub-group.

| Primary outcome, weekend night-time alcohol-related ambulance call-outs | Main analysis |  |  | Sensitivity analysis |  |  |  |  |  |
| --- | --- | --- | --- | --- | --- | --- | --- | --- | --- |
|  | Outcome: number of incidents |  |  | Restricted time series only before COVID-19 restrictions, (outcome: number of incidents) |  |  | Outcome: population adjusted incident rates |  |  |
|  | Coefficient | P-value | 95% CI | Coefficient | P-value | 95% CI | Coefficient | P-value | 95% CI |
| **Intervention area: Aberdeen** |  |  |  |  |  |  |  |  |  |
| **Staggered policy covariate** |  |  |  |  |  |  |  |  |  |
| Policy effect (all covariates) | 2.481 | 0.006 | 0.724, 4.237 | 8.601 | 0.012 | 1.856, 15.346 | 0.080 | <0.001 | 0.045, 0.114 |
| Policy effect (significant covariates) | 3.661 | <0.001 | 2.285, 5.037 | 2.009 | 0.044 | 0.058, 3.959 | 0.081 | <0.001 | 0.047, 0.115 |
| **Dummy policy covariate, sensitivity analysis** |  |  |  |  |  |  |  |  |  |
| Policy effect (all covariates) | 0.988 | 0.162 | -0.396, 2.372 | 0.928 | 0.168 | -0.392, 2.248 | 0.065 | <0.001 | 0.033, 0.097 |
| Policy effect (significant covariates) | 0.980 | 0.168 | -0.413, 2.373 | 0.974 | 0.152 | -0.358, 2.307 | 0.055 | <0.001 | 0.027, 0.083 |
| **Dummy policy covariate with policy implemented with at least half strength, sensitivity analysis** |  |  |  |  |  |  |  |  |  |
| Policy effect (all covariates) | 1.516 | 0.059 | -0.057, 3.090 | 1.180 | 0.158 | -0.457, 2.818 | -0.002 | 0.940 | -0.052, 0.048 |
| Policy effect (significant covariates) | 1.932 | 0.013 | 0.414, 3.450 | 1.149 | 0.148 | -0.407, 2.705 | -0.008 | 0.762 | -0.060, 0.044 |
| **Intervention area: Glasgow** |  |  |  |  |  |  |  |  |  |
| **Dummy policy covariate** |  |  |  |  |  |  |  |  |  |
| Policy effect (all covariates) | -1.597 | 0.097 | -3.485, 0.291 | -2.060 | 0.107 | -4.565, 0.444 | -0.017 | 0.232 | -0.044, 0.011 |
| Policy effect (significant covariates) | -2.104 | <0.001 | -3.070, -1.139 | -2.491 | 0.009 | -4.371, -0.611 | -0.021 | 0.014 | -0.037, -0.004 |

**Table A10.** Effect of policy changes on weekend night-time alcohol-related ambulance call-outs in Aberdeen and Glasgow among male sub-group.

| Primary outcome, weekend night-time alcohol-related ambulance call-outs | Main analysis |  |  | Sensitivity analysis |  |  |  |  |  |
| --- | --- | --- | --- | --- | --- | --- | --- | --- | --- |
|  | Outcome: number of incidents |  |  | Restricted time series only before COVID-19 restrictions, (outcome: number of incidents) |  |  | Outcome: population adjusted incident rates |  |  |
|  | Coefficient | P-value | 95% CI | Coefficient | P-value | 95% CI | Coefficient | P-value | 95% CI |
| **Intervention area: Aberdeen** |  |  |  |  |  |  |  |  |  |
| **Staggered policy covariate** |  |  |  |  |  |  |  |  |  |
| Policy effect (all covariates) | 3.401 | 0.001 | 1.357, 5.444 | 4.045 | 0.001 | 1.552, 6.537 | 0.059 | 0.021 | 0.009, 0.109 |
| Policy effect (significant covariates) | 3.726 | <0.001 | 1.946, 5.507 | 3.995 | 0.002 | 1.487, 6.502 | 0.055 | 0.025 | 0.007, 0.102 |
| **Dummy policy covariate, sensitivity analysis** |  |  |  |  |  |  |  |  |  |
| Policy effect (all covariates) | 1.943 | 0.019 | 0.321, 3.566 | 2.336 | 0.004 | 0.734, 3.938 | 0.070 | 0.004 | 0.023, 0.117 |
| Policy effect (significant covariates) | 1.977 | 0.022 | 0.280, 3.673 | 2.261 | 0.017 | 0.404, 4.119 | 0.080 | <0.001 | 0.037, 0.123 |
| **Dummy policy covariate with policy implemented with at least half strength, sensitivity analysis** |  |  |  |  |  |  |  |  |  |
| Policy effect (all covariates) | 1.894 | 0.025 | 0.243, 3.545 | 2.114 | 0.040 | 0.098, 4.131 | 0.030 | 0.196 | -0.015, 0.075 |
| Policy effect (significant covariates) | 2.315 | 0.002 | 0.815, 3.816 | 1.818 | 0.077 | -0.200, 3.835 | 0.030 | 0.148 | -0.011, 0.070 |
| **Intervention area: Glasgow** |  |  |  |  |  |  |  |  |  |
| **Dummy policy covariate** |  |  |  |  |  |  |  |  |  |
| Policy effect (all covariates) | 2.392 | 0.300 | -2.131, 6.916 | -0.395 | 0.780 | -3.162, 2.372 | 0.036 | 0.382 | -0.044, 0.115 |
| Policy effect (significant covariates) | 1.463 | 0.500 | -2.783, 5.709 | -2.150 | 0.053 | -4.330, 0.029 | 0.019 | 0.616 | -0.054, 0.092 |

**Table A11.** Effect of policy changes on weekend night-time alcohol-related ambulance call-outs in Aberdeen and Glasgow among patients age less than 45 years.

| Primary outcome, weekend night-time alcohol-related ambulance call-outs | Main analysis |  |  | Sensitivity analysis |  |  |  |  |  |
| --- | --- | --- | --- | --- | --- | --- | --- | --- | --- |
|  | Outcome: number of incidents |  |  | Restricted time series only before COVID-19 restrictions, (outcome: number of incidents) |  |  | Outcome: population adjusted incident rates |  |  |
|  | Coefficient | P-value | 95% CI | Coefficient | P-value | 95% CI | Coefficient | P-value | 95% CI |
| **Intervention area: Aberdeen** |  |  |  |  |  |  |  |  |  |
| **Staggered policy covariate** |  |  |  |  |  |  |  |  |  |
| Policy effect (all covariates) | 6.180 | 0.002 | 2.251, 10.109 | 15.626 | <0.001 | 10.292, 20.959 | 0.210 | 0.002 | 0.074, 0.345 |
| Policy effect (significant covariates) | 7.050 | <0.001 | 4.077, 10.023 | 14.904 | <0.001 | 9.983, 19.825 | 0.105 | <0.001 | 0.065, 0.145 |
| **Dummy policy covariate, sensitivity analysis** |  |  |  |  |  |  |  |  |  |
| Policy effect (all covariates) | 1.527 | 0.648 | -5.026, 8.080 | 1.897 | 0.406 | -2.577, 6.371 | 0.066 | 0.123 | -0.018, 0.150 |
| Policy effect (significant covariates) | 1.355 | 0.705 | -5.670, 8.380 | 1.860 | 0.390 | -2.384, 6.104 | 0.088 | <0.001 | 0.050, 0.127 |
| **Dummy policy covariate with policy implemented with at least half strength, sensitivity analysis** |  |  |  |  |  |  |  |  |  |
| Policy effect (all covariates) | 1.500 | 0.873 | -16.828, 19.827 | 2.821 | 0.245 | -1.936, 7.577 | 0.025 | 0.423 | -0.037, 0.088 |
| Policy effect (significant covariates) | 3.732 | 0.562 | -8.876, 16.340 | 2.595 | 0.179 | -1.188, 6.378 | 0.068 | <0.001 | 0.035, 0.101 |
| **Intervention area: Glasgow** |  |  |  |  |  |  |  |  |  |
| **Dummy policy covariate** |  |  |  |  |  |  |  |  |  |
| Policy effect (all covariates) | -0.889 | 0.516 | -3.575, 1.797 | 0.908 | 0.577 | -2.282, 4.099 | -0.011 | 0.633 | -0.054, 0.033 |
| Policy effect (significant covariates) | -2.543 | 0.014 | -4.579, -0.506 | 2.250 | 0.110 | -0.510, 5.010 | -0.013 | 0.513 | -0.050, 0.025 |

**Table A12.** Effect of policy changes on weekend night-time alcohol-related ambulance call-outs in Aberdeen and Glasgow among patients age more than 45 years.

| Primary outcome, weekend night-time alcohol-related ambulance call-outs | Main analysis |  |  | Sensitivity analysis |  |  |  |  |  |
| --- | --- | --- | --- | --- | --- | --- | --- | --- | --- |
|  | Outcome: number of incidents |  |  | Restricted time series only before COVID-19 restrictions, (outcome: number of incidents) |  |  | Outcome: population adjusted incident rates |  |  |
|  | Coefficient | P-value | 95% CI | Coefficient | P-value | 95% CI | Coefficient | P-value | 95% CI |
| **Intervention area: Aberdeen** |  |  |  |  |  |  |  |  |  |
| **Staggered policy covariate** |  |  |  |  |  |  |  |  |  |
| Policy effect (all covariates) | 0.891 | 0.689 | -3.478, 5.261 | 0.315 | 0.877 | -3.664, 4.295 | -0.010 | 0.870 | -0.131, 0.111 |
| Policy effect (significant covariates) | 0.806 | 0.009 | 0.203, 1.408 | 0.357 | 0.681 | -1.346, 2.060 | 0.033 | 0.054 | -0.001, 0.066 |
| **Dummy policy covariate, sensitivity analysis** |  |  |  |  |  |  |  |  |  |
| Policy effect (all covariates) | 0.925 | 0.288 | -0.783, 2.633 | 0.886 | 0.355 | -0.992, 2.764 | 0.018 | 0.544 | -0.040, 0.076 |
| Policy effect (significant covariates) | 0.679 | 0.008 | 0.181, 1.178 | 0.499 | 0.365 | -0.580, 1.578 | 0.046 | 0.004 | 0.015, 0.077 |
| **Dummy policy covariate with policy implemented with at least half strength, sensitivity analysis** |  |  |  |  |  |  |  |  |  |
| Policy effect (all covariates) | 0.163 | 0.857 | -1.616, 1.943 | -0.341 | 0.697 | -2.057, 1.376 | 0.008 | 0.761 | -0.046, 0.062 |
| Policy effect (significant covariates) | 0.571 | 0.053 | -0.008, 1.150 | 0.043 | 0.948 | -1.243, 1.328 | 0.025 | 0.076 | -0.003, 0.053 |
| **Intervention area: Glasgow** |  |  |  |  |  |  |  |  |  |
| **Dummy policy covariate** |  |  |  |  |  |  |  |  |  |
| Policy effect (all covariates) | -1.139 | 0.458 | -4.144, 1.867 | -4.629 | <0.001 | -6.691, -2.567 | -0.055 | <0.001 | -0.084, -0.027 |
| Policy effect (significant covariates) | -1.566 | 0.293 | -4.484, 1.352 | -3.452 | <0.001 | -4.713, -2.190 | -0.066 | <0.001 | -0.090, -0.042 |

**Table A13.** Effect of policy changes on weekend restricted night-time (24:00 to 05:59) recorded crimes in Aberdeen and Glasgow.

| Secondary outcome, weekend restricted night-time (24:00 to 05:59) recorded crimes | Main analysis |  |  | Sensitivity analysis |  |  |  |  |  |
| --- | --- | --- | --- | --- | --- | --- | --- | --- | --- |
|  | Outcome: number of incidents |  |  | Restricted time series only before COVID-19 restrictions, (outcome: number of incidents) |  |  | Outcome: population adjusted incident rates |  |  |
|  | Coefficient | P-value | 95% CI | Coefficient | P-value | 95% CI | Coefficient | P-value | 95% CI |
| **Intervention area: Aberdeen** |  |  |  |  |  |  |  |  |  |
| **Staggered policy covariate** |  |  |  |  |  |  |  |  |  |
| Policy effect (all covariates) | 4.534 | 0.071 | -0.385, 9.454 | 3.752 | 0.179 | -1.721, 9.226 | 0.034 | 0.717 | -0.152, 0.221 |
| Policy effect (significant covariates) | 2.853 | 0.002 | 1.073, 4.634 | 3.483 | 0.003 | 1.153, 5.814 | 0.023 | 0.497 | -0.043, 0.090 |
| **Dummy policy covariate, sensitivity analysis** |  |  |  |  |  |  |  |  |  |
| Policy effect (all covariates) | -0.060 | 0.967 | -2.917, 2.797 | -0.062 | 0.970 | -3.239, 3.116 | 0.004 | 0.950 | -0.110, 0.118 |
| Policy effect (significant covariates) | 1.572 | 0.028 | 0.174, 2.970 | 1.805 | 0.033 | 0.150, 3.460 | 0.031 | 0.325 | -0.030, 0.091 |
| **Dummy policy covariate with policy implemented with at least half strength, sensitivity analysis** |  |  |  |  |  |  |  |  |  |
| Policy effect (all covariates) | 2.757 | 0.007 | 0.759, 4.755 | 2.291 | 0.042 | 0.079, 4.503 | 0.076 | 0.023 | 0.011, 0.142 |
| Policy effect (significant covariates) | 2.642 | <0.001 | 1.271, 4.013 | 2.821 | 0.001 | 1.171, 4.472 | 0.048 | 0.043 | 0.002, 0.094 |
| **Intervention area: Glasgow** |  |  |  |  |  |  |  |  |  |
| **Dummy policy covariate** |  |  |  |  |  |  |  |  |  |
| Policy effect (all covariates) | -6.301 | 0.003 | -10.423, -2.178 | -1.869 | 0.280 | -5.256, 1.518 | -0.108 | 0.002 | -0.175, -0.040 |
| Policy effect (significant covariates) | -5.863 | 0.003 | -9.687, -2.040 | -0.625 | 0.654 | -3.356, 2.106 | -0.100 | 0.002 | -0.164, -0.035 |

### Appendix 2.4: Falsification test

**Table A14.**
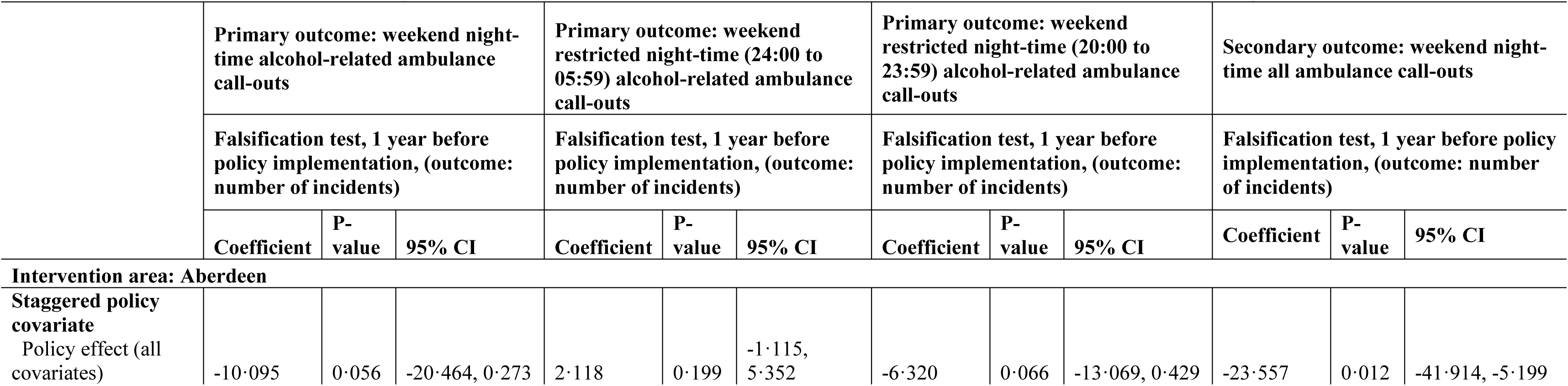
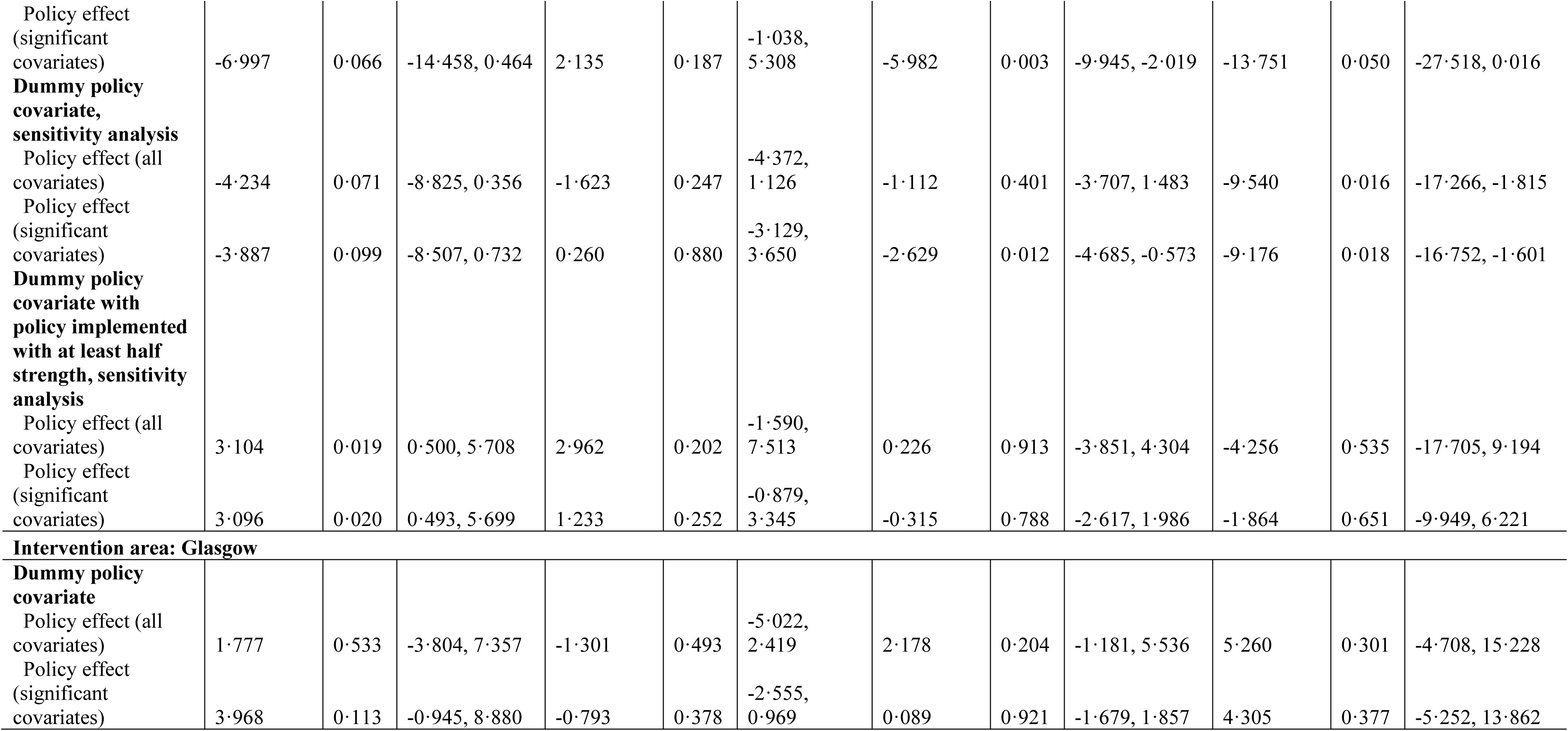
Falsification test for weekend night-time alcohol-related ambulance call-outs and all ambulance call-outs in Aberdeen and Glasgow.

**Table A15.**
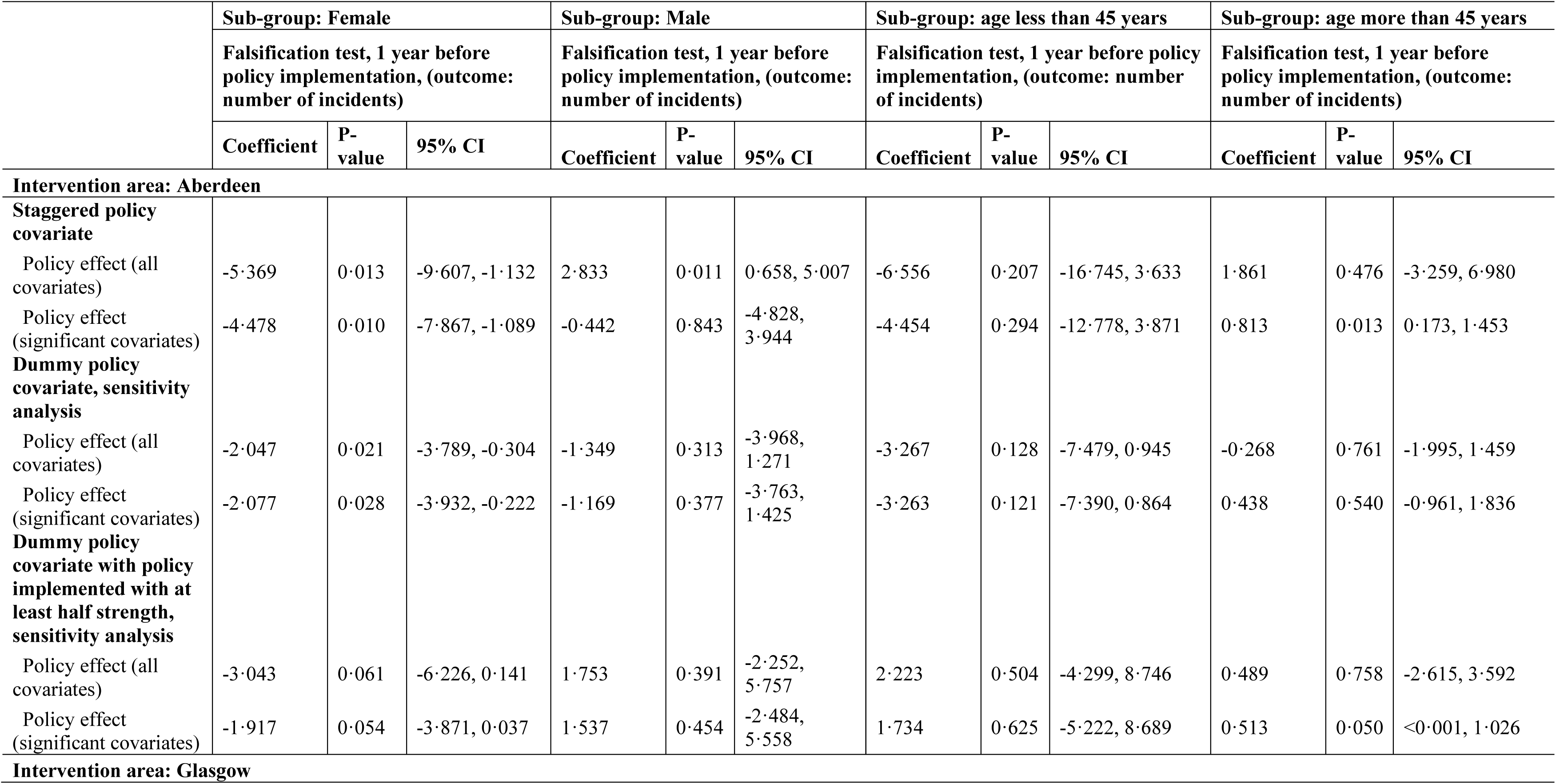
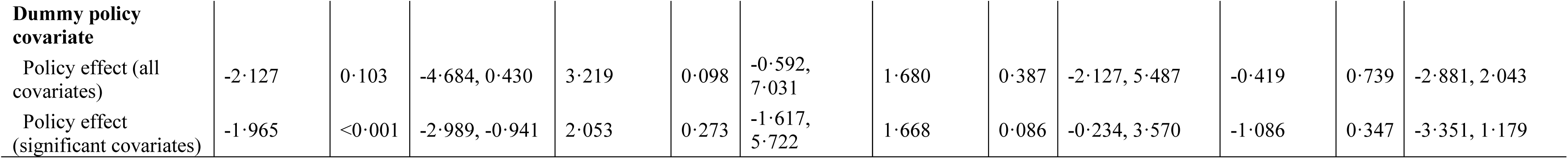
Falsification test for sub-group analysis for the weekend night-time alcohol-related ambulance call-outs in Aberdeen and Glasgow.

**Table A16.**
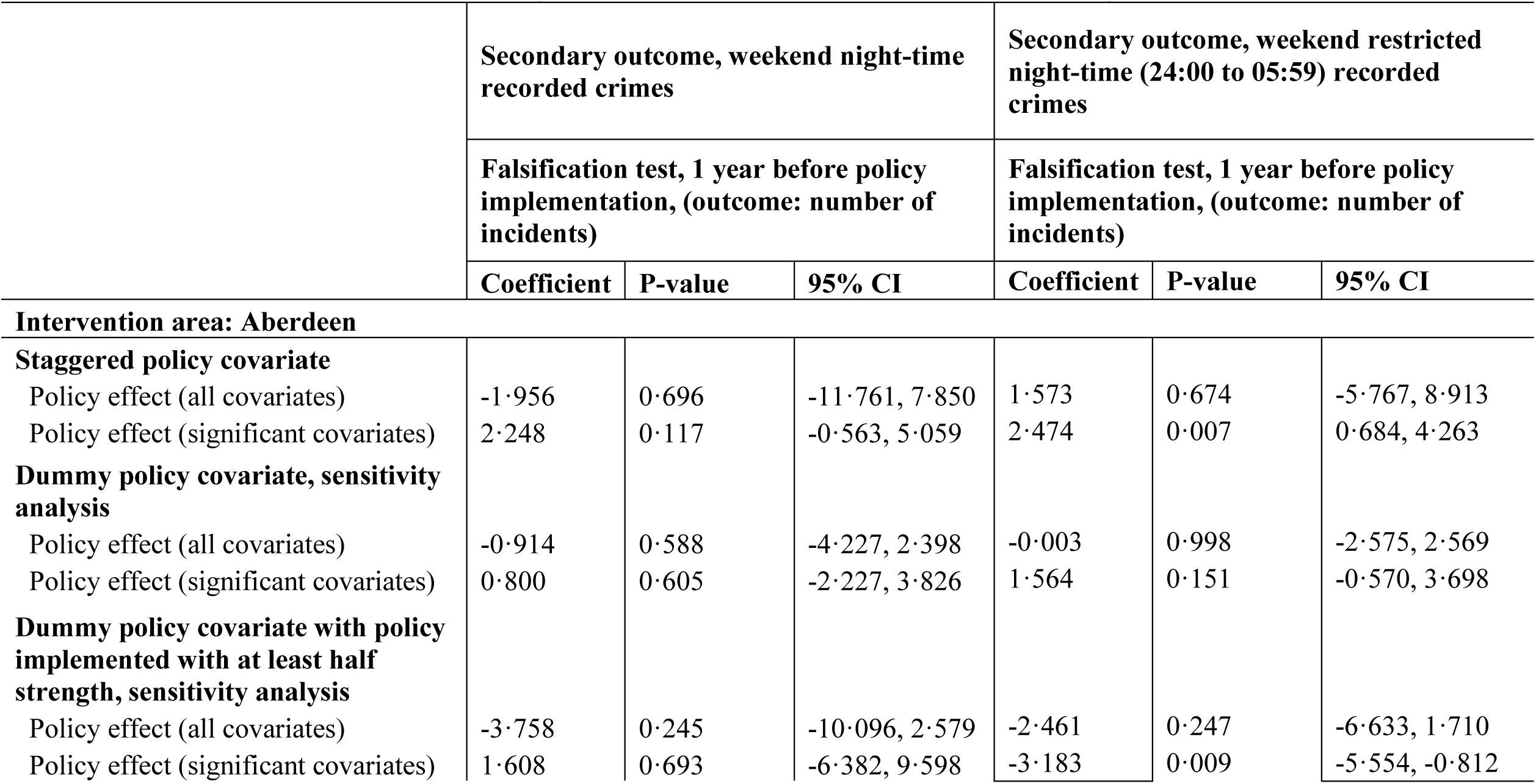
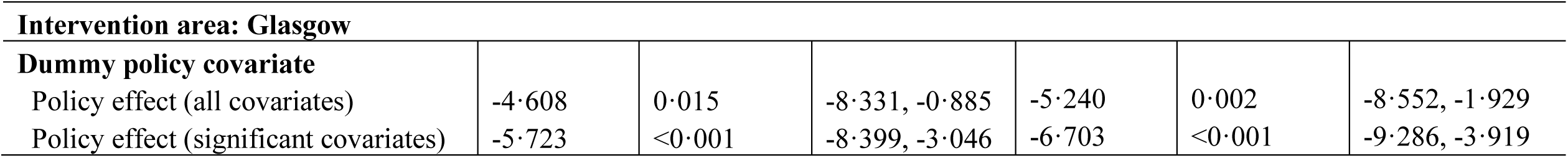
Falsification test for weekend night-time recorded crimes in Aberdeen and Glasgow.

## Appendix 3. Synthetic control

### Appendix 3.1. Alcohol-related ambulance call-outs (Aberdeen)

#### Appendix 3.1.1. Model specification and validation test

The analysis of the synthetic control was conducted in two main steps. First, we assessed nine different model specifications derived from the existing literature on synthetic control methods, following the recommended validation procedure for synthetic control analysis (Table SC 1) [39,40]. The validation test involved calculating the mean squared prediction error (MSPE) by splitting the pre-intervention periods into training and validation sets. Since there are no recommended ratios for splitting the pre-intervention period, we tested three different split ratios—50:50, 70:30, and 80:20— and compared MSPEs across the different specifications. For the alcohol-related ambulance call-outs in Aberdeen, we identified three specifications (1, 2, and 3 (see below Table SC 1)) that produced the lowest MSPEs. However, literature recommends against using all pre-intervention outcome values as covariates, as this can reduce the predictive power of other variables that may significantly influence the construction of the counterfactual [40,41]. Therefore, we selected specification 3 for the remainder of the synthetic control analysis. This specification was chosen because it struck a balance between capturing the pre-intervention trends and allowing other important covariates to contribute to the counterfactual.

#### Appendix 3.1.2. Main analysis and sensitivity tests

For the analysis of alcohol-related ambulance call-outs, we observed that the predictor means for Aberdeen were closely aligned with those of the synthetic Aberdeen, as opposed to the average of all donor pool/control cities (Table SC 2). The number of on-premises alcohol outlets emerged as the most influential covariate in generating the synthetic control weights, alongside the pre-intervention outcome mean. Among the donor pools, Edinburgh was the most significant contributor to the creation of synthetic Aberdeen, followed by North Lanarkshire and East Renfrewshire (Figure SC 01).

From the gap plot between Aberdeen and synthetic Aberdeen, a positive average treatment effect (ATT=0·103) appeared evident (Figure SC 02). However, a placebo test using permutation methods indicated that other council areas might also exhibit a positive ATT although no policy change occurred there (Figure SC 03a). As a sensitivity analysis, we also presented two graphs for the placebo test after excluding council areas with MSPE two times higher and one time higher than Aberdeen’s (Figures SC 03b and SC 03c). This exclusion aimed to improve the robustness of the comparison.

For Aberdeen, we calculated the ATT by accounting for the staggered implementation of the policy, adjusting the ATT accordingly using staggered policy weights. Additionally, we visualised the distribution of effect sizes (i.e., ATT) based on the placebo test, where the second figure only considered positive effect sizes, further narrowing the focus to more relevant comparisons (Figures SC 04a and SC 04b). This distribution provides a clear depiction of how extreme the ATT for Aberdeen is compared to the control/donor cities. From the figures, we observed that while some control units exhibited positive ATTs, the magnitude of Aberdeen’s ATT was comparatively larger. This suggests that the treatment effect in Aberdeen is more pronounced than in most control cities, strengthening the uniqueness of the observed impact in Aberdeen relative to the donor pool.

Finally, we compared post/pre-intervention MSPE ratios for Aberdeen and the control cities and calculated a frequentist p-value. This was done by dividing the number of cities with post/pre MSPE ratios higher than Aberdeen’s by the total number of cities. This resulted in a p-value of 0·222 (6/27) (Figure SC 05a). The second figure illustrating post/pre MSPE ratios was also presented, this time only considering cities with positive effect sizes, enhancing the interpretability of the results with a focus on potential treatment effects (Figure SC 05b). The estimated p-value in this analysis was 0·28, calculated as 5/18 (Figure SC 05b). It is important to note that the minimum possible p-value in synthetic control studies depends on the number of control units included in the donor pool. For instance, with 27 control units, the minimum attainable p-value would be 1/27 (approximately 0·04). In contrast, if 18 control units are used, the minimum p-value would be 1/18, (approximately 0·06). Thus, the number of control units directly influences the lowest possible p-value that can be observed. In addition, we conducted ARIMA based on difference-in-difference analyses with the synthetic control where we modelled the differences of alcohol-related ambulance call-out rates between Aberdeen and Synthetic Aberdeen [42]. We estimated effect size of 0·143 with a p-value <0·001 (Table A17).

**Table SC 1.** Validation test for synthetic control for alcohol-related ambulance call-outs in Aberdeen.

| Specifications | Pre-intervention periods split between training and validation |  |  |
| --- | --- | --- | --- |
|  | Training/validation, 50/50 | Training/validation, 70/30 | Training/validation, 80/20 |
|  | MSPE* (Validation) | MSPE* (Validation) | MSPE* (Validation) |
| 1. (all pre-intervention outcome values and covariates) | 0.0336 | 0.03522 | 0.0480 |
| 2. (all pre-intervention outcome values only) | 0.0335 | 0.03524 | 0.0479 |
| 3. (pre-intervention outcome mean and covariates) | 0.0412 | 0.0384 | 0.0435 |
| 4. (last pre-intervention outcome value and covariates) | 0.0807 | 0.1267 | 0.1002 |
| 5. (pre-intervention outcome mean and last pre-intervention outcome value, and covariates) | 0.0404 | 0.0389 | 0.0473 |
| 6. (first three-fourths of the pre-intervention outcome values and covariates) | 0.0370 | 0.0411 | 0.0503 |
| 7. (last three-fourths of the pre-intervention outcome values and covariates) | 0.0356 | 0.0386 | 0.0528 |
| 8. (odd pre-intervention outcome values and covariates) | 0.0359 | 0.0388 | 0.0516 |
| 9. (even pre-intervention outcome values and covariates) | 0.0367 | 0.0402 | 0.0458 |

**\*Mean Square Prediction Error**

##### Aberdeen- Synthetic Aberdeen, Specification 3

**Table SC 2.**
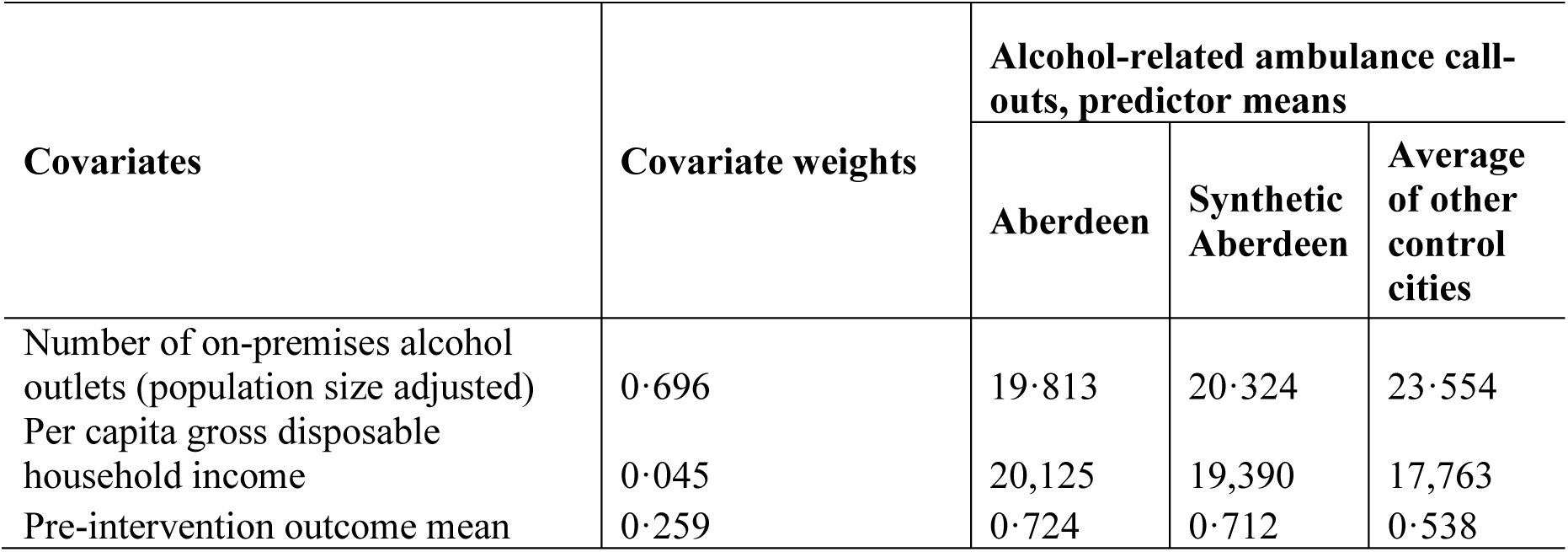
Covariate balance in the pre-intervention periods.

| Covariates | Covariate weights | Alcohol-related ambulance call-outs, predictor means |  |  |
| --- | --- | --- | --- | --- |
|  |  | Aberdeen | Synthetic Aberdeen | Average of other control cities |
| Number of on-premises alcohol outlets (population size adjusted) | 0.696 | 19.813 | 20.324 | 23.554 |
| Per capita gross disposable household income | 0.045 | 20,125 | 19,390 | 17,763 |
| Pre-intervention outcome mean | 0.259 | 0.724 | 0.712 | 0.538 |

**Figure SC 01:**
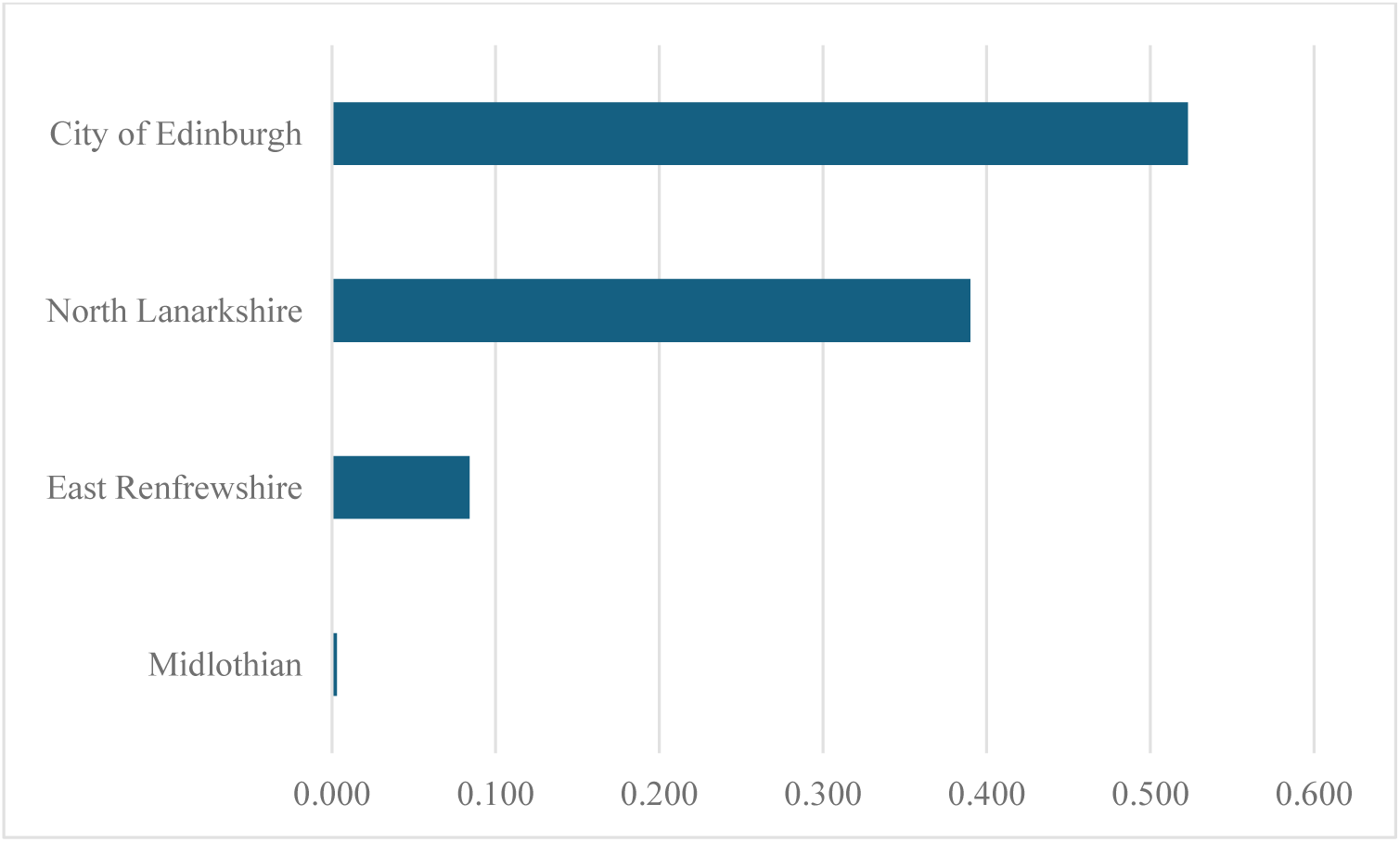
Optimal unit weights.

**Figure SC 02:**
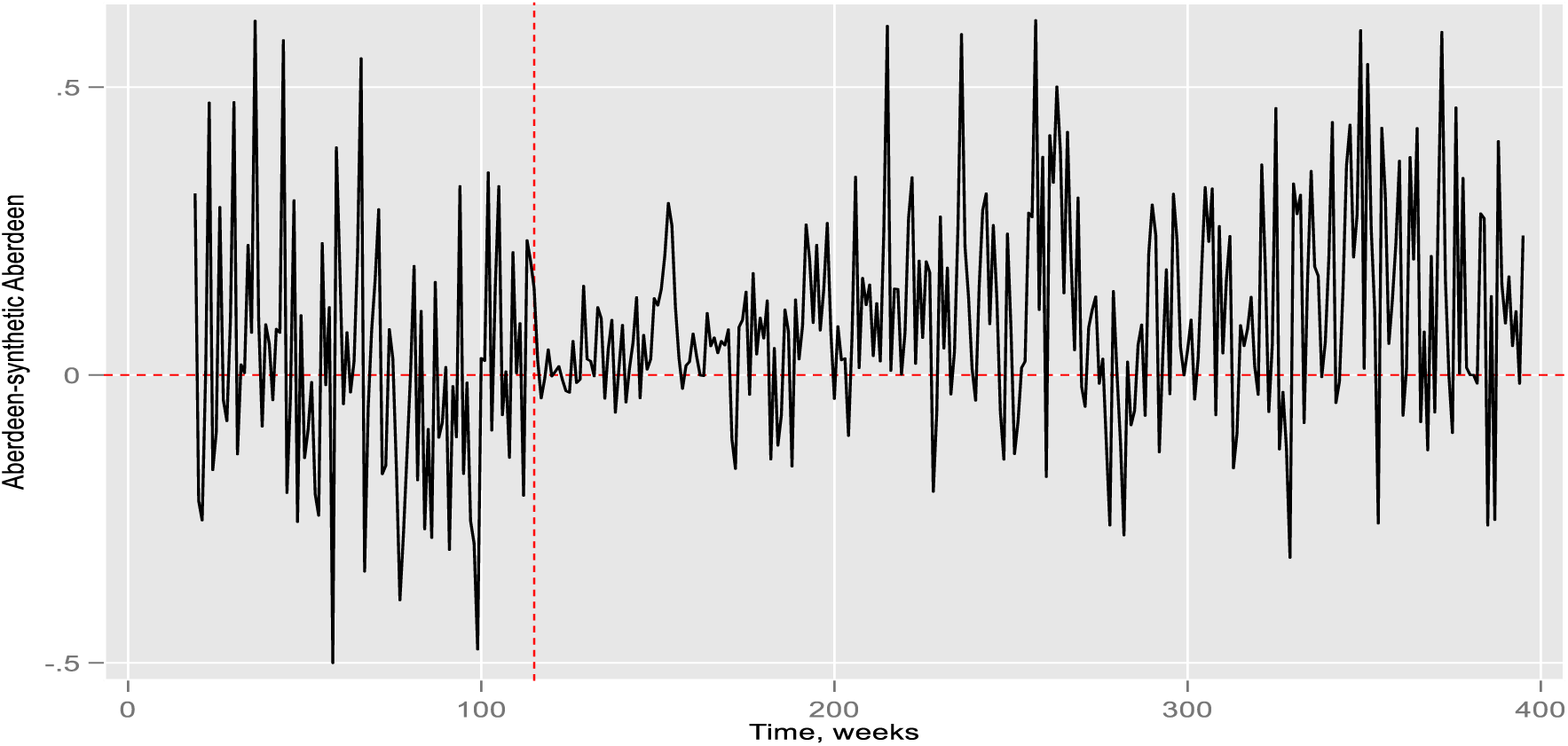
Gaps of Aberdeen and synthetic Aberdeen.

**Figure SC 03a:**
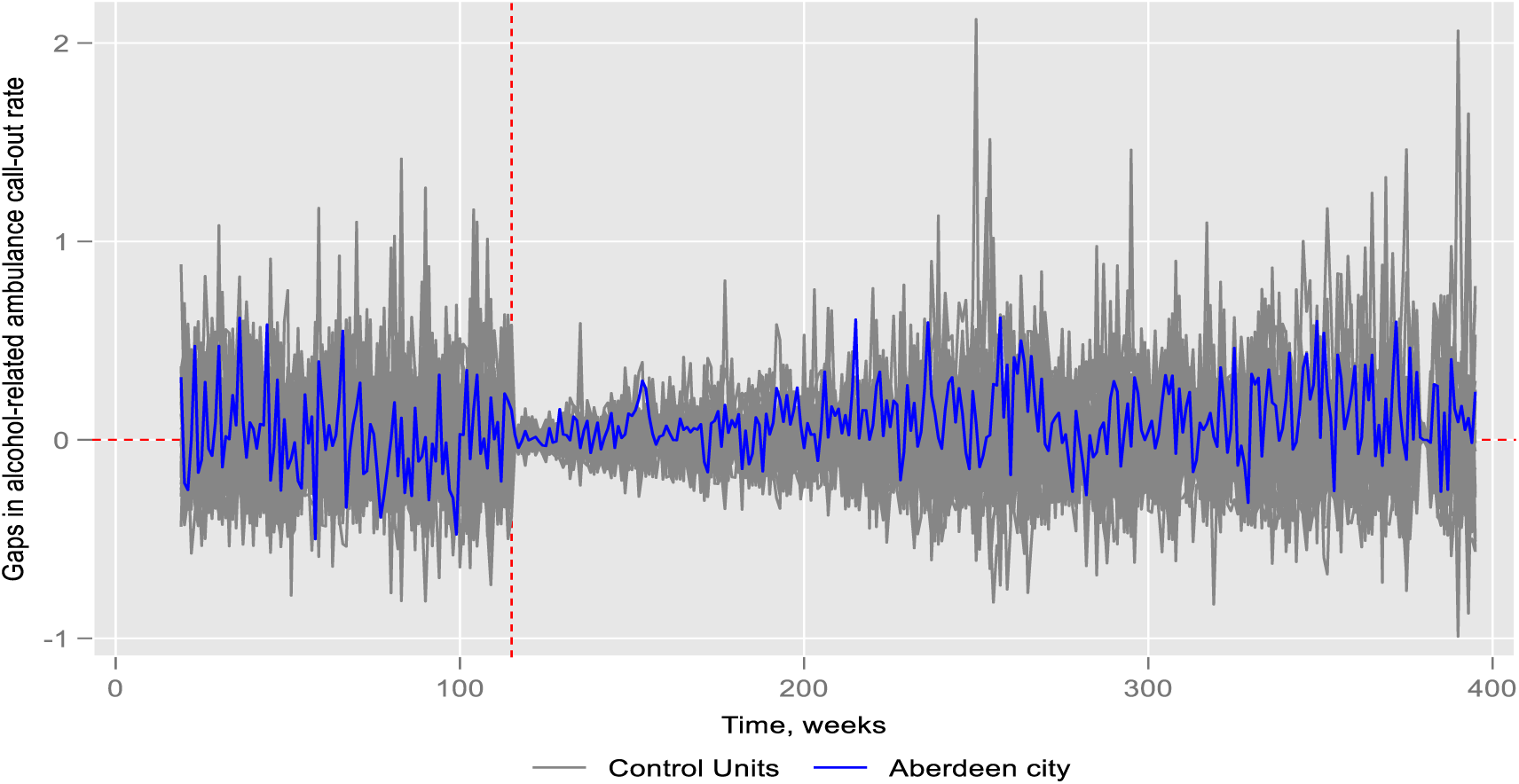
Placebo test.

**Figure SC 03b:**
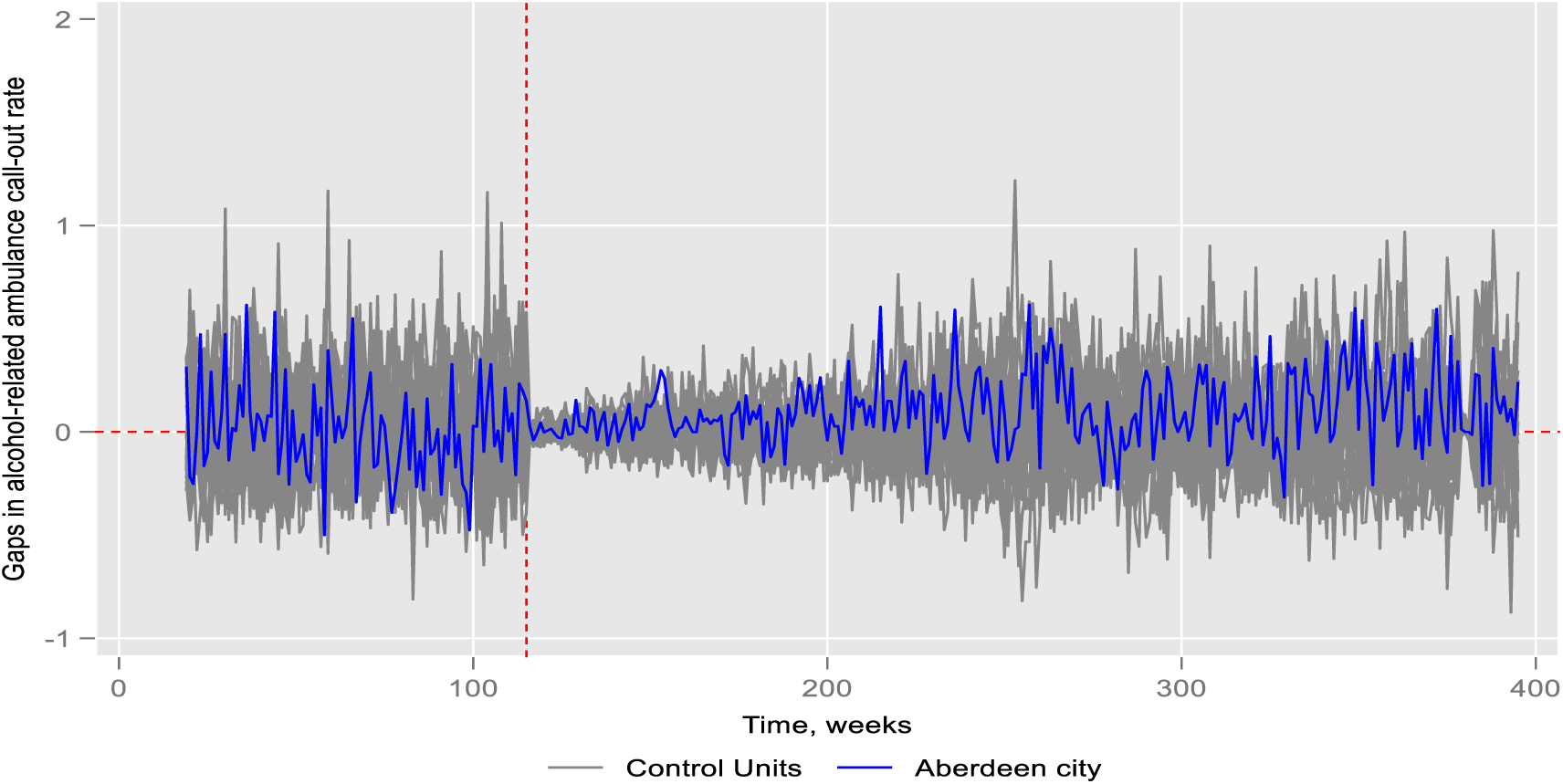
Placebo test excluding control cities MSPE>2 times Aberdeen.

**Figure SC 03c:**
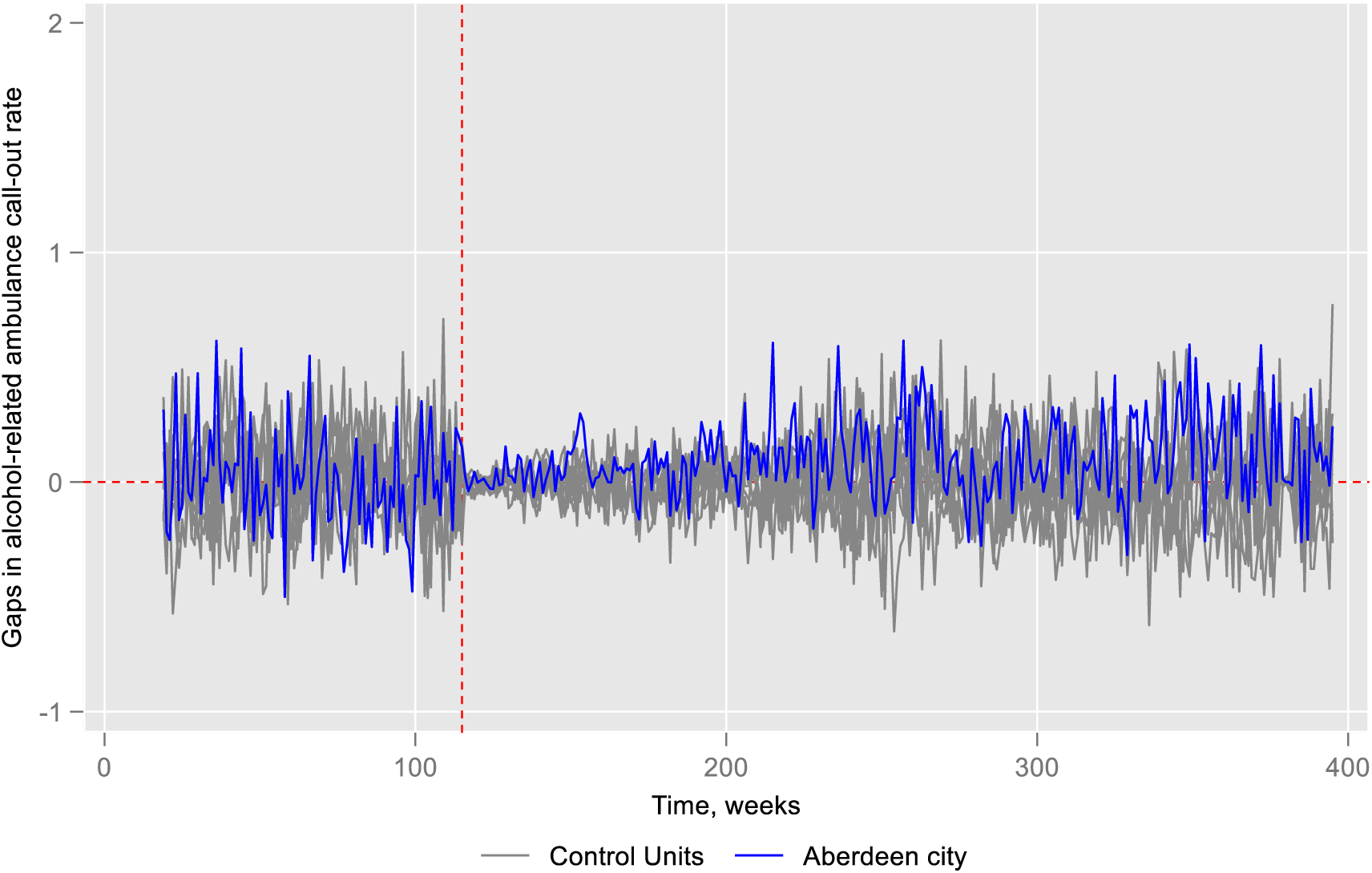
Placebo test excluding control cities if MSPE> Aberdeen.

**Figure SC 04a:**
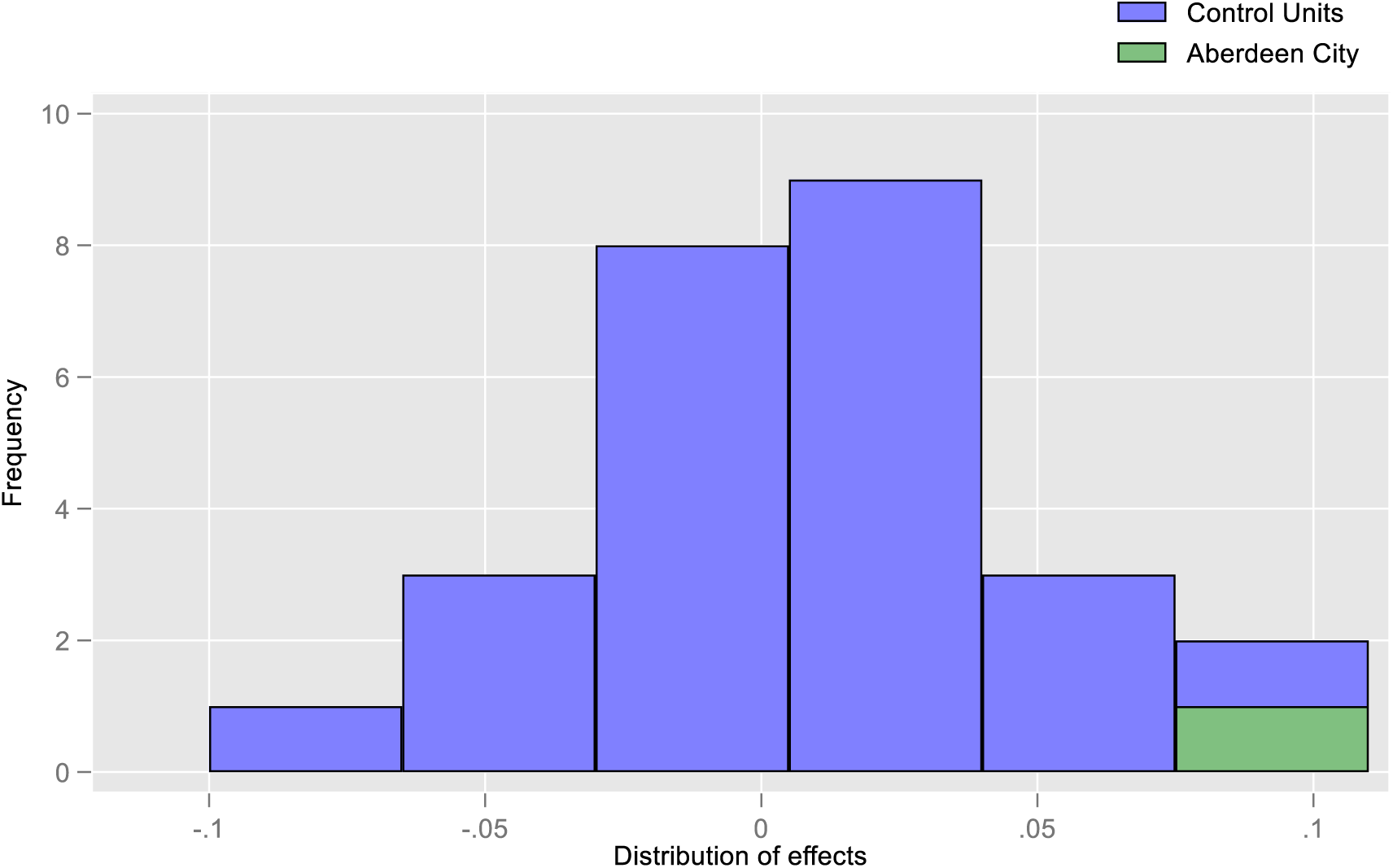
Effect size for placebo test.

**Figure SC 04b:**
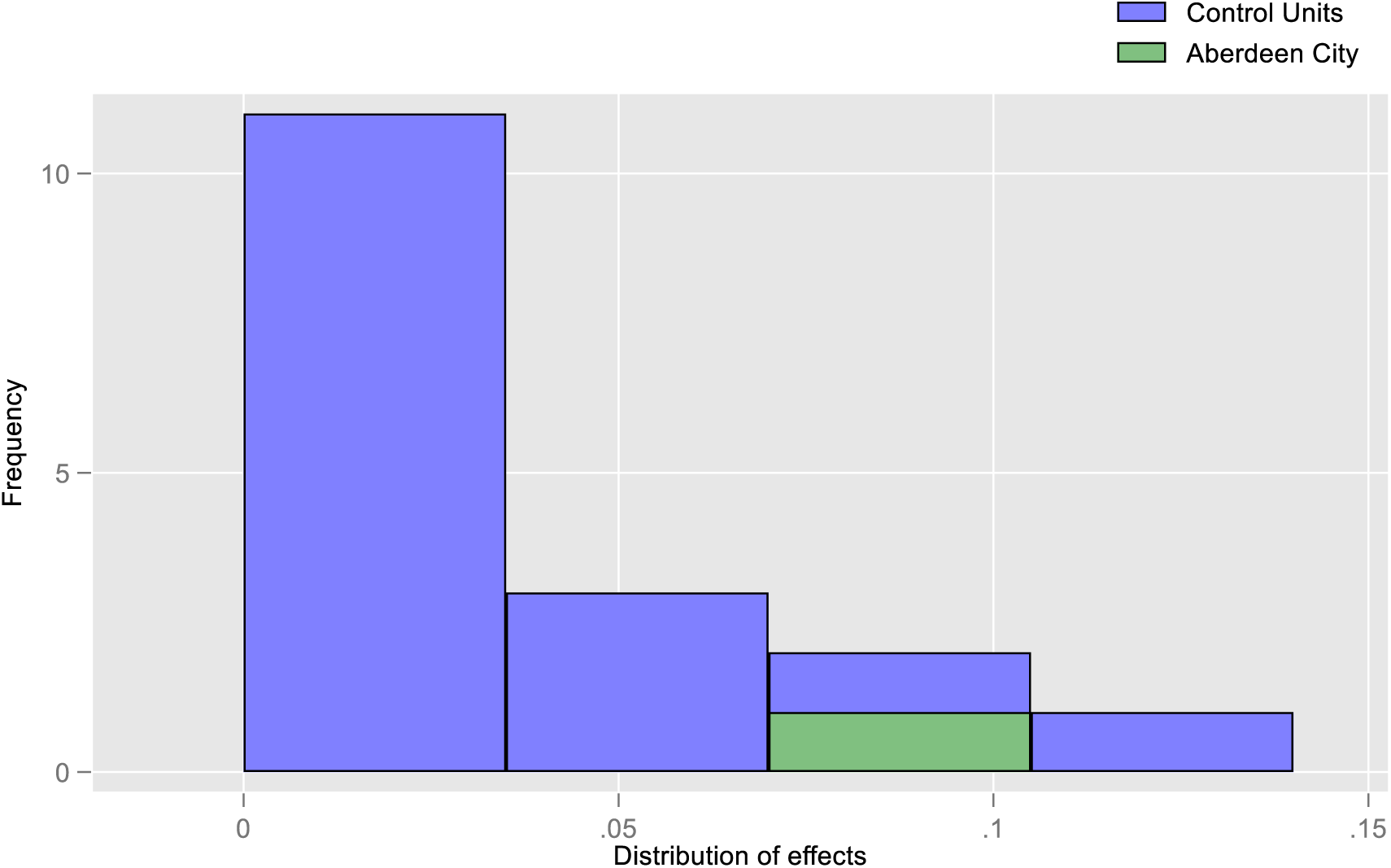
Effect size placebo test considering only positive effect size.

**Figure SC 05a:**
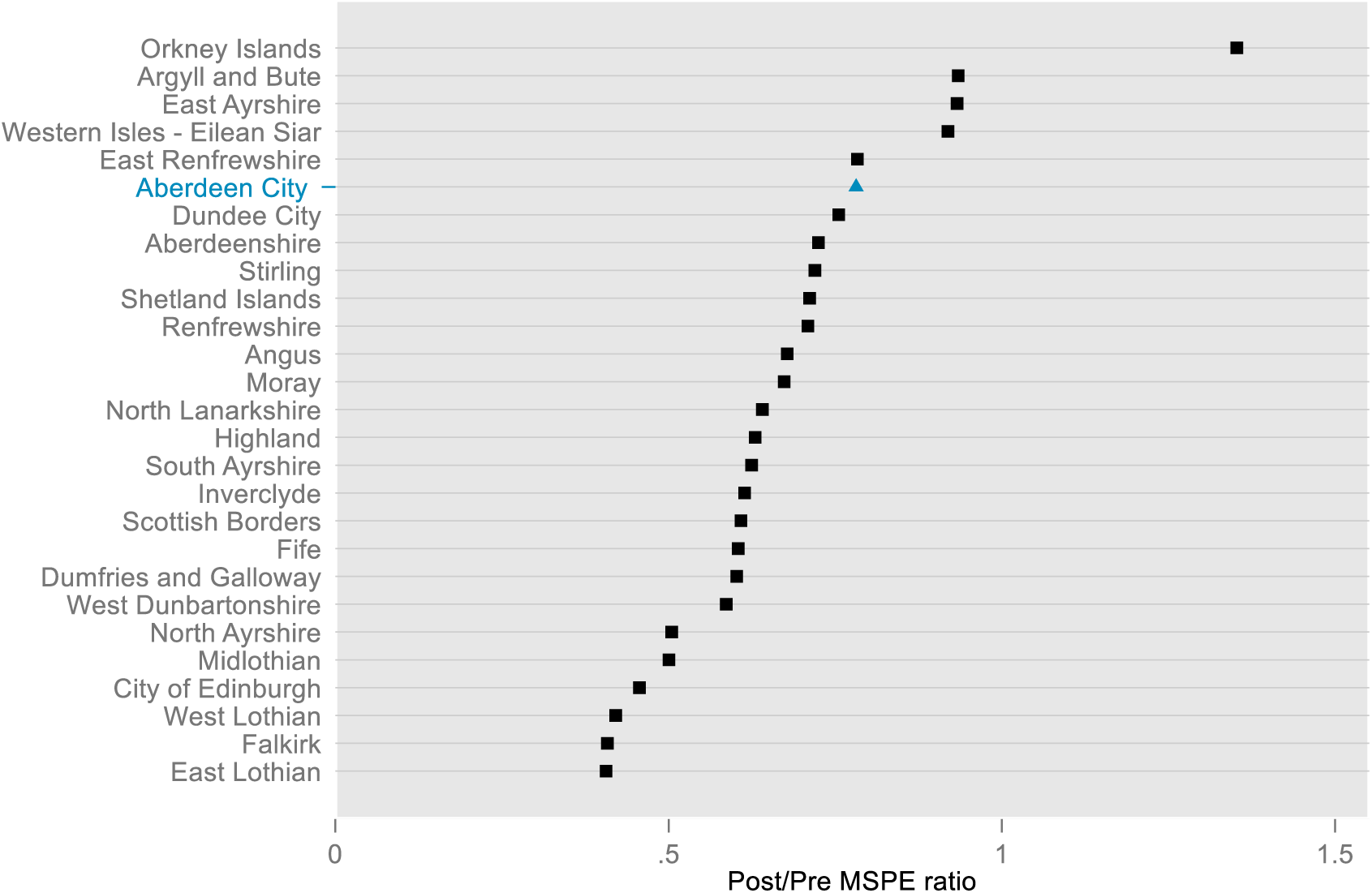
Post/Pre MSPE ratio across cities.

**Figure SC 05b:**
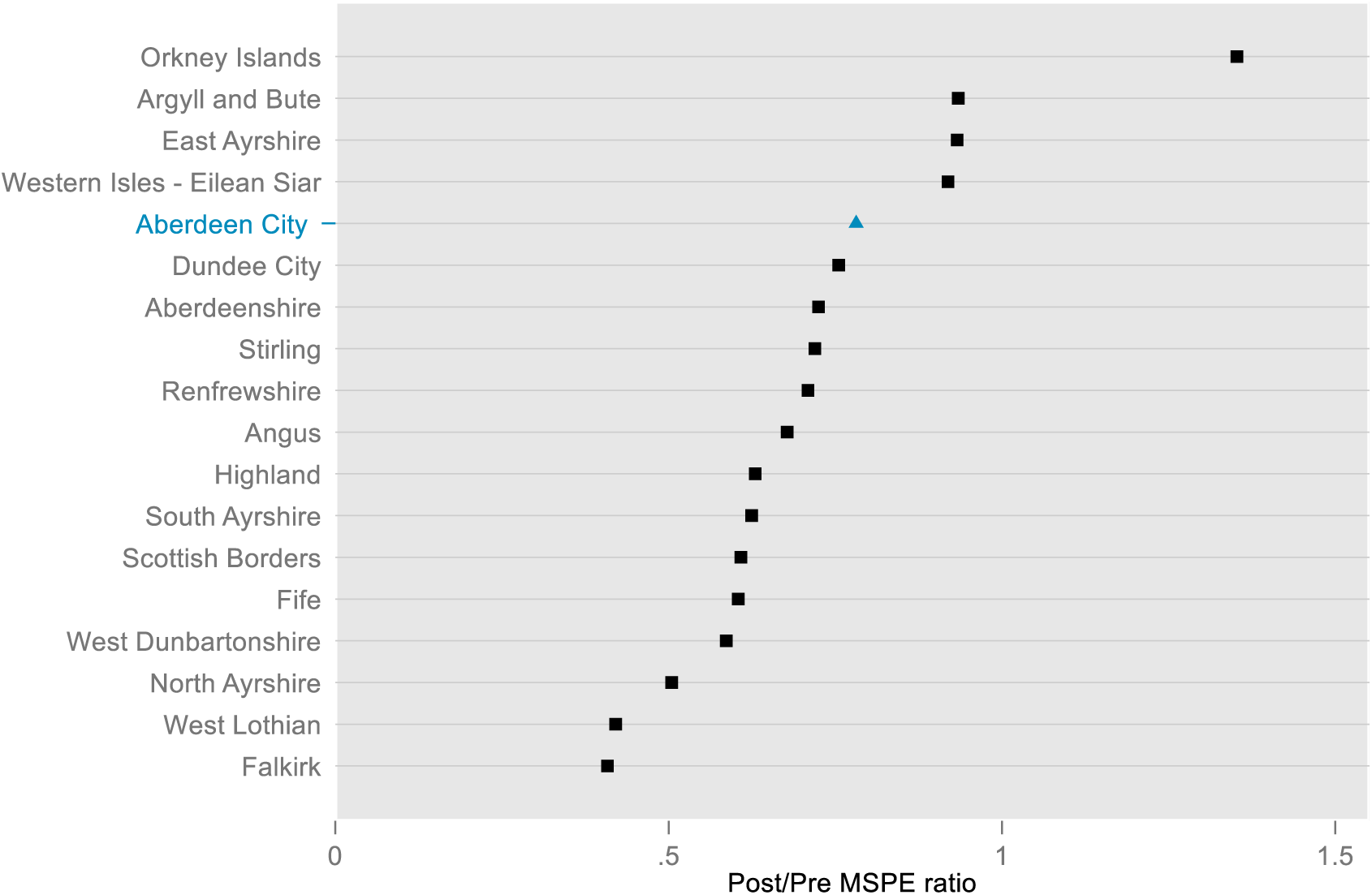
Post/Pre MSPE ratio across cities considering only positive effect size.

### Appendix 3.2. Alcohol-related ambulance call-outs (Glasgow)

#### Appendix 3.2.1. Model specification and validation test

For the analysis of alcohol-related ambulance call-outs in Glasgow, we identified two specifications (3 and 7) that yielded the lowest MSPEs (Table SC 3). Specification 3 produced the smallest MSPE on two occasions and had the lowest overall value. As a result, we selected specification 3 for the remainder of the analysis.

#### Appendix 3.2.2. Main analysis and sensitivity tests

We observed that the predictor means for Glasgow were closely aligned with those of the synthetic Glasgow, compared to the average of all donor pool/control cities (Table SC 4). Per capita gross disposable household income (63%) and the number of on-premises alcohol outlets (37%) emerged as the most influential covariates in generating the synthetic control weights. Among the donor pool cities, Dundee and North Ayrshire were the most significant contributors to the creation of synthetic Glasgow (Figure SC 06).

For Glasgow, we estimated a positive average treatment effect (ATT) of 0·012. However, the effect sizes estimated using ARIMA models were negative, indicating a discrepancy between the synthetic control and ARIMA approaches. Additionally, the p-value from the placebo test was 0·926, indicating a high level of uncertainty (Figure SC 10a). When we further modelled ARIMA with synthetic control, we estimated negative and statistically significant effect sizes (Table A17). Despite the different nature of p-values (inferential for ARIMA and frequentist for Synthetic) the different results from different approaches in terms of direction and significance weakened our confidence in interpreting the overall impact of the policy change in Glasgow, making these results inconclusive.

**Table SC 3.** Validation test for synthetic control for alcohol-related ambulance call-outs in Glasgow.

| Specifications | Pre-intervention periods split between training and validation |  |  |
| --- | --- | --- | --- |
|  | Training/validation, 50/50 | Training/validation, 70/30 | Training/validation, 80/20 |
|  | MSPE* (Validation) | MSPE* (Validation) | MSPE* (Validation) |
| 1. (all pre-intervention outcome values and covariates) | 0.0618 | 0.05474 | 0.0662 |
| 2. (all pre-intervention outcome values only) | 0.0618 | 0.05474 | 0.0662 |
| 3. (pre-intervention outcome mean and covariates) | 0.0604 | 0.0667 | 0.0489 |
| 4. (last pre-intervention outcome value and covariates) | 0.0810 | 0.0706 | 0.0605 |
| 5. (pre-intervention outcome mean and last pre-intervention outcome value, and covariates) | 0.0604 | 0.0532 | 0.0729 |
| 6. (first three-fourths of the pre-intervention outcome values and covariates) | 0.0643 | 0.0606 | 0.0662 |
| 7. (last three-fourths of the pre-intervention outcome values and covariates) | 0.0613 | 0.0531 | 0.0620 |
| 8. (odd pre-intervention outcome values and covariates) | 0.0618 | 0.0547 | 0.0662 |
| 9. (even pre-intervention outcome values and covariates) | 0.0618 | 0.0547 | 0.0662 |

##### Glasgow- Synthetic Glasgow, Specification 3

**Table SC 4.** Covariate balance in the pre-intervention periods.

**Glasgow- Synthetic Glasgow, Specification 3:** **Table SC 4. Covariate balance in the pre-intervention periods**
| Covariates | Covariate weights | Alcohol-related ambulance call-outs, predictor means |  |  |
| --- | --- | --- | --- | --- |
|  |  | Glasgow | Synthetic Glasgow | Average of other control cities |
| Number of on-premises alcohol outlets (population size adjusted) | 0·370 | 21·416 | 21·446 | 23·524 |
| Per capita gross disposable household income | 0·628 | 15852 | 15851 | 18260 |
| Pre-intervention outcome mean | 0·002 | 0·964 | 0·835 | 0·595 |

**Figure SC 06:**
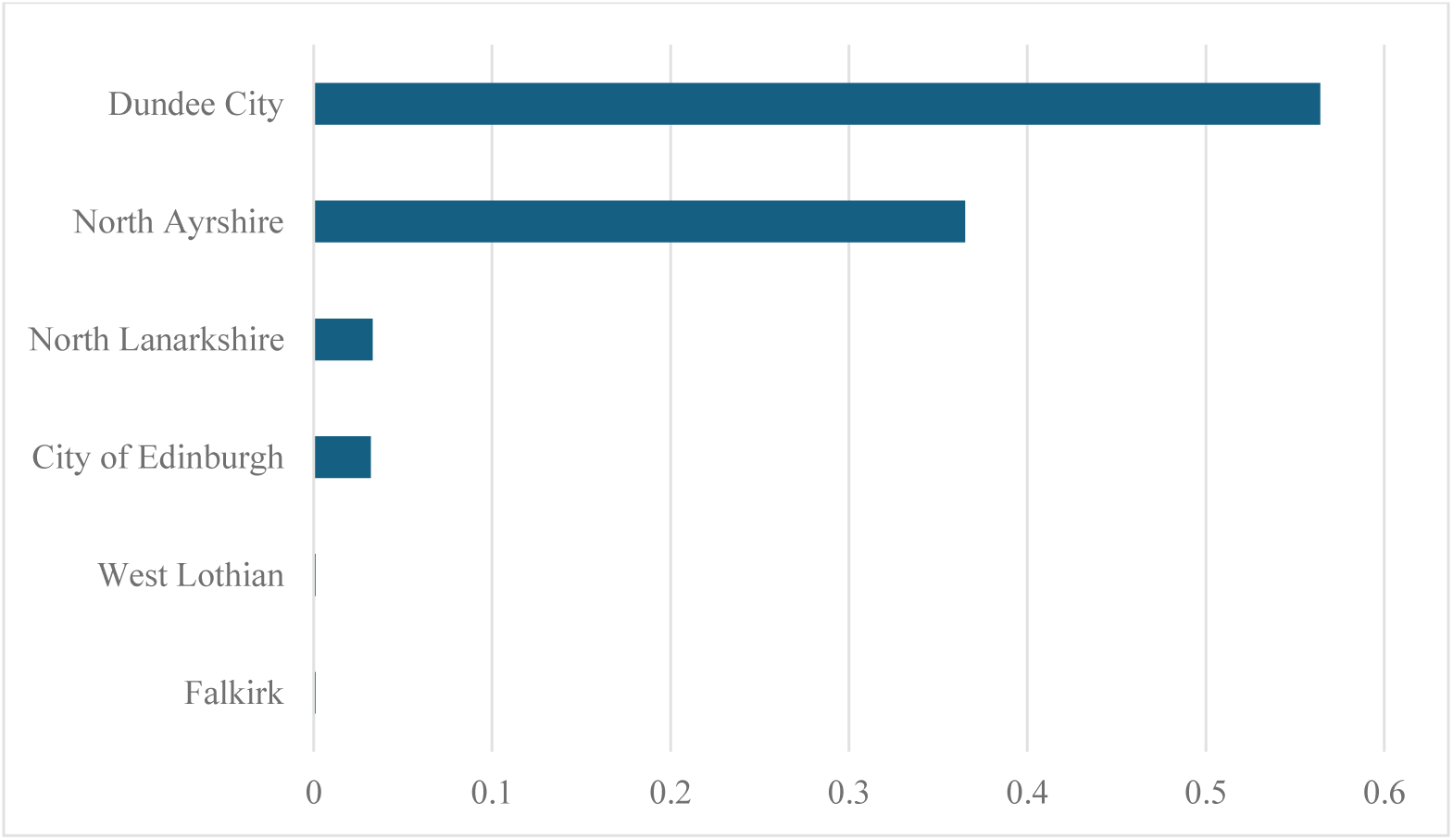
Optimal unit weights.

**Figure SC 07:**
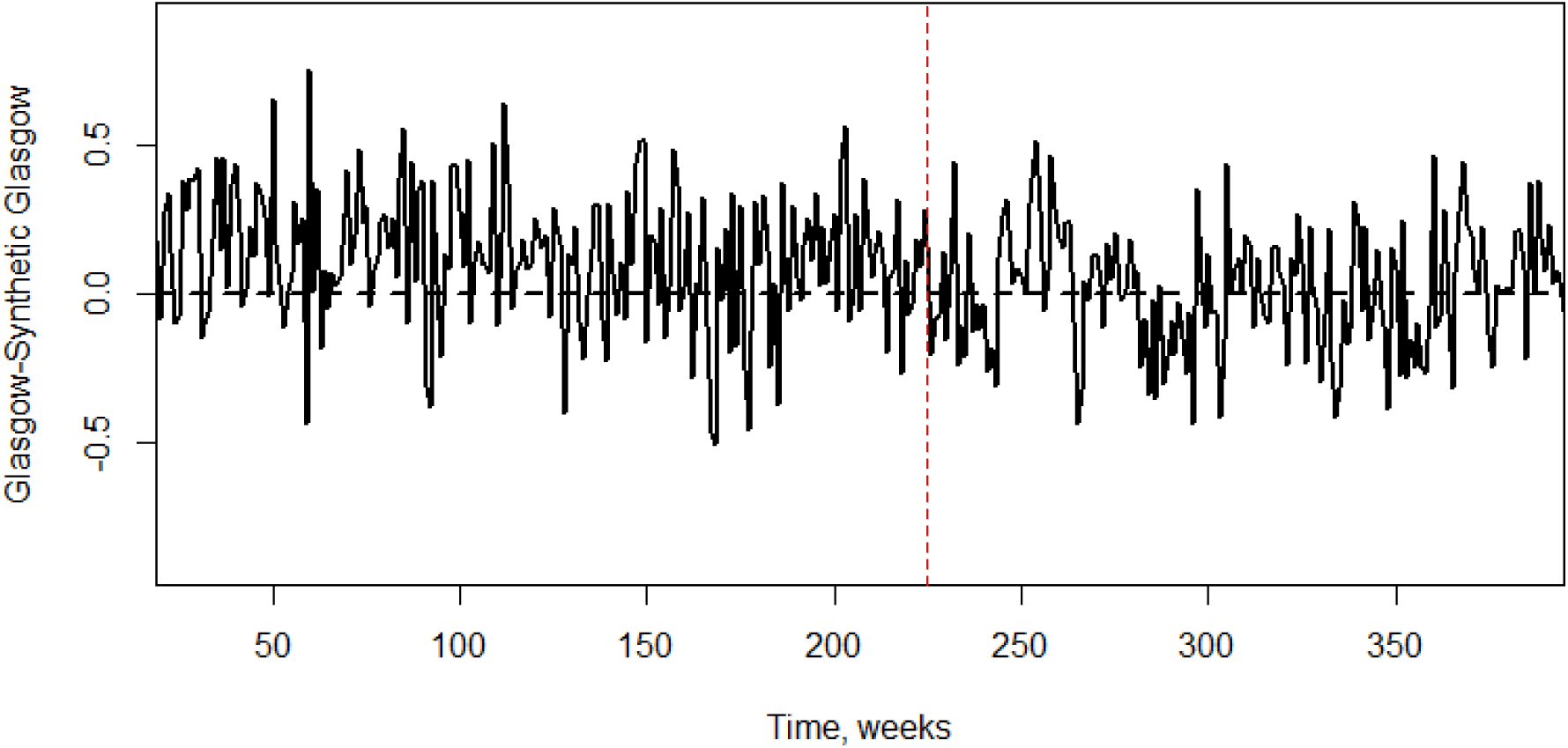
Gaps of Glasgow and synthetic Glasgow.

**Figure SC 08a:**
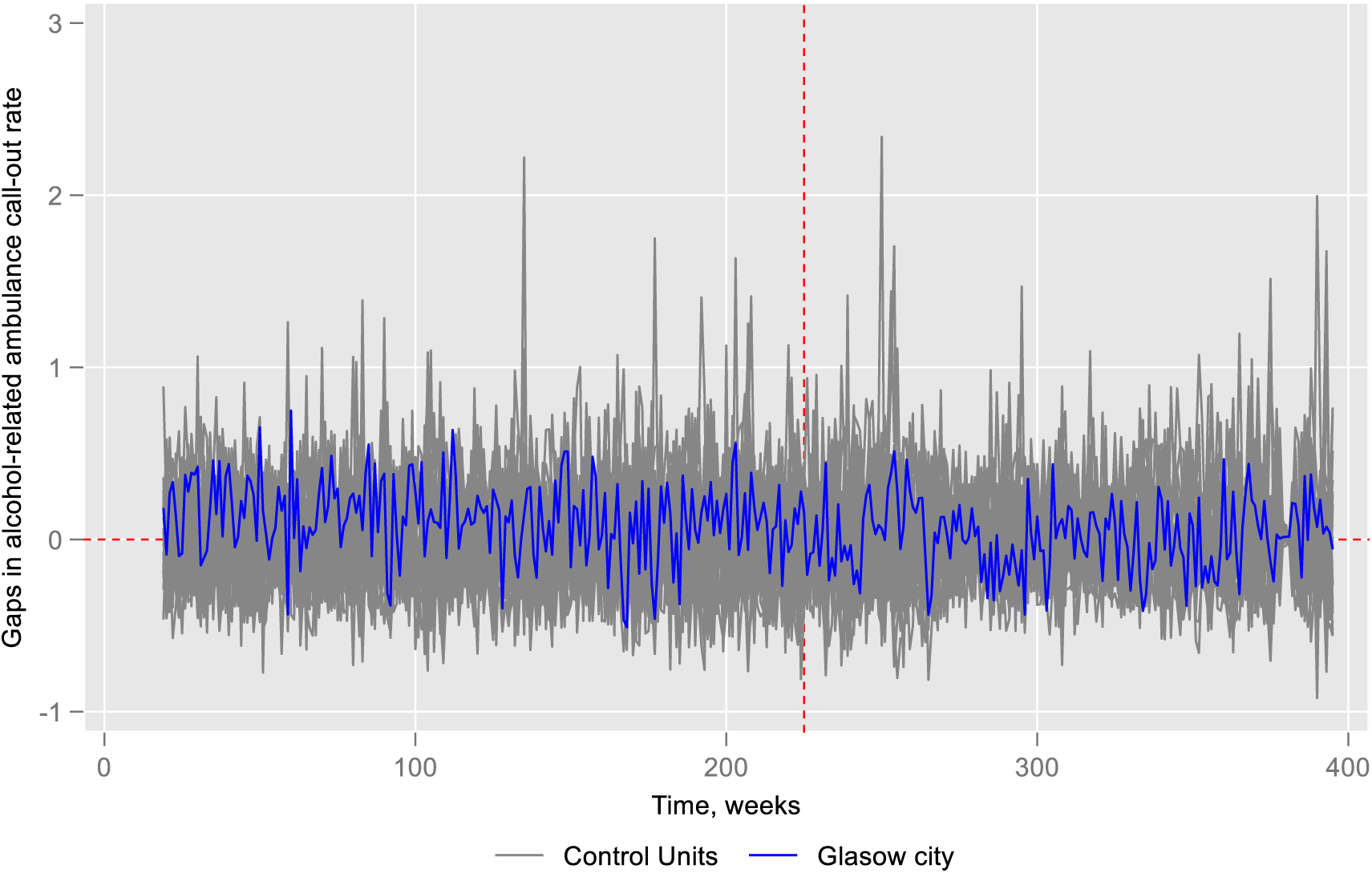
Placebo test.

**Figure SC 08b:**
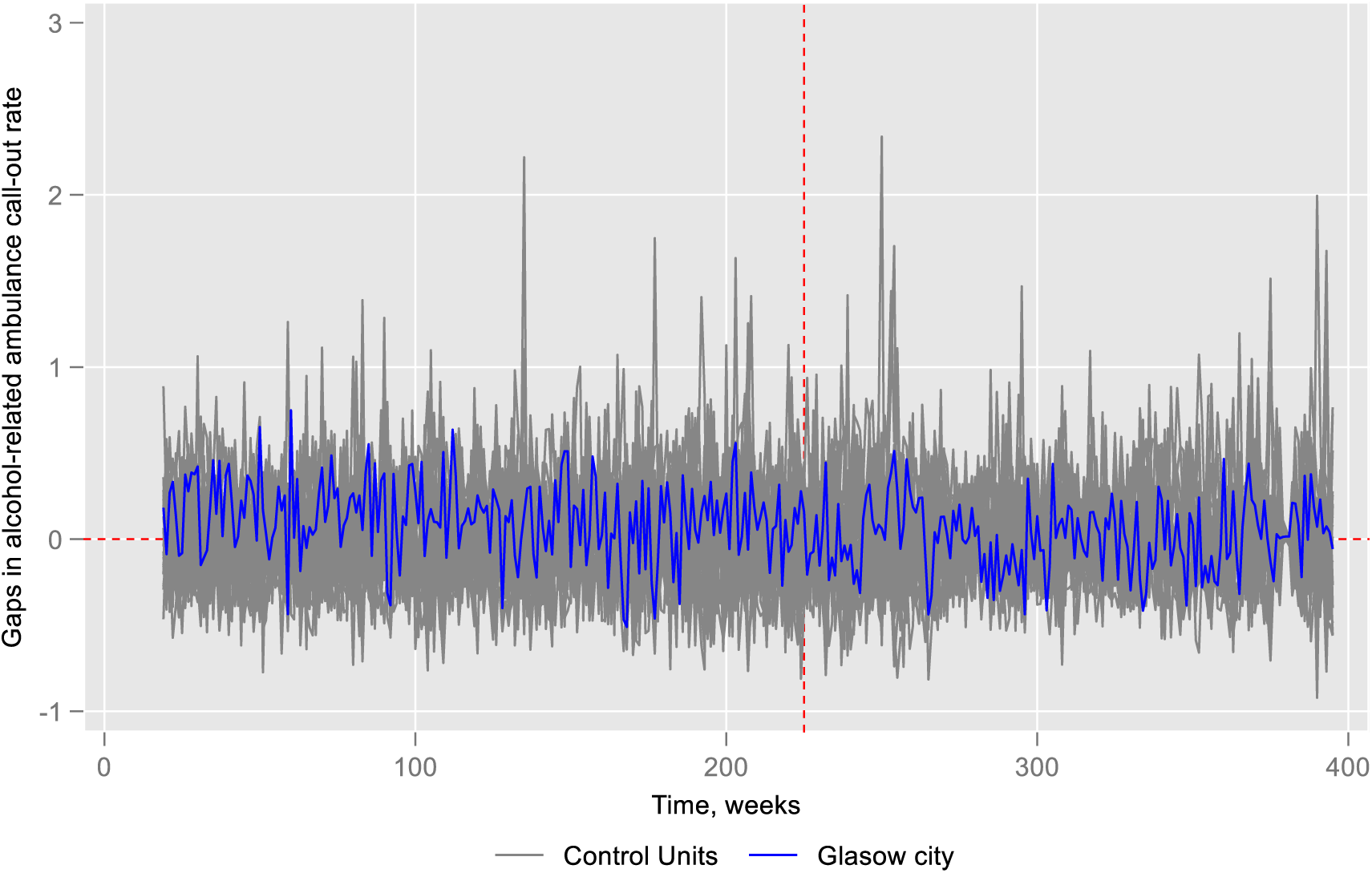
Placebo test excluding control cities MSPE>2 times Glasgow.

**Figure SC 08c:**
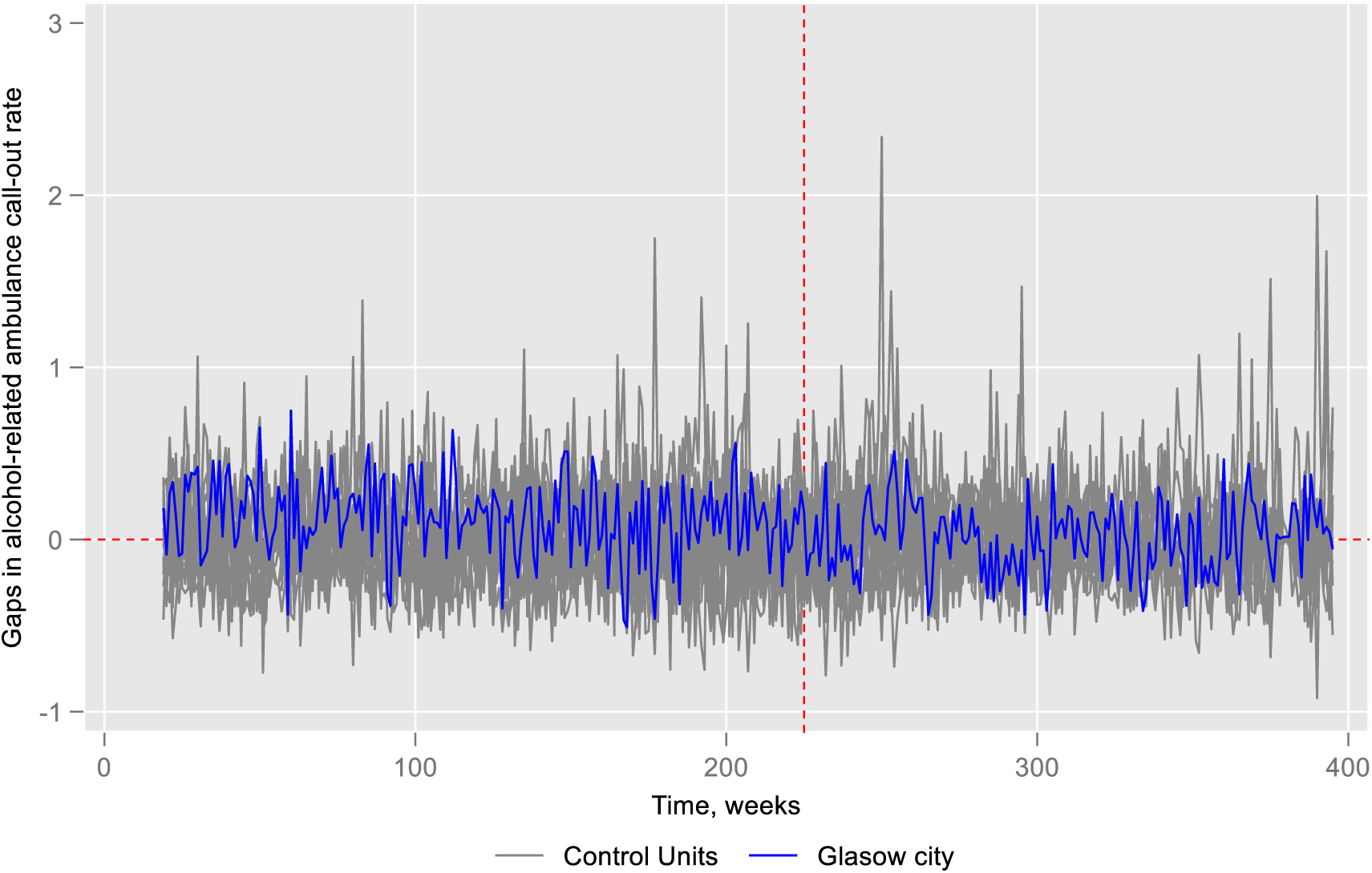
Placebo test excluding control cities if MSPE> Glasgow.

**Figure SC 09a:**
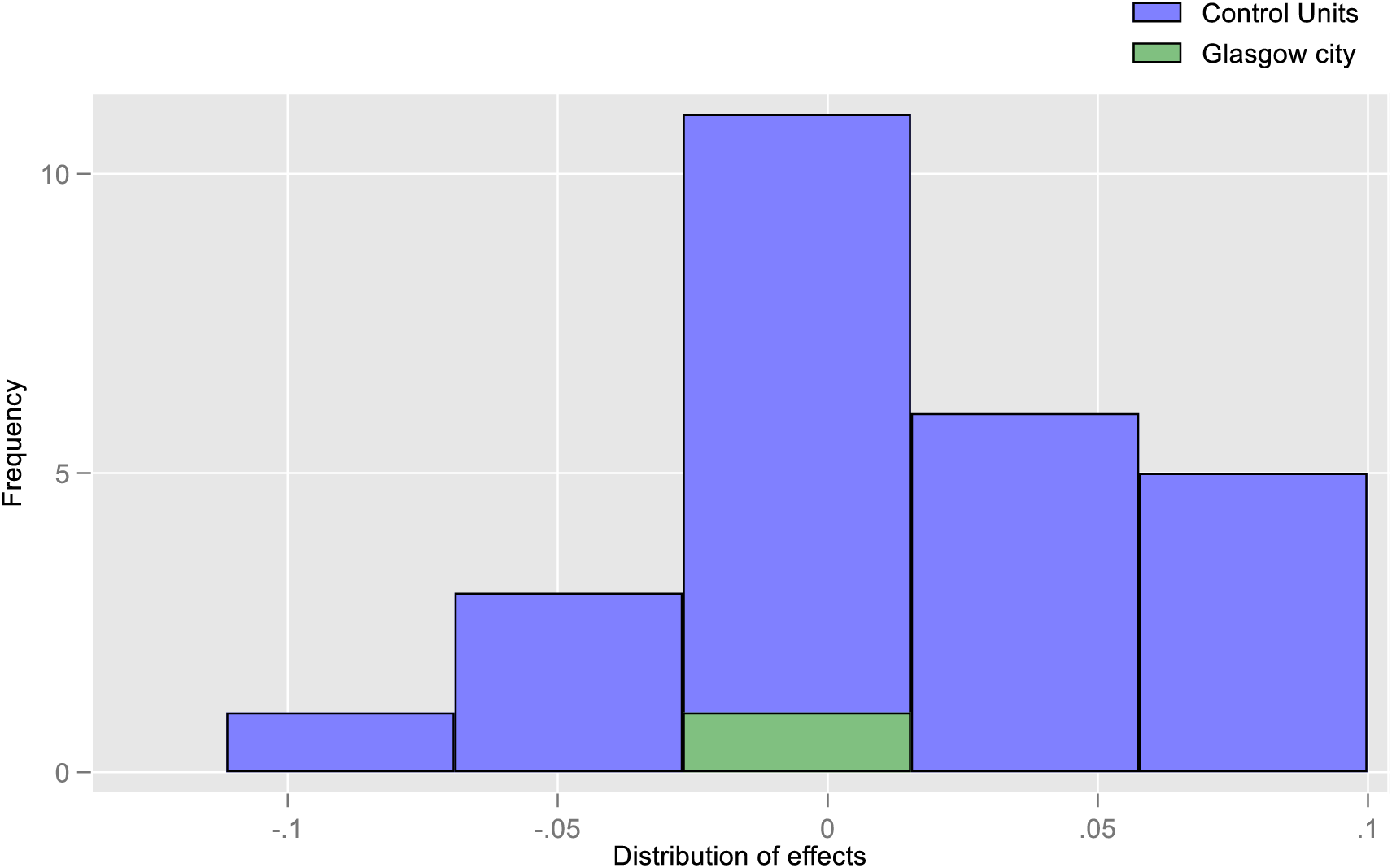
Effect size for placebo test.

**Figure SC 09b:**
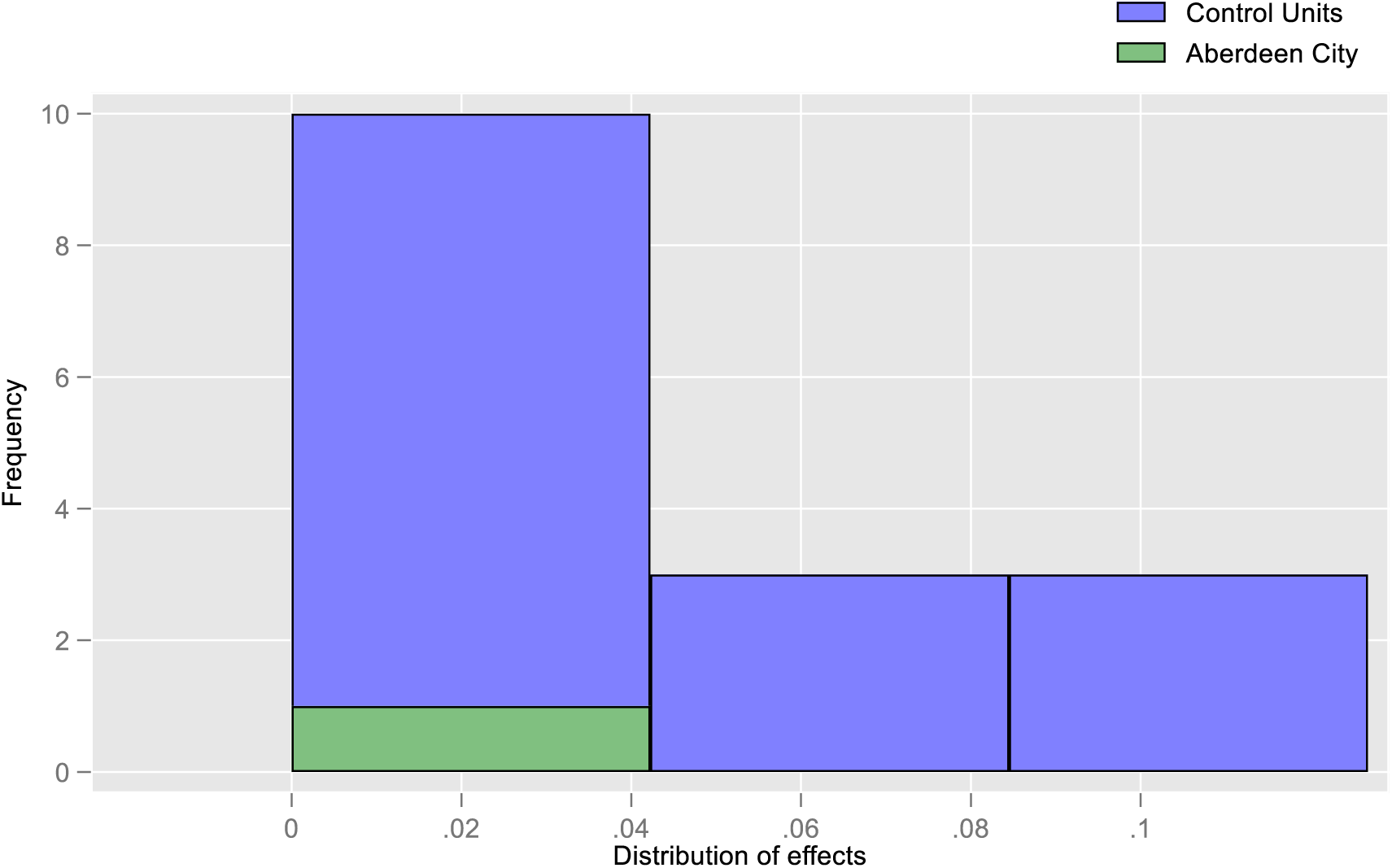
Effect size placebo test considering only positive effect size.

**Figure SC 10a:**
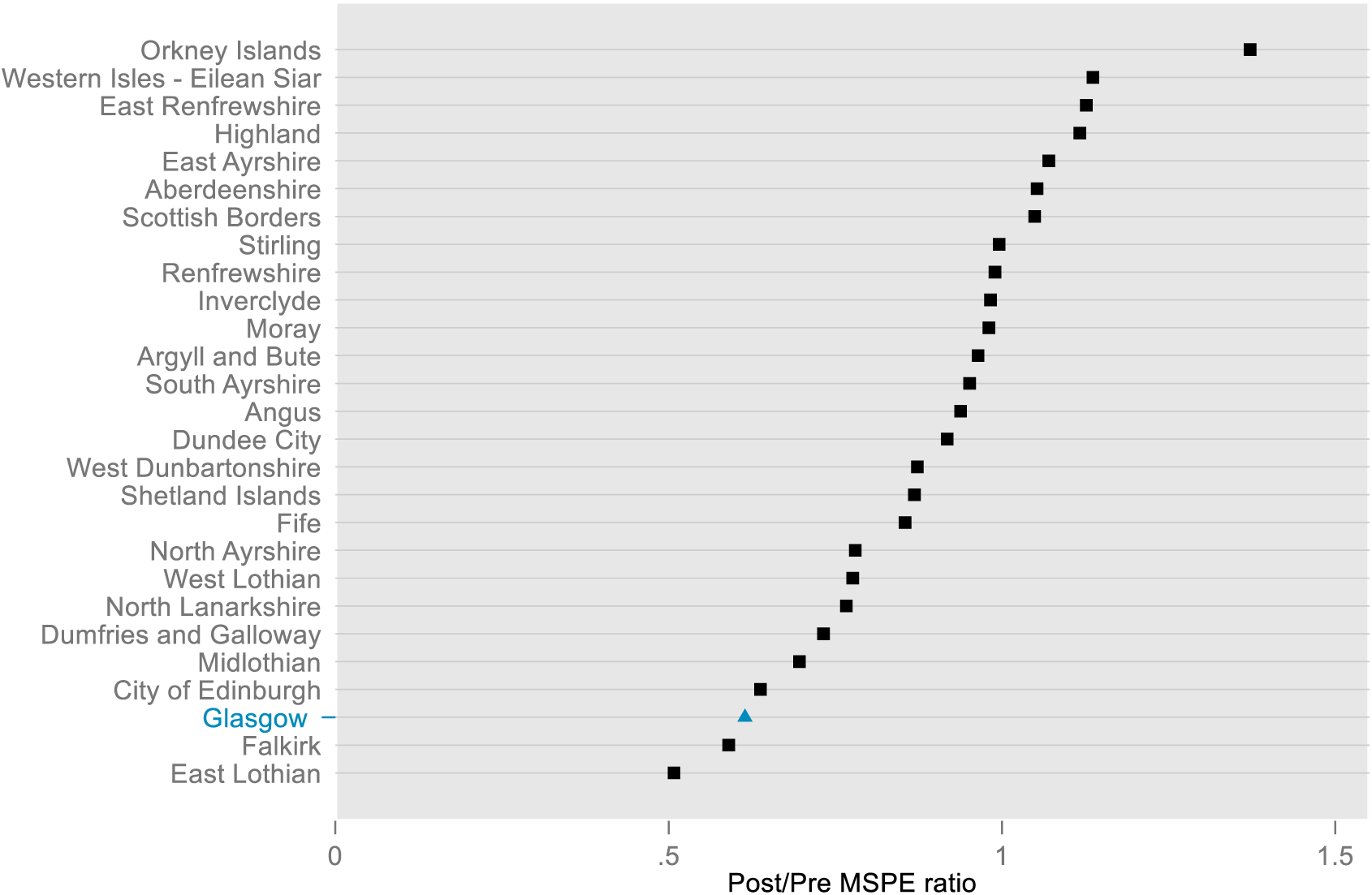
Post/Pre MSPE ratio across cities.

**Figure SC 10b:**
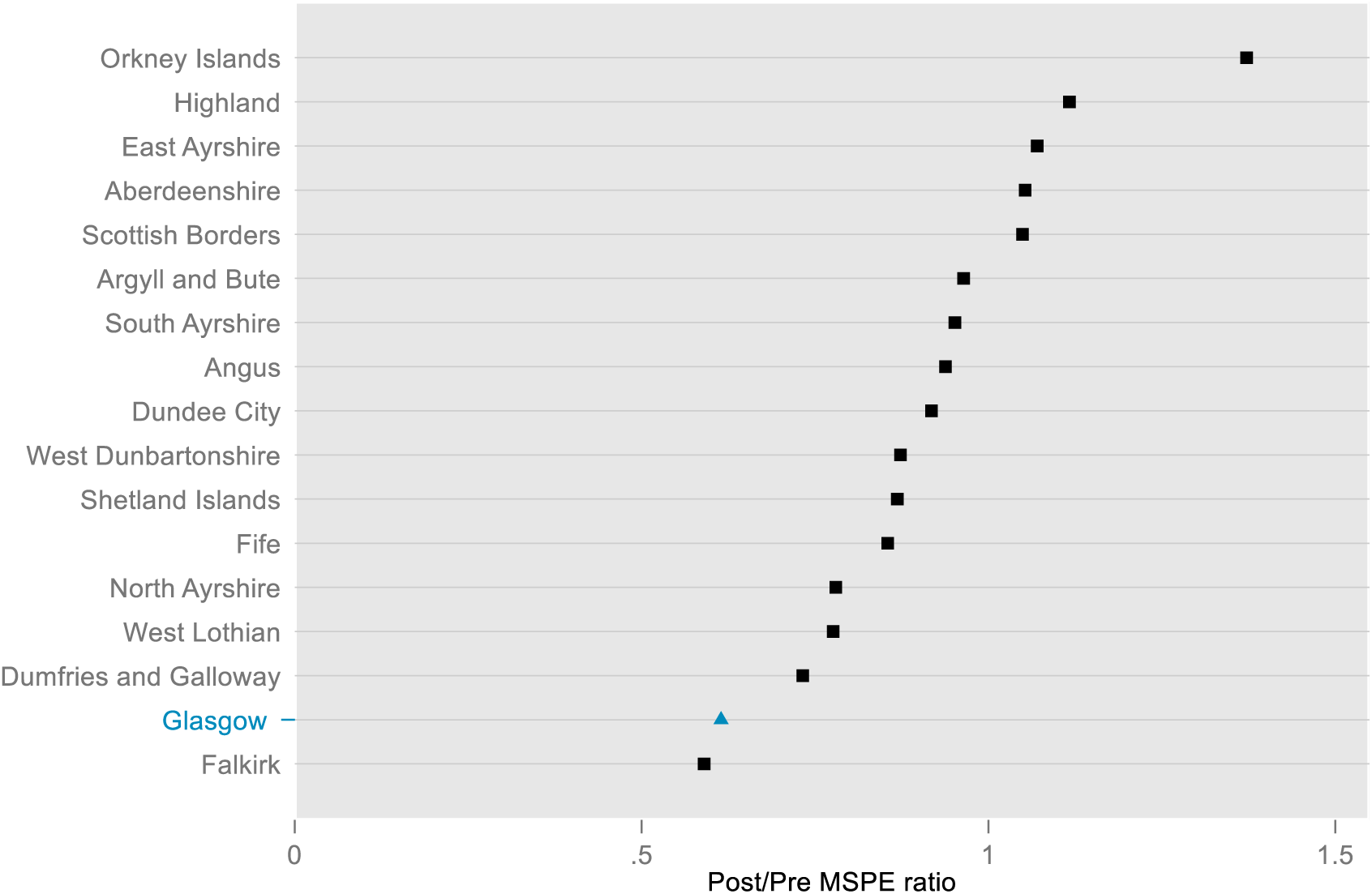
Post/Pre MSPE ratio across cities considering only positive effect size.

### Appendix 3.3. Reported crimes (Aberdeen)

#### Appendix 3.3.1. Model specification and validation test

For the reported crimes in Aberdeen, we identified three specifications (6, 7, and 9) that generated the lowest MSPEs (Table SC 5). Specification 6 had the lowest MSPE value among them. As a result, we selected specification 6 for the remainder of the analysis.

#### Appendix 3.3.2. Main analysis and sensitivity tests

We observed that the predictor means for Aberdeen were closely aligned with those of the synthetic Aberdeen with some exceptions (Table SC 6). Among the donor pool cities, Dundee, Edinburgh, West Lothian, and Falkirk were the most significant contributors to the creation of synthetic Aberdeen for reported crimes (Figure SC 11).

We estimated a positive average treatment effect (ATT) of 0·022, consistent with the direction of the effect sizes estimated using ARIMA based on difference-in-Differences approaches. However, the p-value from the placebo test for the synthetic control was 0·333 (Figure SC 15a). When we further modelled ARIMA with synthetic control, we also estimated a positive but statistically insignificant effect size of 0·016 (Table A18). Although the effect sizes across different approaches (ARIMA, synthetic control, and synthetic control + ARIMA) were homogeneous in terms of direction (positive), they were not always statistically significant. This underscores the uncertainty in the results and the need for caution in their interpretation.

**Table SC 5.** Validation test for synthetic control for reported crimes in Aberdeen.

| Specifications | Pre-intervention periods split between training and validation |  |  |
| --- | --- | --- | --- |
|  | Training/validation, 50/50 | Training/validation, 70/30 | Training/validation, 80/20 |
|  | MSPE* (Validation) | MSPE* (Validation) | MSPE* (Validation) |
| 1. (all pre-intervention outcome values and covariates) | 0.0772 | 0.05798 | 0.0570 |
| 2. (all pre-intervention outcome values only) | 0.0767 | 0.05798 | 0.0570 |
| 3. (pre-intervention outcome mean and covariates) | 0.0724 | 0.0639 | 0.0665 |
| 4. (last pre-intervention outcome value and covariates) | 0.1128 | 0.2336 | 0.0729 |
| 5. (pre-intervention outcome mean and last pre-intervention outcome value, and covariates) | 0.0722 | 0.0640 | 0.0682 |
| 6. (first three-fourths of the pre-intervention outcome values and covariates) | 0.0858 | 0.0714 | 0.0534 |
| 7. (last three-fourths of the pre-intervention outcome values and covariates) | 0.0707 | 0.0640 | 0.0693 |
| 8. (odd pre-intervention outcome values and covariates) | 0.0745 | 0.0647 | 0.0565 |
| 9. (even pre-intervention outcome values and covariates) | 0.0772 | 0.0570 | 0.0556 |

##### Aberdeen- Synthetic Aberdeen, Specification 6

**Table SC 6.** Covariate balance in the pre-intervention periods.

Aberdeen- Synthetic Aberdeen, Specification 6: **Table SC 6. Covariate balance in the pre-intervention periods**
| Covariates | Covariate weights | Reported crimes, predictor means |  |  |
| --- | --- | --- | --- | --- |
|  |  | Aberdeen | Synthetic Aberdeen | Average of other control cities |
| Number of on-premises alcohol outlets (population size adjusted) | 0·043 | 19·794 | 20·535 | 23·558 |
| Per capita gross disposable household income | 0·001 | 20125 | 17415 | 17773 |
| Pre-intervention outcome, period-19 | 0·026 | 0·695 | 0·702 | 0·655 |
| Pre-intervention outcome, period-20 | 0·000 | 0·825 | 0·976 | 0·534 |
| Pre-intervention outcome, period-21 | 0·004 | 0·825 | 0·773 | 0·749 |
| Pre-intervention outcome, period-22 | 0·103 | 1·086 | 0·895 | 0·677 |
| Pre-intervention outcome, period-23 | 0·004 | 0·608 | 0·720 | 0·641 |
| Pre-intervention outcome, period-24 | 0·000 | 0·825 | 0·926 | 0·695 |
| Pre-intervention outcome, period-25 | 0·040 | 0·565 | 0·679 | 0·764 |
| Pre-intervention outcome, period-26 | 0·019 | 0·825 | 0·902 | 0·644 |
| Pre-intervention outcome, period-27 | 0·030 | 0·738 | 0·700 | 0·674 |
| Pre-intervention outcome, period-28 | 0·001 | 0·434 | 0·733 | 0·574 |
| Pre-intervention outcome, period-29 | 0·081 | 0·695 | 0·667 | 0·598 |
| Pre-intervention outcome, period-30 | 0·001 | 0·739 | 0·884 | 0·642 |
| Pre-intervention outcome, period-31 | 0·020 | 1·260 | 1·086 | 0·905 |
| Pre-intervention outcome, period-32 | 0·017 | 0·565 | 0·966 | 0·765 |
| Pre-intervention outcome, period-33 | 0·013 | 0·695 | 0·872 | 0·777 |
| Pre-intervention outcome, period-34 | 0·047 | 0·826 | 0·783 | 0·595 |
| Pre-intervention outcome, period-35 | 0·044 | 1·478 | 0·852 | 0·716 |
| Pre-intervention outcome, period-36 | 0·062 | 0·826 | 0·691 | 0·642 |
| Pre-intervention outcome, period-37 | 0·005 | 0·435 | 0·712 | 0·668 |
| Pre-intervention outcome, period-38 | 0·047 | 0·869 | 1·028 | 0·631 |
| Pre-intervention outcome, period-39 | 0·116 | 1·043 | 0·912 | 0·663 |
| Pre-intervention outcome, period-40 | 0·078 | 0·696 | 0·677 | 0·545 |
| Pre-intervention outcome, period-41 | 0·094 | 1·261 | 0·962 | 0·544 |
| Pre-intervention outcome, period-42 | 0·097 | 0·870 | 0·725 | 0·724 |
| Pre-intervention outcome, period-43 | 0·007 | 1·044 | 0·632 | 0·592 |

**Figure SC 11:**
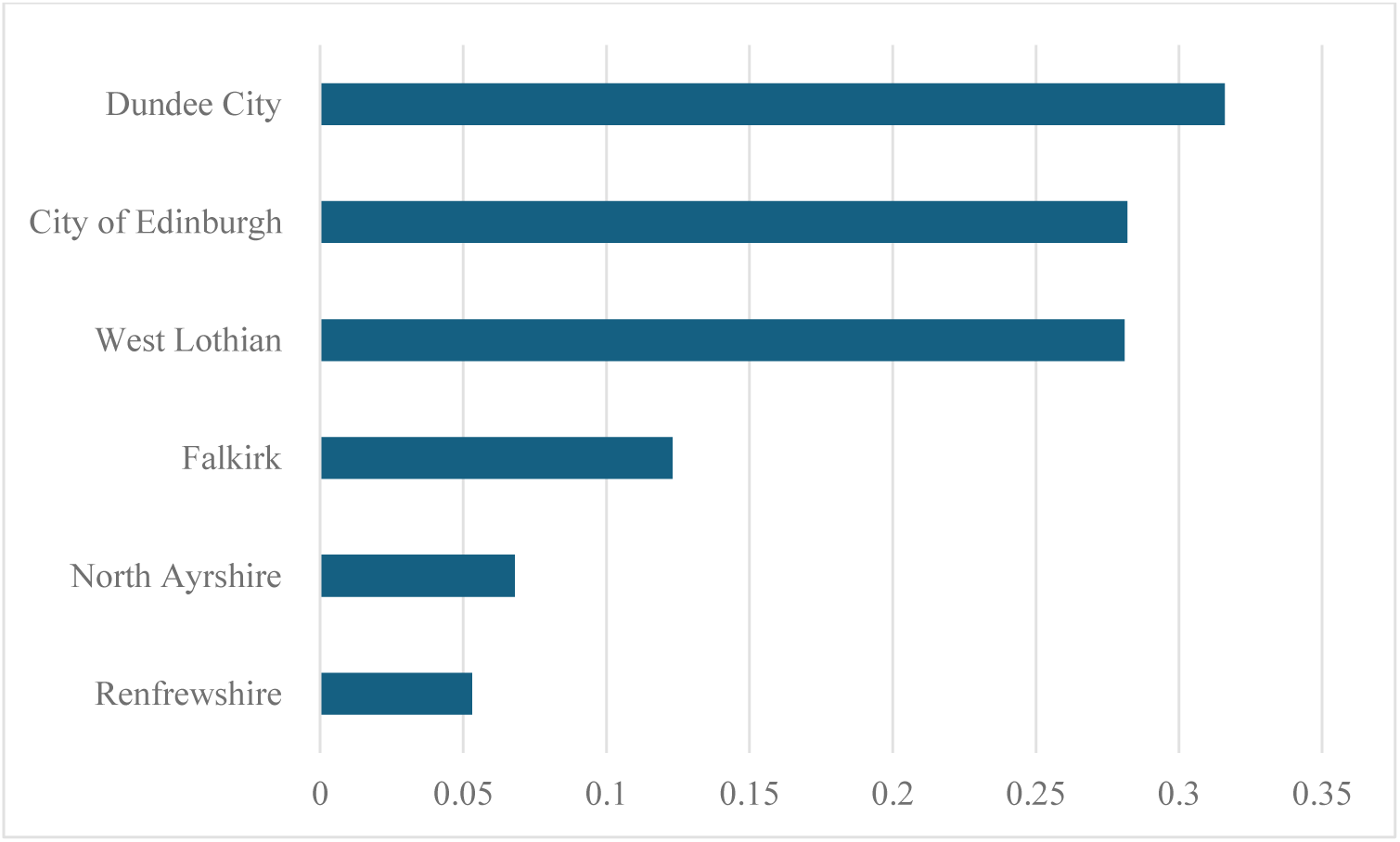
Optimal unit weights.

**Figure SC 12:**
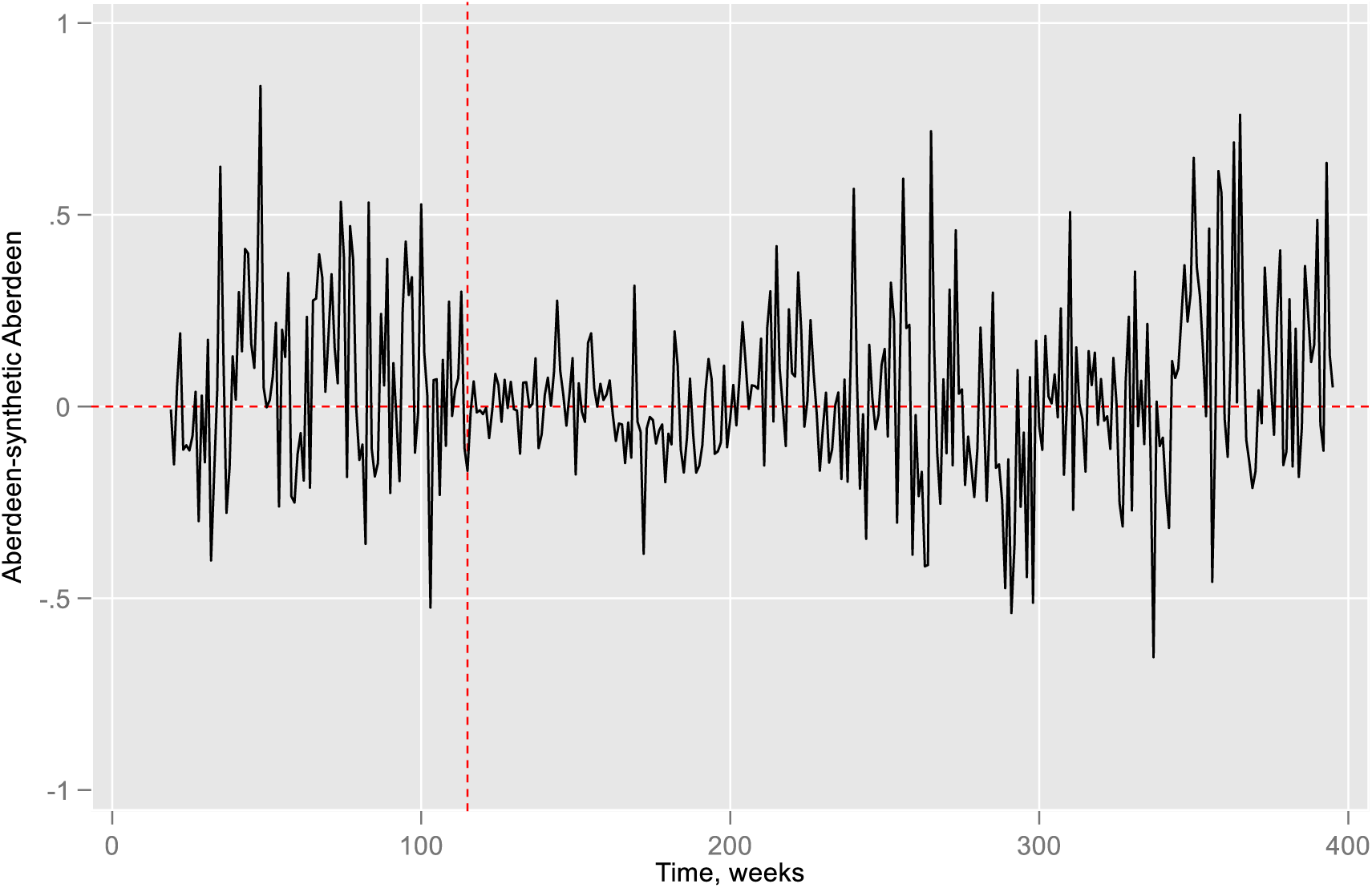
Gaps of Aberdeen and synthetic Aberdeen.

**Figure SC 13a:**
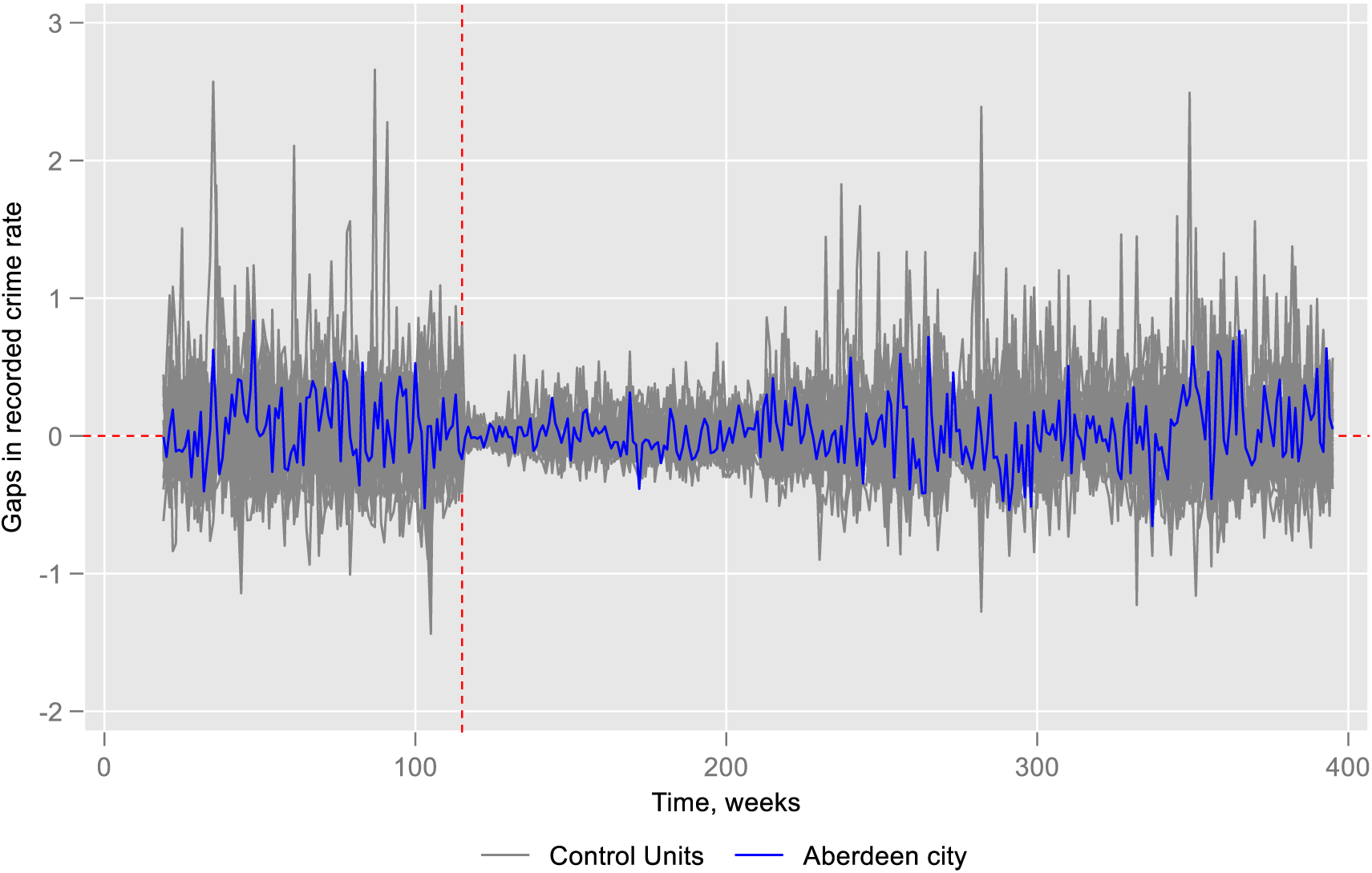
Placebo test.

**Figure SC 13b:**
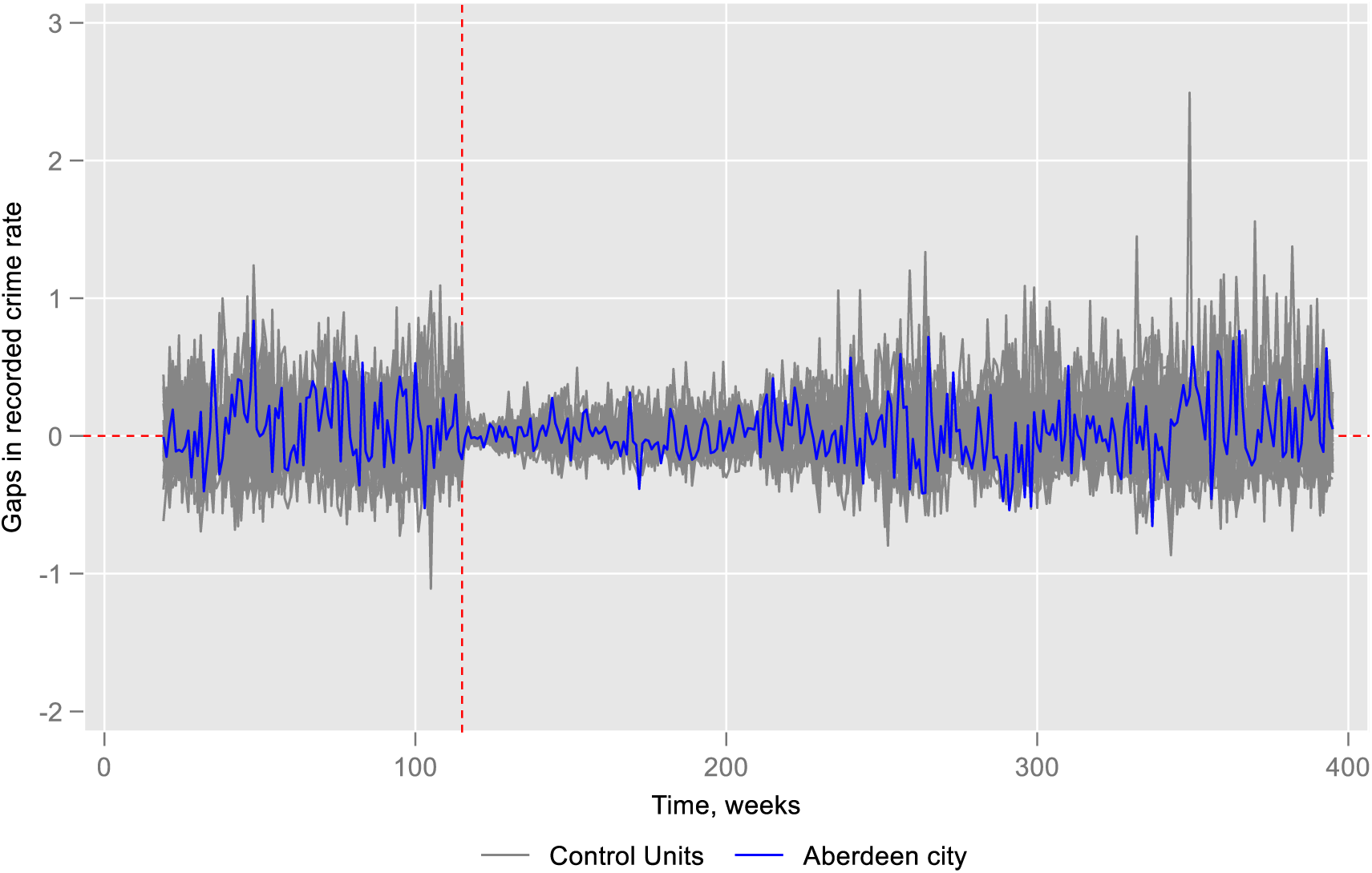
Placebo test excluding control cities MSPE>2 times Aberdeen.

**Figure SC 13c:**
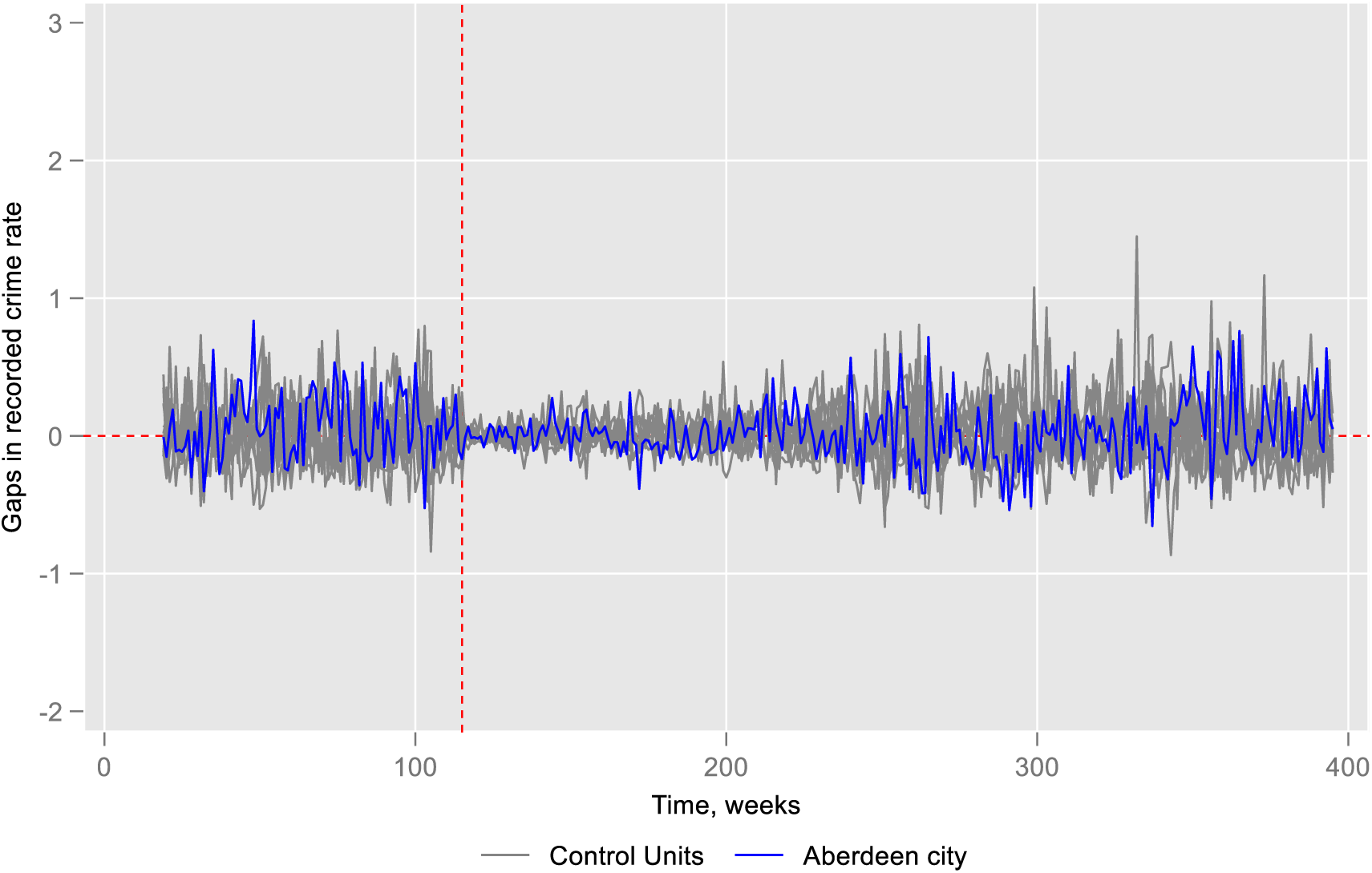
Placebo test excluding control cities if MSPE> Aberdeen.

**Figure SC 14a:**
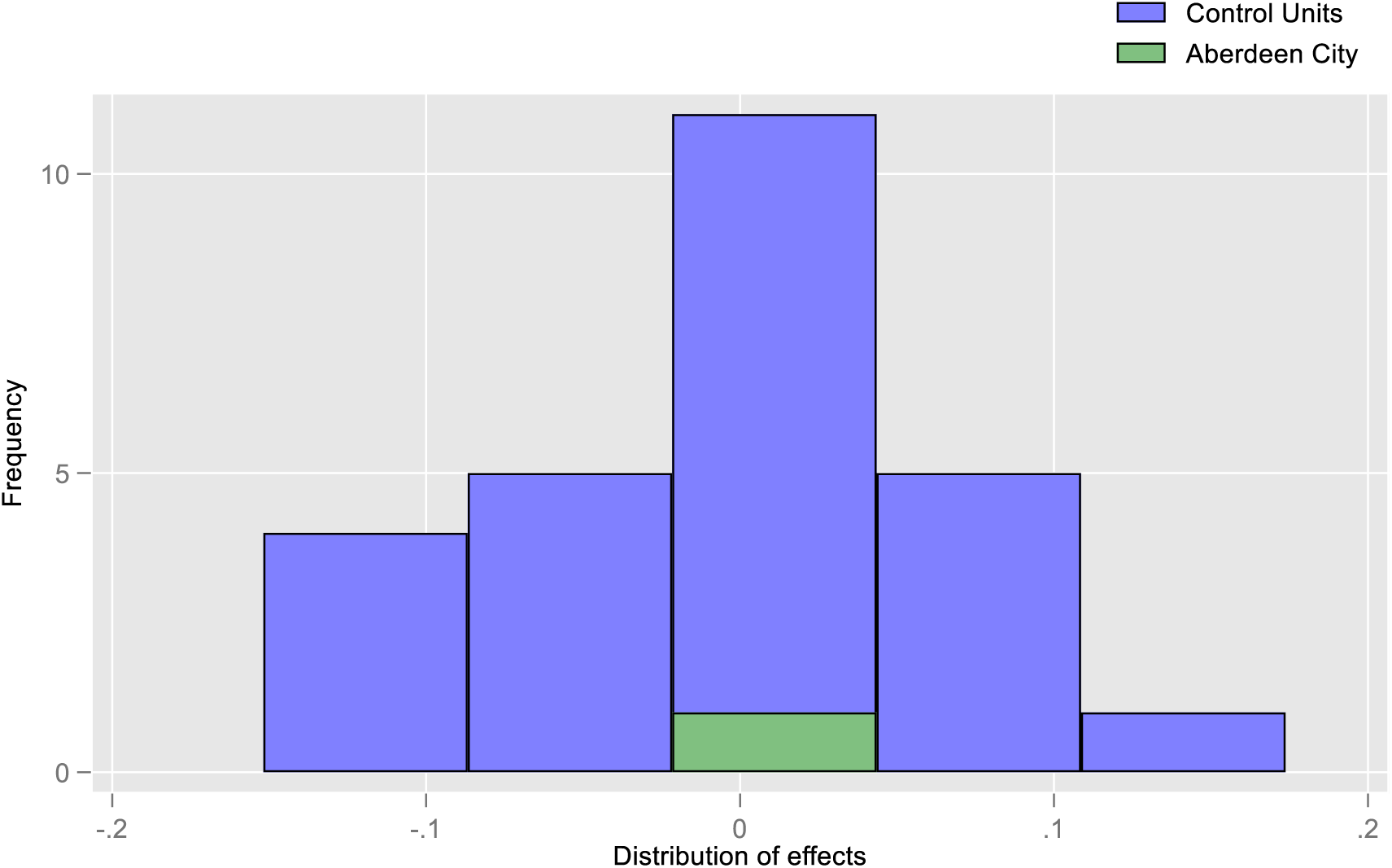
Effect size for placebo test.

**Figure SC 14b:**
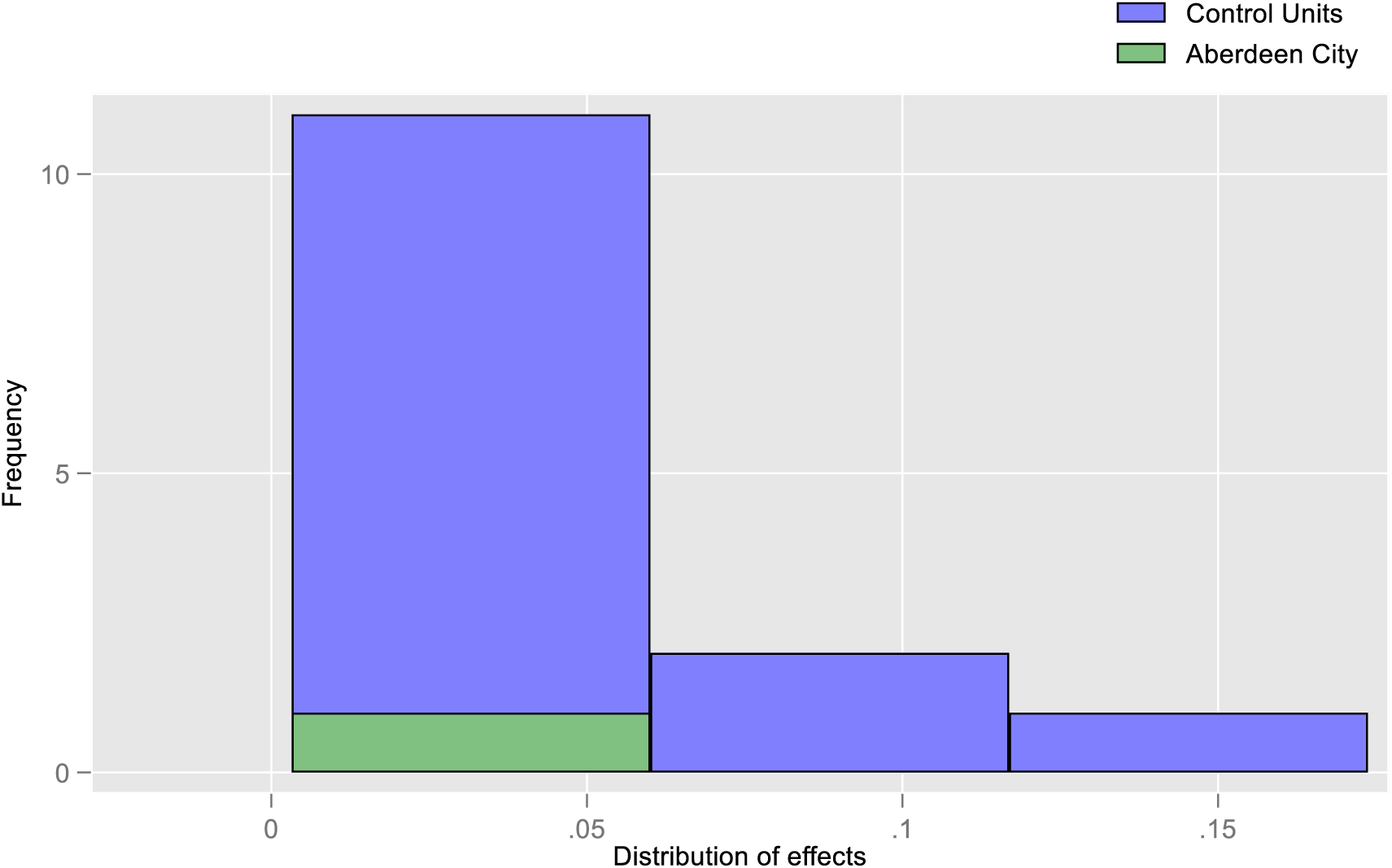
Effect size placebo test considering only positive effect size.

**Figure SC 15a:**
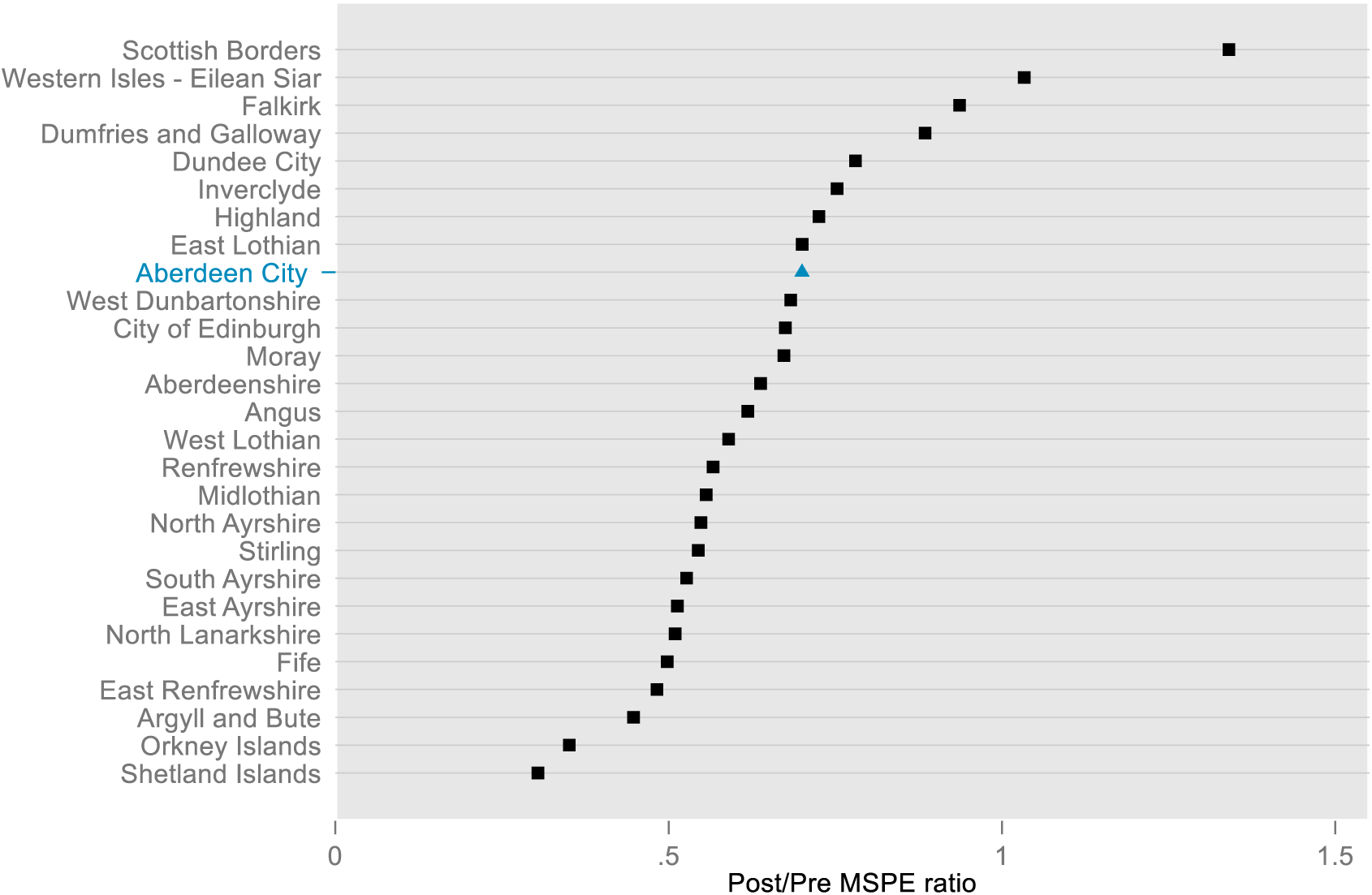
Post/Pre MSPE ratio across cities.

**Figure SC 15b:**
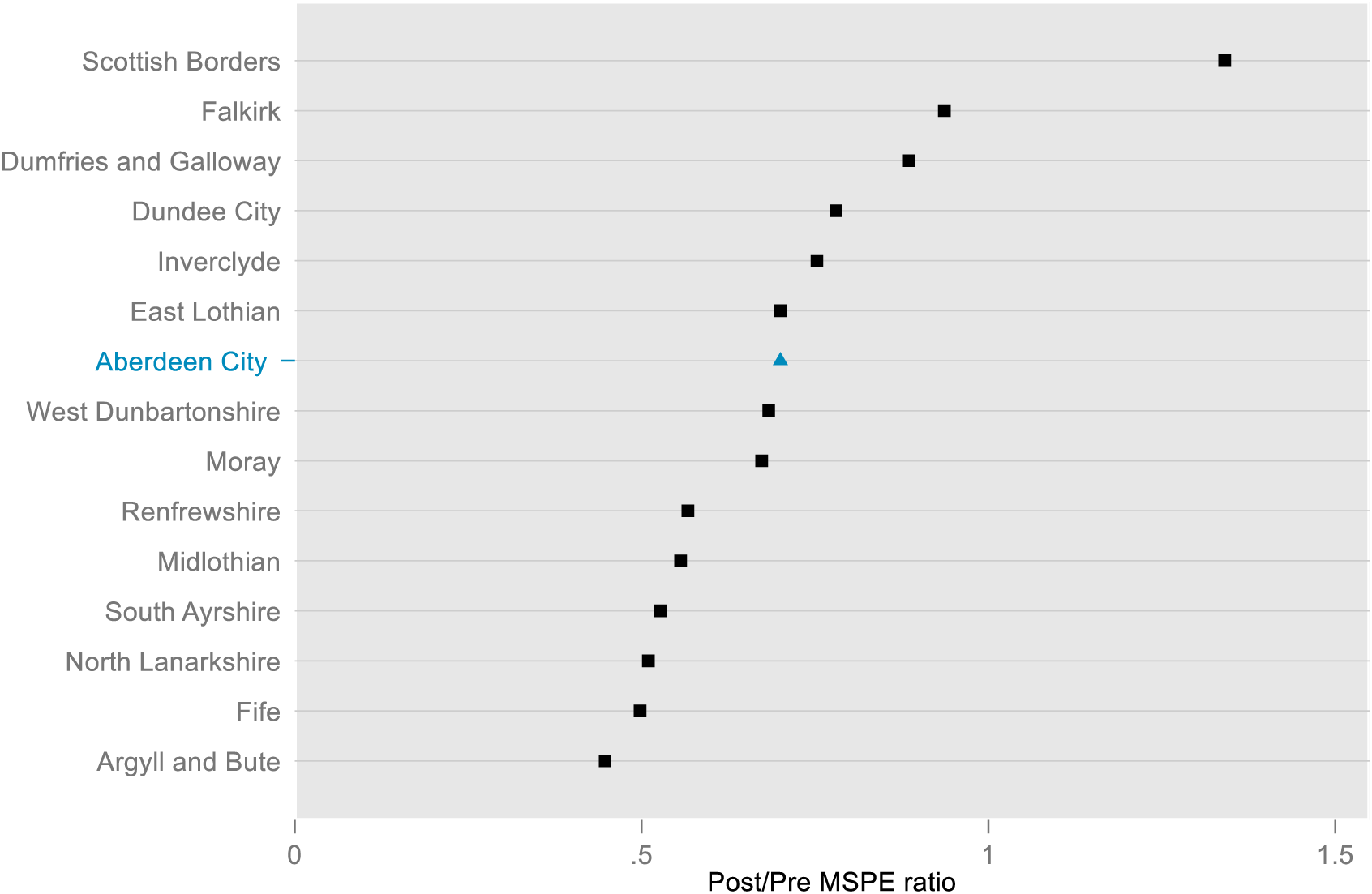
Post/Pre MSPE ratio across cities considering only positive effect size.

### Appendix 3.4. Reported crimes (Glasgow)

#### Appendix 3.4.1. Model specification and validation test

For the reported crimes in Glasgow, we identified specifications 6, 7, and 9 that generated the lowest MSPEs (Table SC 7). Among them specification 7 a had the lowest value of MSPE. As a result, we selected specification 7 for the remainder of the analysis.

#### Appendix 3.4.2. Main analysis and sensitivity tests

We observed that the predictor means for Glasgow were closely aligned with those of the synthetic Glasgow with some exceptions (Table SC 8). Among the donor pool cities, Edinburgh, Dundee, and Falkirk were the most significant contributors to the creation of synthetic Glasgow for reported crimes (Figure SC 11).

We estimated a positive average treatment effect (ATT) of 0·148. However, the effect sizes estimated using ARIMA based on difference-in-Differences models were negative and insignificant, indicating a discrepancy between the synthetic control and ARIMA approaches. Additionally, the p-value from the placebo test was 0·519, indicating a high level of uncertainty (Figure SC 20a). When we further modelled ARIMA with synthetic control, we estimated positive and statistically insignificant effect size (Table A18). Despite the different nature of p-values (inferential for ARIMA and frequentist for Synthetic) the different results from different approaches in terms of direction and significance weakened our confidence in interpreting the overall impact of the policy change in Glasgow, making these results unconclusive.

**Table SC 7.** Validation test for synthetic control for reported crimes in Glasgow.

| Specifications | Pre-intervention periods split between training and validation |  |  |
| --- | --- | --- | --- |
|  | Training/validation, 50/50 | Training/validation, 70/30 | Training/validation, 80/20 |
|  | MSPE* (Validation) | MSPE* (Validation) | MSPE* (Validation) |
| 1. (all pre-intervention outcome values and covariates) | 0.0765 | 0.05166 | 0.0473 |
| 2. (all pre-intervention outcome values only) | 0.0765 | 0.05133 | 0.0473 |
| 3. (pre-intervention outcome mean and covariates) | 0.0853 | 0.0819 | 0.0632 |
| 4. (last pre-intervention outcome value and covariates) | 0.1298 | 0.0670 | 0.0598 |
| 5. (pre-intervention outcome mean and last pre-intervention outcome value, and covariates) | 0.0930 | 0.0643 | 0.0584 |
| 6. (first three-fourths of the pre-intervention outcome values and covariates) | 0.0745 | 0.0513 | 0.0473 |
| 7. (last three-fourths of the pre-intervention outcome values and covariates) | 0.0724 | 0.0513 | 0.0473 |
| 8. (odd pre-intervention outcome values and covariates) | 0.0766 | 0.0517 | 0.0473 |
| 9. (even pre-intervention outcome values and covariates) | 0.0724 | 0.0513 | 0.0475 |

##### Glasgow- Synthetic Glasgow, Specification 7

**Table SC 8.** Covariates balance in the pre-intervention periods.

Glasgow- Synthetic Glasgow, Specification 7: **Table SC 8. Covariates balance in the pre-intervention periods**
| Covariates | Covariate weights | Reported crimes, predictor means |
| --- | --- | --- |

|  |  | **Glasgow** | **Synthetic**<br>**Glasgow** | **Average**<br>**of other**<br>**control**<br>**cities** |
| --- | --- | --- | --- | --- |
| Number of on-premises alcohol outlets<br>(population size adjusted) | 0·016 | 21·371 | 23·402 | 23·529 |
| Per capita gross disposable household<br>income | 0·008 | 15852 | 18900 | 18246 |
| Pre-intervention outcome, period-169 | 0·019 | 0·733 | 0·835 | 0·748 |
| Pre-intervention outcome, period-170 | 0·017 | 0·701 | 0·720 | 0·456 |
| Pre-intervention outcome, period-171 | 0·012 | 0·637 | 0·701 | 0·413 |
| Pre-intervention outcome, period-172 | 0·011 | 0·987 | 1·085 | 0·539 |
| Pre-intervention outcome, period-173 | 0·017 | 1·002 | 0·779 | 0·543 |
| Pre-intervention outcome, period-174 | 0·015 | 1·114 | 0·676 | 0·519 |
| Pre-intervention outcome, period-175 | 0·005 | 0·891 | 0·411 | 0·421 |
| Pre-intervention outcome, period-176 | 0·026 | 0·954 | 0·811 | 0·516 |
| Pre-intervention outcome, period-177 | 0·016 | 0·922 | 0·964 | 0·567 |
| Pre-intervention outcome, period-178 | 0·014 | 0·763 | 0·398 | 0·501 |
| Pre-intervention outcome, period-179 | 0·021 | 0·938 | 0·681 | 0·495 |
| Pre-intervention outcome, period-180 | 0·020 | 0·826 | 0·589 | 0·470 |
| Pre-intervention outcome, period-181 | 0·014 | 0·826 | 0·618 | 0·468 |
| Pre-intervention outcome, period-182 | 0·014 | 1·080 | 1·178 | 0·795 |
| Pre-intervention outcome, period-183 | 0·006 | 0·889 | 0·545 | 0·525 |
| Pre-intervention outcome, period-184 | 0·017 | 0·826 | 0·798 | 0·556 |
| Pre-intervention outcome, period-185 | 0·020 | 0·730 | 0·636 | 0·458 |
| Pre-intervention outcome, period-186 | 0·015 | 0·746 | 0·855 | 0·562 |
| Pre-intervention outcome, period-187 | 0·016 | 0·793 | 0·717 | 0·505 |
| Pre-intervention outcome, period-188 | 0·014 | 0·809 | 0·622 | 0·523 |
| Pre-intervention outcome, period-189 | 0·017 | 0·682 | 0·920 | 0·565 |
| Pre-intervention outcome, period-190 | 0·006 | 0·682 | 0·857 | 0·528 |
| Pre-intervention outcome, period-191 | 0·024 | 1·094 | 0·723 | 0·597 |
| Pre-intervention outcome, period-192 | 0·018 | 0·824 | 0·690 | 0·585 |
| Pre-intervention outcome, period-193 | 0·020 | 0·713 | 0·535 | 0·401 |
| Pre-intervention outcome, period-194 | 0·018 | 0·681 | 0·573 | 0·419 |
| Pre-intervention outcome, period-195 | 0·016 | 0·776 | 0·804 | 0·539 |
| Pre-intervention outcome, period-196 | 0·023 | 0·792 | 0·969 | 0·543 |
| Pre-intervention outcome, period-197 | 0·017 | 0·586 | 0·516 | 0·566 |
| Pre-intervention outcome, period-198 | 0·027 | 0·886 | 0·575 | 0·629 |
| Pre-intervention outcome, period-199 | 0·020 | 0·918 | 0·950 | 0·591 |
| Pre-intervention outcome, period-200 | 0·018 | 1·013 | 0·962 | 0·528 |
| Pre-intervention outcome, period-201 | 0·017 | 0·823 | 0·822 | 0·475 |
| Pre-intervention outcome, period-202 | 0·013 | 0·854 | 0·694 | 0·552 |
| Pre-intervention outcome, period-203 | 0·018 | 0·522 | 0·468 | 0·370 |
| Pre-intervention outcome, period-204 | 0·028 | 1·360 | 0·913 | 0·616 |
| Pre-intervention outcome, period-205 | 0·022 | 0·711 | 0·666 | 0·459 |
| Pre-intervention outcome, period-206 | 0·013 | 0·616 | 0·794 | 0·563 |
| Pre-intervention outcome, period-207 | 0·020 | 1·043 | 0·720 | 0·595 |
| Pre-intervention outcome, period-208 | 0·018 | 0·964 | 0·702 | 0·594 |
| Pre-intervention outcome, period-209 | 0·020 | 0·521 | 0·469 | 0·267 |
| Pre-intervention outcome, period-210 | 0·021 | 0·805 | 0·447 | 0·344 |
| Pre-intervention outcome, period-211 | 0·036 | 0·758 | 0·527 | 0·341 |
| Pre-intervention outcome, period-212 | 0·013 | 1·074 | 0·562 | 0·413 |
| Pre-intervention outcome, period-213 | 0·016 | 0·932 | 0·950 | 0·529 |
| Pre-intervention outcome, period-214 | 0·016 | 0·774 | 0·622 | 0·557 |
| Pre-intervention outcome, period-215 | 0·011 | 0·821 | 0·563 | 0·410 |
| Pre-intervention outcome, period-216 | 0·022 | 0·868 | 0·726 | 0·476 |
| Pre-intervention outcome, period-217 | 0·012 | 1·026 | 0·608 | 0·496 |
| Pre-intervention outcome, period-218 | 0·020 | 0·884 | 0·617 | 0·559 |
| Pre-intervention outcome, period-219 | 0·021 | 0·758 | 0·512 | 0·483 |
| Pre-intervention outcome, period-220 | 0·018 | 0·852 | 0·630 | 0·480 |
| Pre-intervention outcome, period-221 | 0·018 | 0·947 | 0·900 | 0·507 |
| Pre-intervention outcome, period-222 | 0·019 | 0·458 | 0·597 | 0·521 |
| Pre-intervention outcome, period-223 | 0·014 | 0·647 | 0·422 | 0·401 |
| Pre-intervention outcome, period-224 | 0·017 | 0·883 | 0·706 | 0·422 |

**Figure SC 16:**
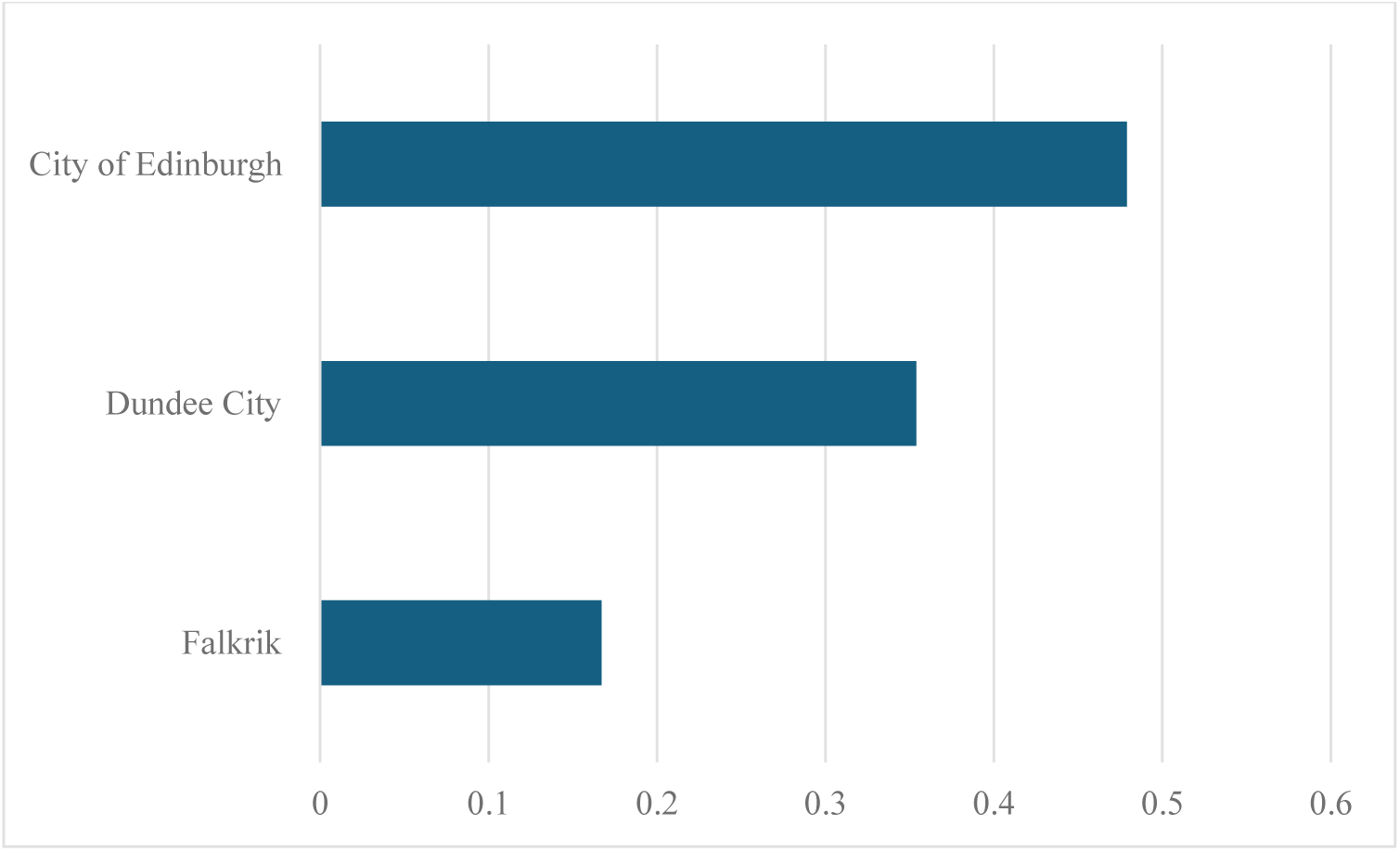
Optimal unit weights.

**Figure SC 17:**
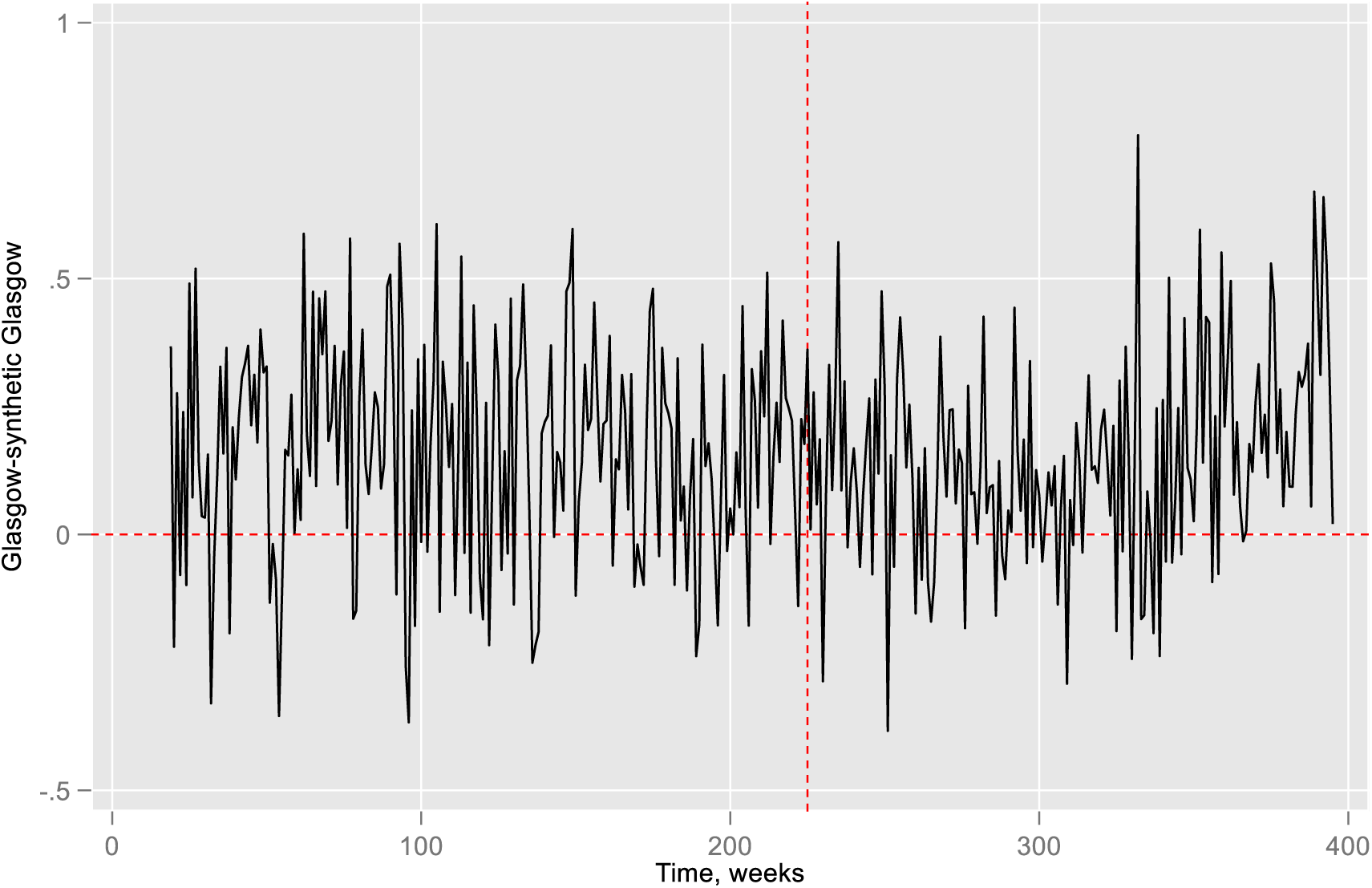
Gaps of Glasgow and synthetic Glasgow.

**Figure SC 18a:**
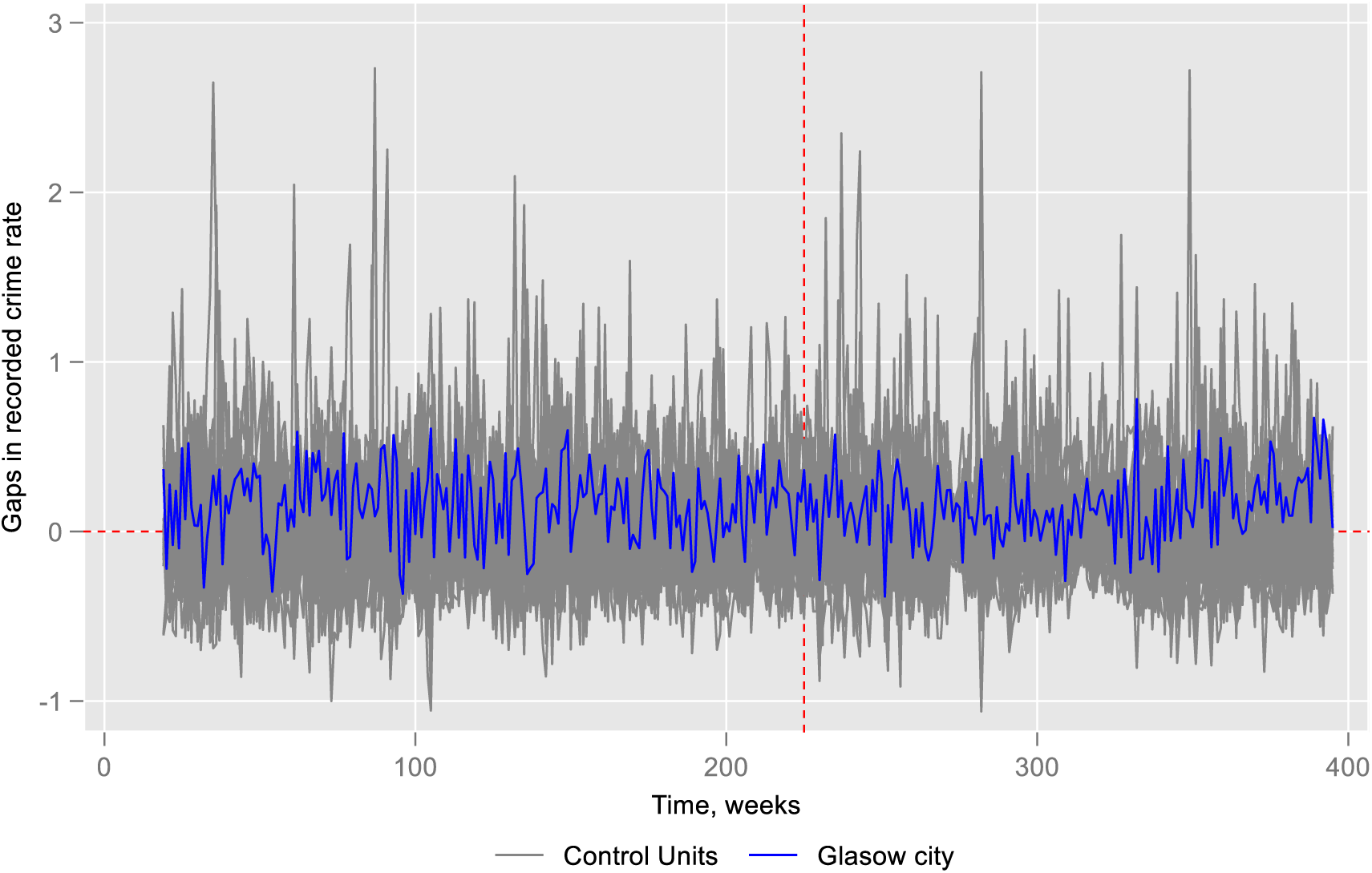
Placebo test.

**Figure SC 18b:**
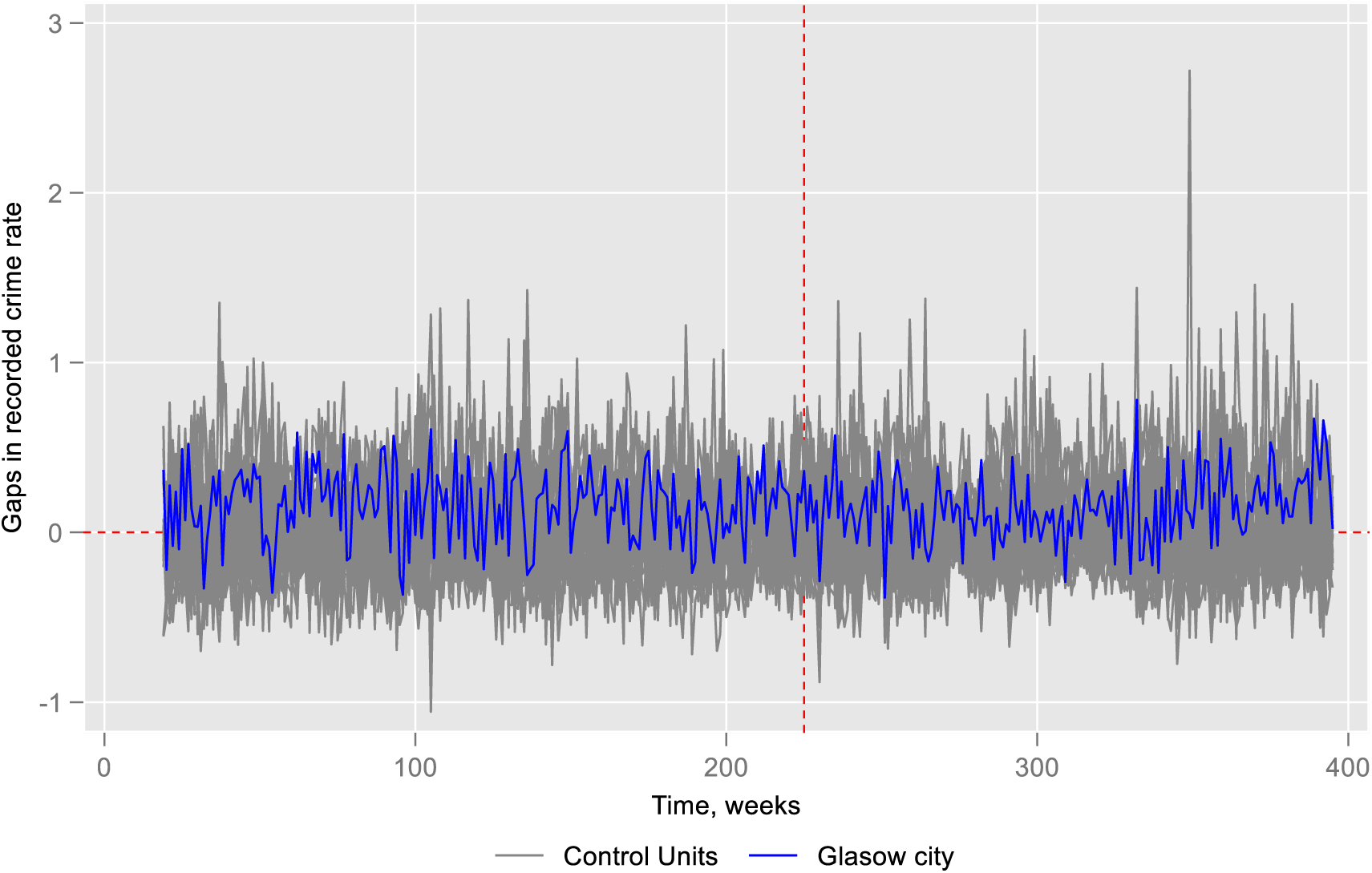
Placebo test excluding control cities MSPE>2 times Glasgow.

**Figure SC 18c:**
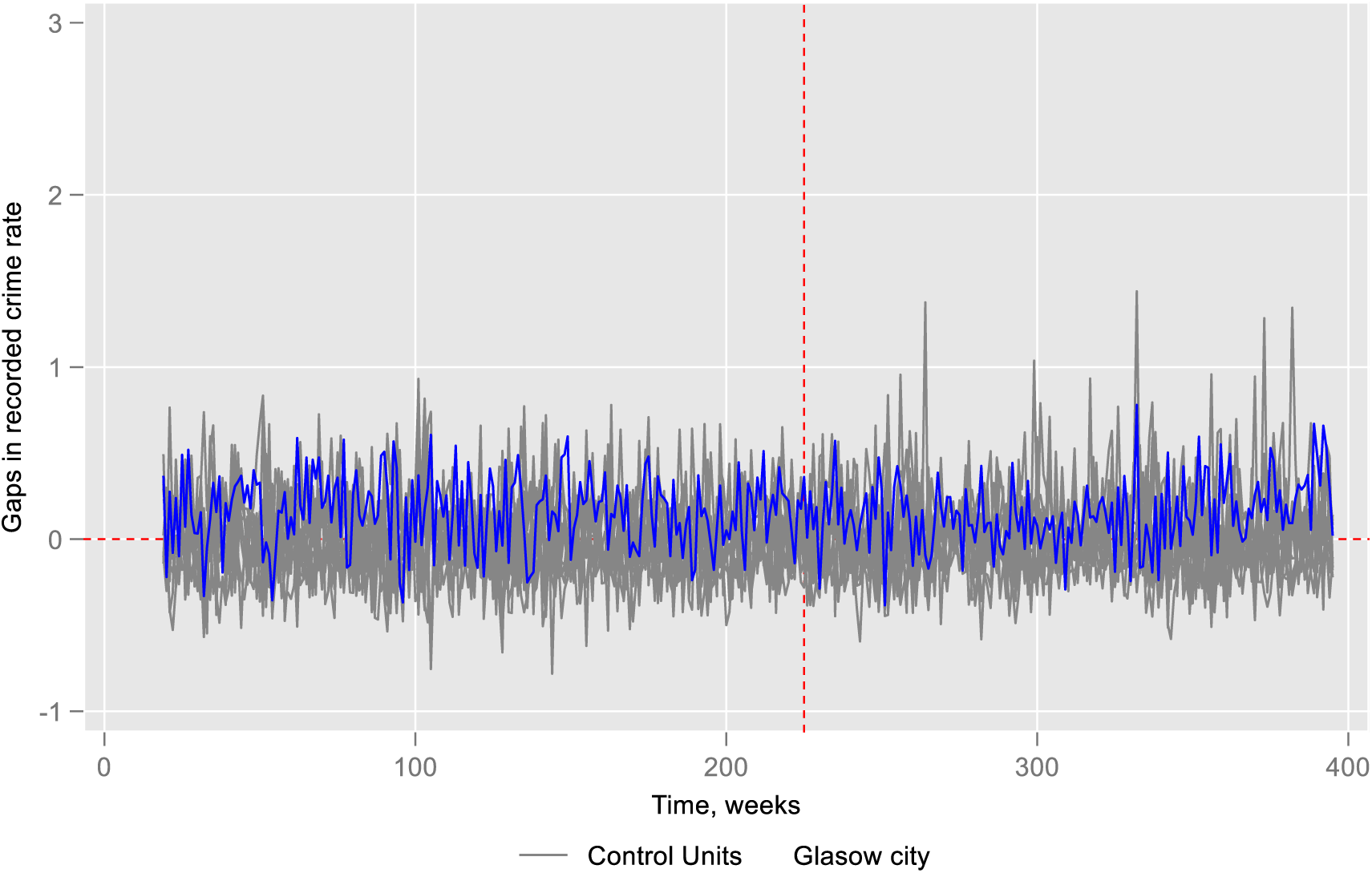
Placebo test excluding control cities if MSPE> Glasgow.

**Figure SC 19a:**
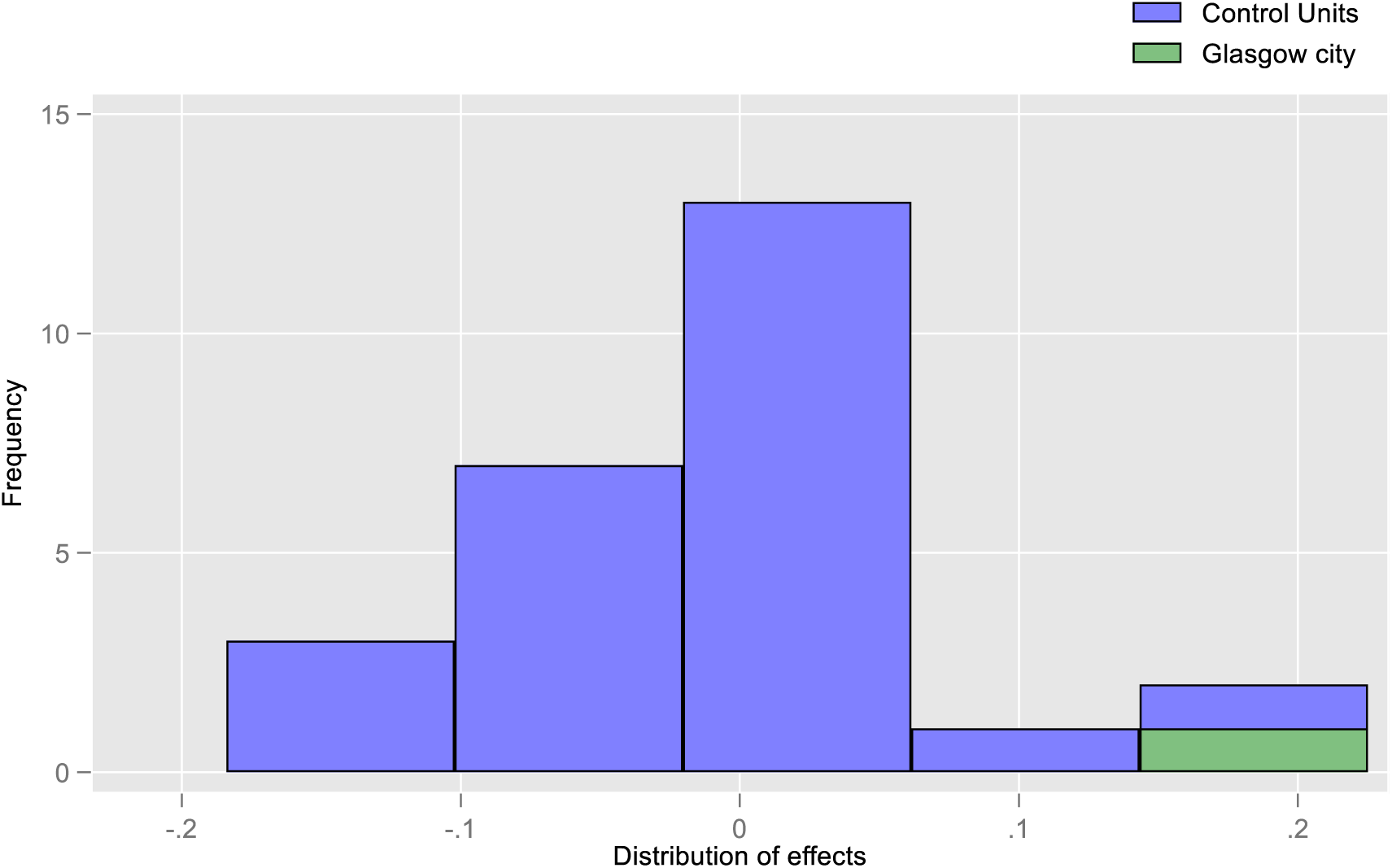
Effect size for placebo test.

**Figure SC 19b:**
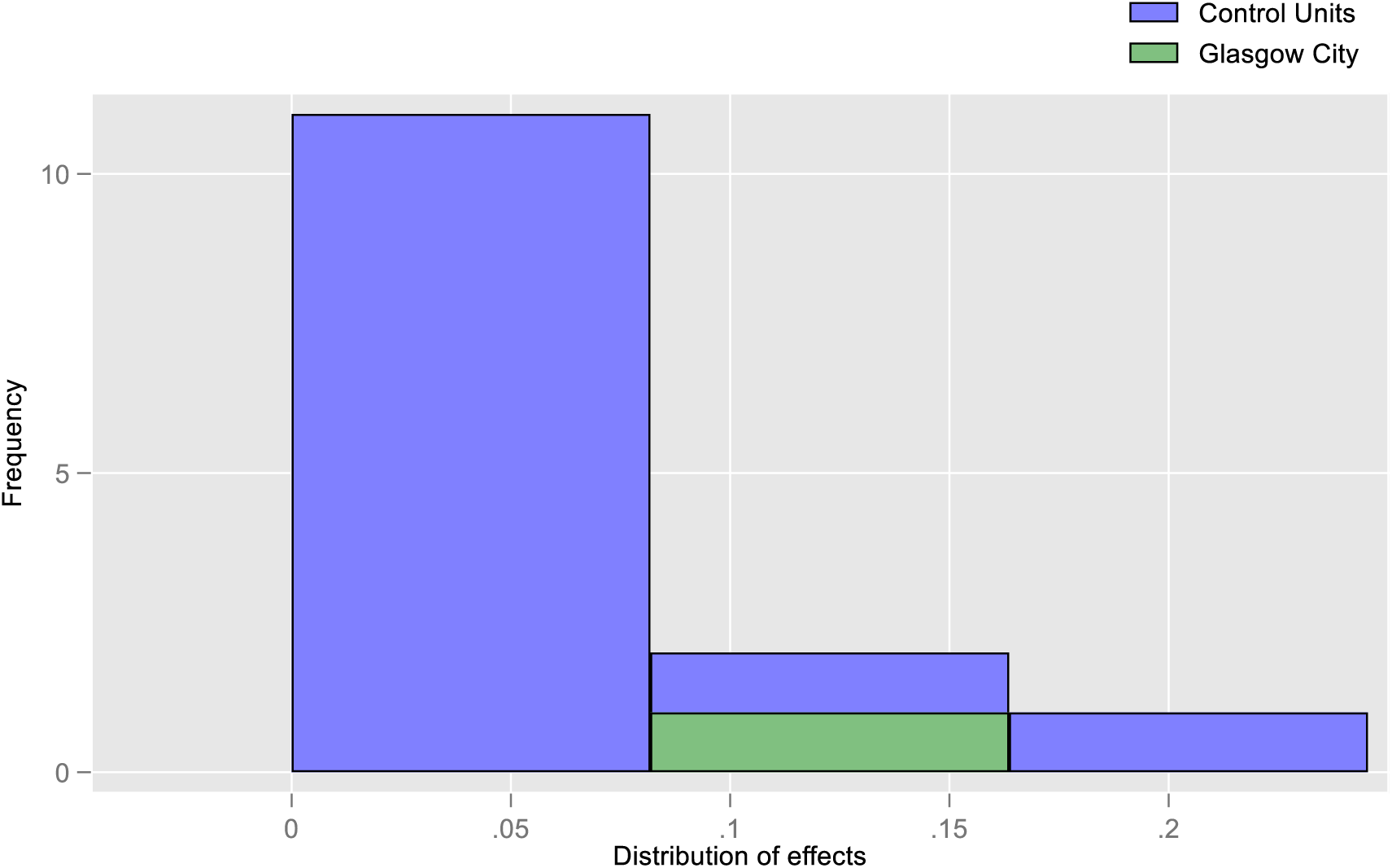
Effect size placebo test considering only positive effect size.

**Figure SC 20a:**
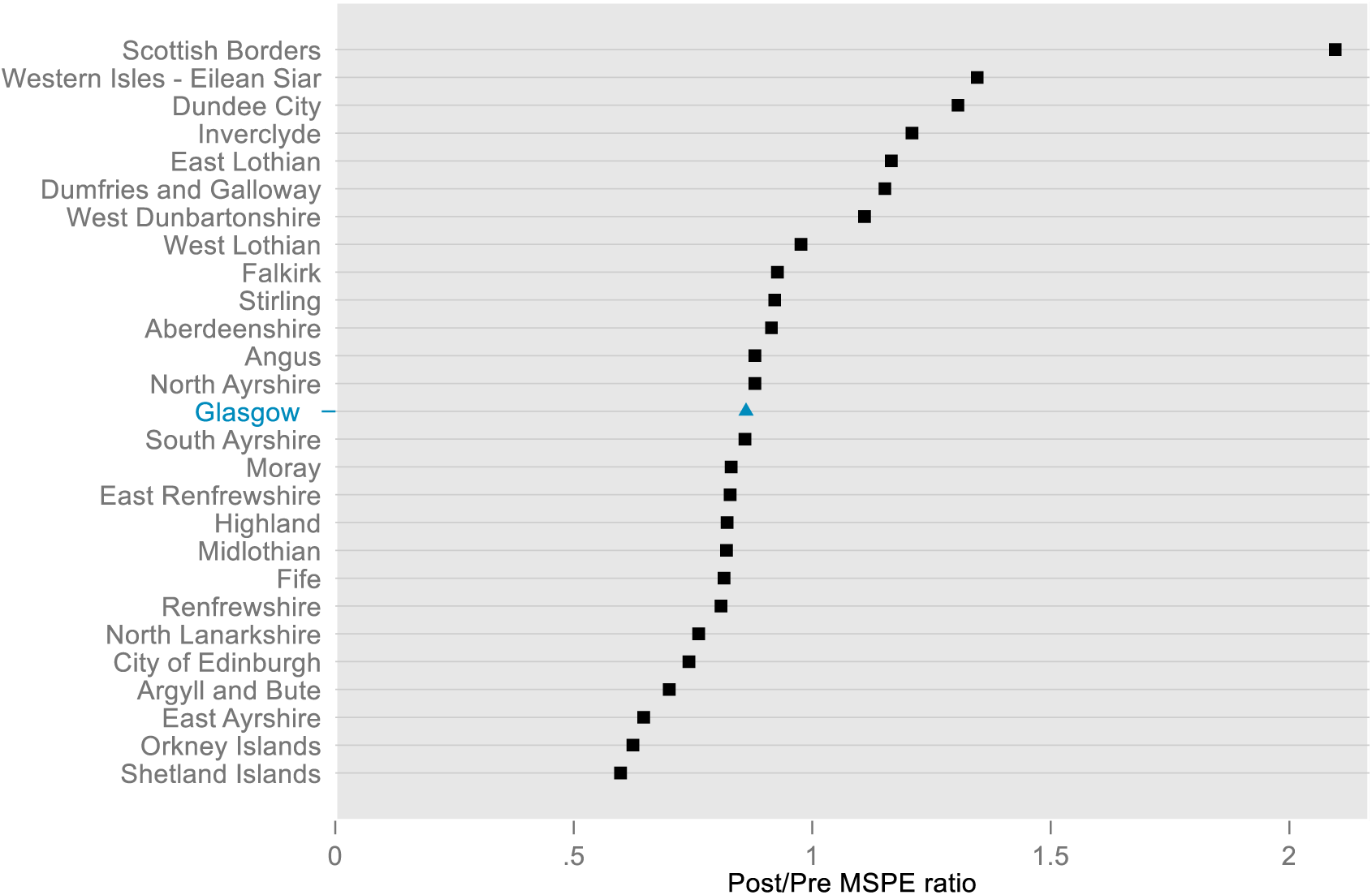
Post/Pre MSPE ratio across cities.

**Figure SC 20b:**
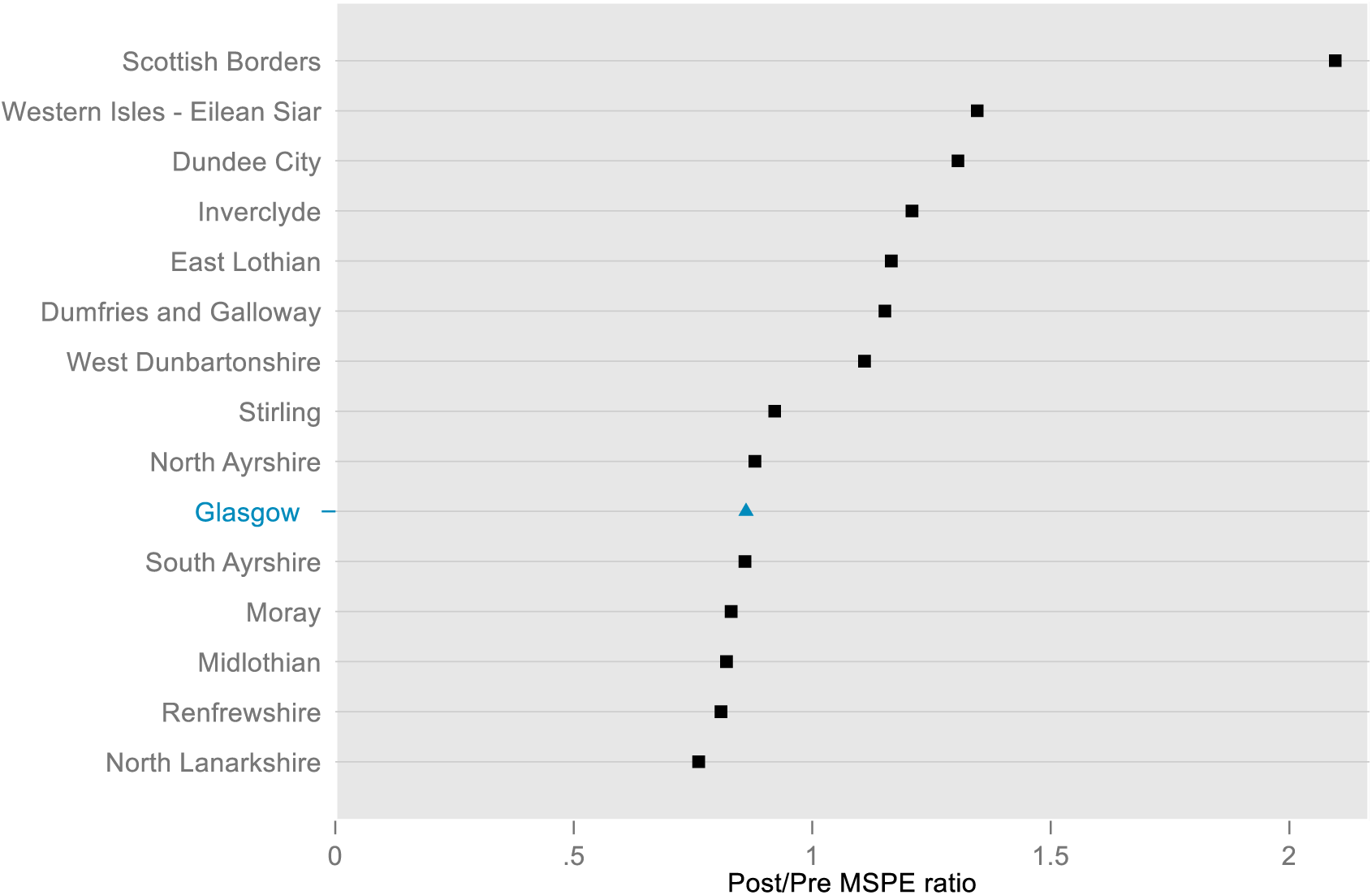
Post/Pre MSPE ratio across cities considering only positive effect size.

## Appendix 4. Synthetic control and ARIMA

**Table A17.**
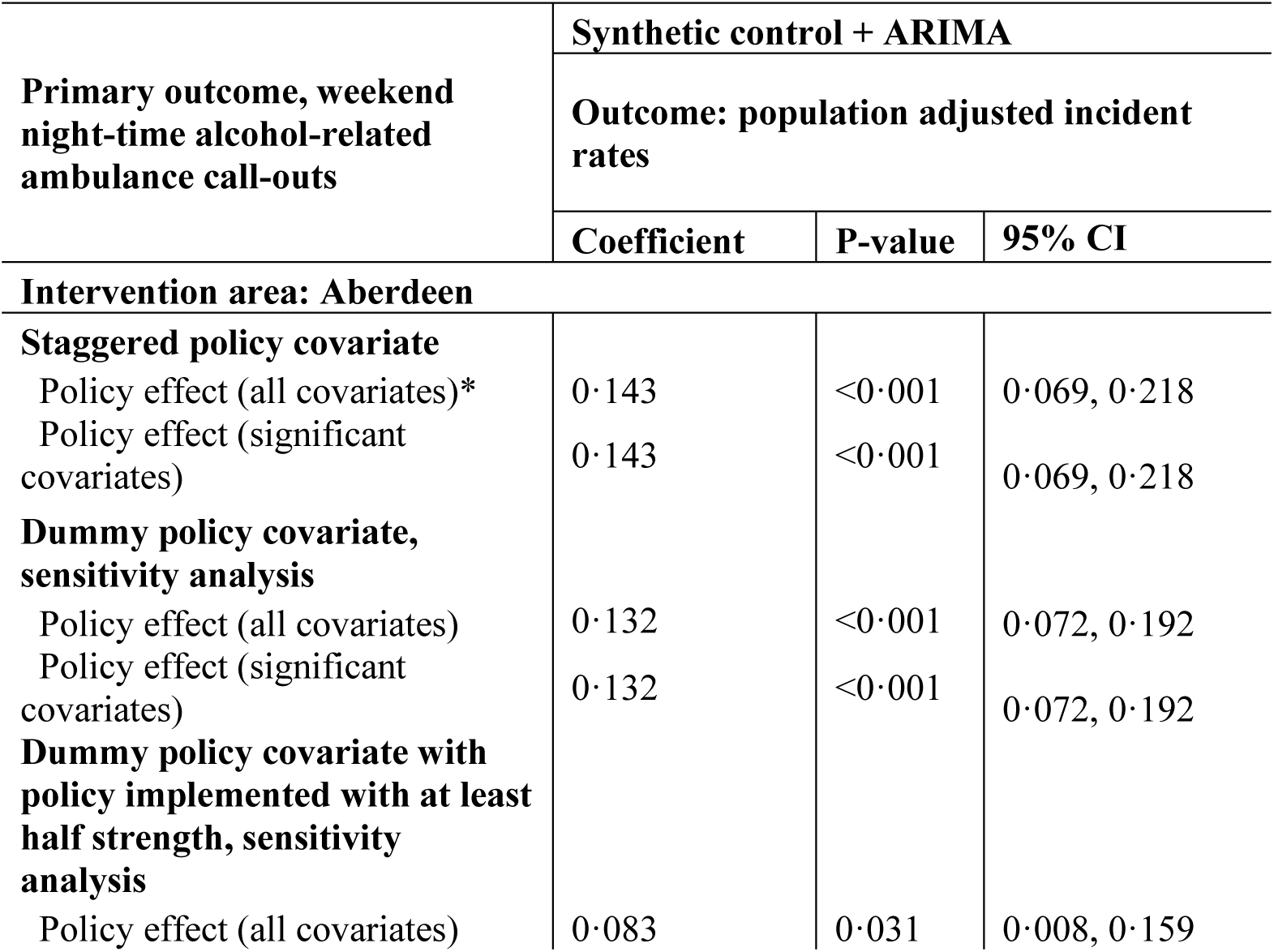
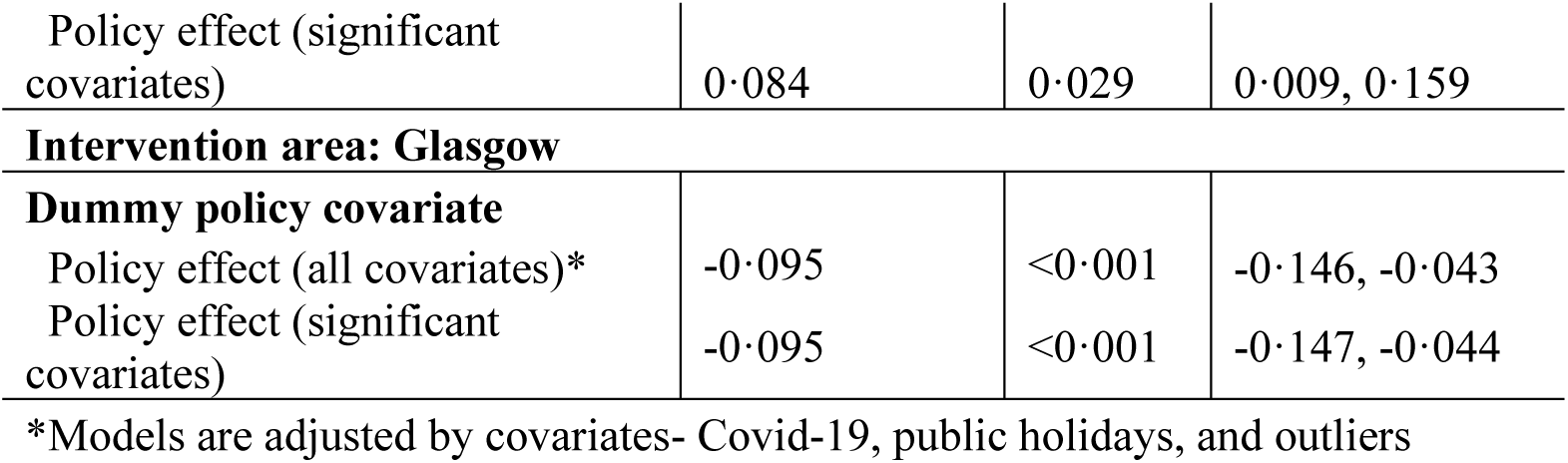
Effect of policy changes on weekend night-time alcohol-related ambulance call-outs in Aberdeen and Glasgow.

| Primary outcome, weekend night-time alcohol-related ambulance call-outs | Synthetic control + ARIMA |  |  |
| --- | --- | --- | --- |
|  | Outcome: population adjusted incident rates |  |  |
|  | Coefficient | P-value | 95% CI |
| **Intervention area: Aberdeen** |  |  |  |
| **Staggered policy covariate** |  |  |  |
| Policy effect (all covariates)* | 0.143 | <0.001 | 0.069, 0.218 |
| Policy effect (significant covariates) | 0.143 | <0.001 | 0.069, 0.218 |
| **Dummy policy covariate, sensitivity analysis** |  |  |  |
| Policy effect (all covariates) | 0.132 | <0.001 | 0.072, 0.192 |
| Policy effect (significant covariates) | 0.132 | <0.001 | 0.072, 0.192 |
| **Dummy policy covariate with policy implemented with at least half strength, sensitivity analysis** |  |  |  |
| Policy effect (all covariates) | 0.083 | 0.031 | 0.008, 0.159 |
| Policy effect (significant covariates) | 0·084 | 0·029 | 0·009, 0·159 |
| **Intervention area: Glasgow** |  |  |  |
| **Dummy policy covariate** |  |  |  |
| Policy effect (all covariates)* | -0·095 | <0·001 | -0·146, -0·043 |
| Policy effect (significant covariates) | -0·095 | <0·001 | -0·147, -0·044 |

\*Models are adjusted by covariates- Covid-19, public holidays, and outliers

**Table A18.** Effect of policy changes on weekend night-time recorded crimes in Aberdeen and Glasgow.

| Secondary outcome, weekend night-time recorded crimes | Synthetic control + ARIMA |  |  |
| --- | --- | --- | --- |
|  | Outcome: population adjusted incident rates |  |  |
|  | Coefficient | P-value | 95% CI |
| **Intervention area: Aberdeen** |  |  |  |
| **Staggered policy covariate** |  |  |  |
| Policy effect (all covariates)* | -0·029 | 0·599 | -0·136, 0·079 |
| Policy effect (significant covariates) | -0·032 | 0·488 | -0·123, 0·059 |
| **Dummy policy covariate, sensitivity analysis** |  |  |  |
| Policy effect (all covariates) | -0·032 | 0·444 | -0·116, 0·051 |
| Policy effect (significant covariates) | -0·035 | 0·393 | -0·115, 0·045 |
| **Dummy policy covariate with policy implemented with at least half strength, sensitivity analysis** |  |  |  |
| Policy effect (all covariates) | 0·007 | 0·876 | -0·081, 0·094 |
| Policy effect (significant covariates) | -0·001 | 0·985 | -0·076, 0·075 |
| **Intervention area: Glasgow** |  |  |  |
| **Dummy policy covariate** |  |  |  |
| Policy effect (all covariates)* | 0·029 | 0·268 | -0·22, 0·080 |
| Policy effect (significant covariates) | 0·029 | 0·269 | -0·022, 0·080 |

\*Models are adjusted by covariates- Covid-19, public holidays, and outliers

## REFERENCES

1 Brauer M, Roth GA, Aravkin AY, et al. Global burden and strength of evidence for 88 risk factors in 204 countries and 811 subnational locations, 1990–2021: a systematic analysis for the Global Burden of Disease Study 2021. Lancet. 2024;403:2162–203.

2 World Health Organization. Global status report on alcohol and health 2018. Geneva: World Health Organization 2018. 10.1037/cou0000248

3 Gakidou E, Afshin A, Abajobir AA, et al. Global, regional, and national comparative risk assessment of 84 behavioural, environmental and occupational, and metabolic risks or clusters of risks, 1990-2016: A systematic analysis for the Global Burden of Disease Study 2016. Lancet. 2017;390:1345–422.

4 Griswold MG, Fullman N, Hawley C, et al. Alcohol use and burden for 195 countries and territories, 1990-2016: A systematic analysis for the Global Burden of Disease Study 2016. Lancet. 2018;392:1015–35.

5 National Records of Scotland. Alcohol-specific deaths 2023. 2024. https://www.nrscotland.gov.uk/statistics-and-data/statistics/statistics-by-theme/vital-events/deaths/alcohol-deaths

6 Public Health Scotland. Alcohol Related Hospital Statistics Scotland 2022/23. 2023.

7 Manca F, Lewsey J, Waterson R, et al. Estimating the burden of alcohol on ambulance callouts through development and validation of an algorithm using electronic patient records. Int J Environ Res Public Health. 2021;18. doi: 10.3390/ijerph18126363

8 World Health Organization. The SAFER technical package: five areas of intervention at national and subnational levels. 2019. https://apps.who.int/iris/handle/10665/330053

9 Mäkelä P, Warpenius K. Night-time is the right time? Late-night drinking and assaults in Finnish public and private settings. Drug Alcohol Rev. 2020;39:321–9.

10 Wilkinson C, Livingston M, Room R. Impacts of changes to trading hours of liquor licences on alcohol-related harm: A systematic review 2005-2015. Public Heal Res Pract. 2016;26:1–7.

11 Nepal S, Kypri K, Tekelab T, et al. Effects of extensions and restrictions in alcohol trading hours on the incidence of assault and unintentional injury: Systematic review. J Stud Alcohol Drugs. 2020;81:5–23.

12 Popova S, Giesbrecht N, Bekmuradov D, et al. Hours and days of sale and density of alcohol outlets: Impacts on alcohol consumption and damage: A systematic review. Alcohol Alcohol. 2009;44:500–16.

13 Stockwell T, Chikritzhs T. Do relaxed trading hours for bars and clubs mean more relaxed drinking? A review of international research on the impacts of changes to permitted hours of drinking. Crime Prev Community Saf. 2009;11:153–70.

14 Robert A. Hahn, Kuzara JL, Elder R, et al. Effectiveness of Policies Restricting Hours of Alcohol Sales in Preventing Excessive Alcohol Consumption and Related Harms. Am J Prev Med. 2010;39:590–604.

15 Alcohol Focus Scotland. Review of statements of licensing policy 2018 to 2023. Published Online First: 2020.

16 Humphreys DK, Eisner MP. Evaluating a natural experiment in alcohol policy The Licensing Act (2003) and the requirement for attention to implementation. Criminol Public Policy. 2010;9:41–67.

17 Humphreys DK, Eisner MP, Wiebe DJ. Evaluating the Impact of Flexible Alcohol Trading Hours on Violence: An Interrupted Time Series Analysis. PLoS One. 2013;8. doi: 10.1371/journal.pone.0055581

18 Stevely AK, de Vocht F, Neves RB, et al. Evaluating the effects of the Licensing Act 2003 on the characteristics of drinking occasions in England and Wales: a theory of change-guided evaluation of a natural experiment. Addiction. 2021;116:2348–59.

19 Sheikh N, Henriques-Cadby I, Haghpanahan H, et al. Evaluating later or expanded premises hours for alcohol in the night-time economy (ELEPHANT): statistical analysis plan. medRxiv. Published Online First: 2024. doi: 10.1101/2024.05.07.24306991

20 Abadie A, Diamond A, Hainmueller J. Synth: An R Package for Synthetic Control Methods in Comparative Case Studies. J Stat Softw. 2011;42:1–17.

21 Degli Esposti M, Spreckelsen T, Gasparrini A, et al. Can synthetic controls improve causal inference in interrupted time series evaluations of public health interventions? Int J Epidemiol. 2020;49:2010–20.

22 Justice Analytical Services. Bringing Together Scotland ’ s Crime Statistics. Edinburgh: Scotland 2014.

23 de Goeij MCM, Veldhuizen EM, Buster MCA, et al. The impact of extended closing times of alcohol outlets on alcohol-related injuries in the nightlife areas of Amsterdam: A controlled before-and-after evaluation. Addiction. 2015;110:955–64.

24 Mitchell, et al. ‘It’s a bit of a mess’: The impact of later alcohol trading hours for bars and clubs in Scotland according to qualitative interviews with local stakeholders. 2025.

25 McClelland R, Mucciolo L. An update on the synthetic control method as a tool to understand state policy. 2022. https://www.taxpolicycenter.org/sites/default/files/publication/163919/an_update_on_the_synthetic_control_method_as_a_tool_to_understand_state_policy.pdf

26 Huntington-Klein N. *The Effect : An introduction to Research Design and Causality*. 1st ed. Chapman & Hall 2022.

27 Department of Justice of Ireland. General Scheme: Sale of Alcohol Bill 2022 including press release. 2022. https://www.drugsandalcohol.ie/37347/ (accessed 29 October 2025)

28 Department for Business & Trade. Licensing policy sprint: joint industry and HM government taskforce report. Policy paper. London 2025. https://www.gov.uk/government/publications/licensing-taskforce-report-and-government-response/licensing-policy-sprint-joint-industry-and-hm-government-taskforce-report

29 Department for Business & Trade. Licensing taskforce report: government response. Policy paper. London 2025. https://www.gov.uk/government/publications/licensing-taskforce-report-and-government-response/licensing-taskforce-report-government-response

30 Fitzgerald N, Egan M, O’Donnell R, et al. Public health engagement in alcohol licensing in England and Scotland: the ExILEnS mixed-method, natural experiment evaluation. Public Heal Res (Southampton, England). 2024;13:1–84.

31 Scottish Government. Licensing ( Scotland ) Act 2005. 2005. https://www.legislation.gov.uk/asp/2005/16/contents

32 City of Glasgow Licensing Board. Licensing Policy Statement: Licensing (Scotland) Act 2005. 2018. https://www.glasgow.gov.uk/CHttpHandler.ashx?id=17578&p=0

33 Home Office. Revised guidance issued under section 182 of Licensing Act 2003. Published Online First: 2025.

34 Scottish Government. Quarterly National Accounts Scotland, 2017 Quarter 3. 2018. https://www.webarchive.org.uk/wayback/archive/20180212144444/http://www.gov.scot/Topics/Statistics/Browse/Economy/QNA2017Q3 (accessed 22 November 2022)

35 Centre for Environmental Data Analysis. Met Office weather datasets. CEDA Arch. 2019. https://data.ceda.ac.uk/badc/ukmo-hadobs/data/insitu/MOHC/HadOBS/HadUK-Grid/v1.2.0.ceda/region (accessed 10 November 2023)

36 The Scottish Government. Scottish Liquor Licensing Statistics. 2022. https://www.gov.scot/publications/scottish-liquor-licensing-statistics/

37 Scottish Parliament Information Centre (SPICe). Timeline of Coronavirus (COVID-19) in Scotland. SPICe Spotlight. 2023. https://spice-spotlight.scot/2023/05/10/timeline-of-coronavirus-covid-19-in-scotland/ (accessed 11 March 2024)

38 The Scottish Government. Public and bank holidays. Scotl. bank holidays. 2022. https://www.mygov.scot/scotland-bank-holidays (accessed 12 March 2024)

39 Abadie A, Vives-i-Bastida J. Synthetic Controls in Action. arXiv Prepr. Published Online First: 2022.

40 Ferman B, Pinto C, Possebom V. Cherry Picking with Synthetic Controls. J Policy Anal Manag. 2020;39:510–32.

41 Kaul A, Klößner S, Pfeifer G, et al. Standard Synthetic Control Methods: The Case of Using All Preintervention Outcomes Together With Covariates. J Bus Econ Stat. 2022;40:1282–90.

42 Kreif N, Grieve R, Hangartner D, et al. Examination of the synthetic control method for evaluating health policies with multiple treated units. Health Econ. 2015;25:1514– 28.

